# ̃Evaluating reduction in CoViD-19 cases by isolation and protective measures in São Paulo State, Brazil, and scenarios of release

**DOI:** 10.1101/2020.05.19.20099309

**Authors:** Hyun Mo Yang, Luis Pedro Lombardi Junior, Fabio Fernandes Morato Castro, Ariana Campos Yang

**Affiliations:** Department of Applied Mathematics, State University of Campinas Praça Sérgio Buarque de Holanda, 651; CEP: 13083-859, Campinas, SP, Brazil; Department of Applied Mathematics, State University of Campinas Praga Sérgio Buarque de Holanda, 651; CEP: 13083-859, Campinas, SP, Brazil; Division of Allergy and Immunology, General Hospital of the Medicine School of University of São Paulo Av. Dr. Eneas Carvalho de Aguiar, 255; CEP: 05403-000, São Paulo, SP, Brazil

**Keywords:** mathematical model, new coronavirus, pulses of isolation and release, personal and collective protective measures, lockdown

## Abstract

São Paulo State registered the first case of CoViD-19 on 26 February, the first death due to CoViD-19 on 16 March, and implemented the isolation of the population in non-essential activities on 24 March, which is programmed to end on 1 June. A mathematical model considering young (below 60 years old) and elder (above 60 years) subpopulations was formulated based on the natural history of CoViD-19 to study the transmission of the new coronavirus in São Paulo State, Brazil. This deterministic model used the data collected in São Paulo State to estimate the model parameters and to evaluate the effects of herd protection, that is, isolation and personal and collective protective measures. Based on the estimated parameters, we evaluated the scenarios of three releases divided in equal proportions elapsed by 14 days between releases, but beginning in three different times (the first release occurring on 1 and 23 June, and 6 July). We concluded that these three strategies of release are equivalent (little difference) in reducing the number of severe CoViD-19 if social behaviour does not change. However, if protective measures as using face mask and hygiene (washing hands, for instance) and social distancing could be massively disseminated in the population to decrease the transmission of CoViD-19 by 80%, we concluded that the health care system may not collapse with release.

## 1 Introduction

Coronavirus disease 2019 (CoViD-19), which is caused by severe acute respiratory syndrome coronavirus 2 (SARS-CoV-2), a strain of the RNA-based SARS-CoV-1,was declared a pandemic by the World Health Organization (WHO) on 11 March 2020. The rapid spreading of SARS-CoV-2 (new coronavirus) is due to the fact that this virus can be transmitted by droplets that escape the lungs through coughing or sneezing and infect humans (direct transmission), or they are deposited in surfaces and infect humans when in contact with this contaminated surface (indirect transmission). This virus enters into susceptible persons through the nose, mouth, or eyes, and infects cells in the respiratory tract, being capable to release millions of new viruses. In serious cases, due to new coronavirus infection, immune cells overreact and attack the lung cell causing acute respiratory disease syndrome and possibly death. In general, the fatality rate in elder patients (60 years or more) is much higher than the population average.

Currently, there is not a vaccine, neither an effective treatment. Hence, isolation is the main, if not unique, way of controlling the dissemination of the new coronavirus in a population by flattening the curve of epidemics. However, the isolation as a control measure arises an important question: are there reliable strategies to release these isolated persons aiming to avoid the retaken of its original progression of infection?

Mathematical models allow us to understand the progression of viral infections if the natural history of the disease is well documented. Based on this knowledge being improved as epidemics evolves, in [31] we considered continuous isolation and release rates in the modelling to describe new coronavirus epidemics, which was improved by considering intermittent pulses in isolation and release in [32]. In both models, asymptomatic and severe CoviD-19 persons are allowed to transmit virus. Here, we improve previous models allowing transmission by mild CoViD-19 persons and incorporating personal (face mask, hygiene, etc.) and collective (social distancing) protective measures that reduce the transmission of virus. Taking into account the pulses in isolation and releases, we describe the current epidemiological status of São Paulo State, Brazil, by estimating the model parameters. Based on these estimated parameters, we evaluate the epidemiological scenarios arising from release beginning on 1 June.

The paper is structured as follows. In Section 2, we introduce a model, which is numerically studied in Section 3. Discussions are presented in Section 4, and conclusions in Section 5.

## 2 Material and methods

In a community where SARS-CoV-2 (new coronavirus) is circulating, the risk of infection is greater in elder than young persons, as well as elder persons are under increased probability of being symptomatic and higher CoViD-19 induced mortality. Hence, the community is divided into two groups, composed by young (under 60 years old, denoted by subscript *y*), and elder (above 60 years old, denoted by subscript *o*) persons. The vital dynamics of this community is described by per-capita rates of birth (*ϕ*) and mortality (*μ*).

For each subpopulation *j* (*j* = *y*, *o*), all persons are divided into nine classes: susceptible *S_j_*, susceptible persons who are isolated *Q_j_*, exposed and incubating *E_j_*, asymptomatic *A_j_*, symptomatic persons in the initial phase of CoViD-19 (or pre-diseased) *D*_1_*_j_*, pre-diseased persons caught by test and then isolated *Q*_1_*_j_*, symptomatic persons with severe CoViD-19 *D*_2_*_j_*, mild CoViD-19 *Q*_2_*_j_*, and mild CoViD-19 persons isolating themselves by educational campaign *Q*_3_*_j_*. However, all young and elder persons in classes *A_j_*, *Q*_1_*_j_*, *Q*_2_*_j_*, *Q*_3_*_j_* and *D*_2_*_j_* enter into the same immune class I (this is the 10^th^ class, but common to both subpopulations).

The natural history of new coronavirus infection is the same for young (*j* = *y*) and elder (*j* = *o*) subpopulations. We assume that persons in the asymptomatic (*A_j_*), pre-diseased (*D*_1_*_j_*), and a fraction *z_j_* of mild CoViD-19 (*Q*_2_*_j_*) classes are transmitting the virus, and other infected classes (*Q*_1_*_j_*, (1 − *z_j_*) *Q*_2_*_j_* and *D*_2_*_j_*) are under voluntary or forced isolation. Susceptible persons are infected according to *λ_j_S_j_* (known as the mass action law [4]) and enter into class *E_j_*, where *λ<u>_j_</u>* is the per-capita incidence rate (or force of infection) defined by *λ_j_* = *λ* (*δ_jy_* + *ψδ_jo_*,), with *λ* being

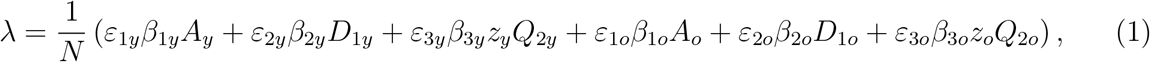

where *δ_ij_* is Kronecker delta, with *δ_ij_* = 1 if *i* = *j*, and 0, if *i* ≠ *j*; and *β*_1_*_j_*, *β*_2_*_j_* and *β*_3_*_j_* are the transmission rates, that is, the rates at which a virus encounters a susceptible people and infects him/her, and *ε*_1_*_j_*, *ε*_2_*_j_* and *ε*_3_*_j_* are reduction factors due to protective measures adopted by population. After an average period 1/*σ_j_* in class *E_j_*, where *σ_j_* is the incubation rate, exposed persons enter into the asymptomatic class *A_j_* (with probability *p_j_*) or pre-diseased class *D*_1_*_j_* (with probability 1 − *p_j_*). After an average period 1/*γ_j_* in class *A_j_*, where *γ_j_* is the recovery rate of asymptomatic persons, symptomatic persons acquire immunity (recovered) and enter into immune class I. Possibly asymptomatic persons can manifest symptoms at the end of this period, and a fraction 1 − *χ_j_* enters into mild CoViD-19 class *Q*_2_*_j_*. Another route of exit from class *A_j_* is being caught by a test at a rate *η_j_* and enter into class I (we assume that this person indeed adopts isolation, which is the reason to enter into class I at a rate of testing). With respect to symptomatic persons, after an average period 1/*γ_j_* in class *D*_1_*_j_*, where *γ*_1_*_j_* is the infection rate of pre-diseased persons, pre-diseased persons enter into severe CoViD-19 class *D*_2_*_j_* (with probability 1 − *m_j_*) or class *Q*_2_*_j_* (with probability *m_j_*), or they are caught by test at a rate *η*_1_*_j_* and enter into class *Q*_1_*_j_*. Persons in class *D*_2_ acquire immunity after period 1/*γ*_2_*_j_*, where *γ*_2_*_j_* is the recovery rate of severe CoViD-19, and enter into class *I* or die under the disease induced (additional) mortality rate *α_j_*. Another route of exiting class *D*_2_*_j_* is by treatment, described by the treatment rate *θ_j_*. Class *Q*_1_*_j_* is composed by mild and severe CoViD-19 persons who came from class *D*_1_*_j_* caught by test, hence they enter into class *D*_2_*_j_* (with rate (1 − m*_j_*) *Y_j_*) or class I (with rate *m_j_γ_j_* + *γ*_2_*_j_*, assuming adherence to isolation). Persons in class *Q*_2_*_j_* acquire immunity after period 1/*γ*_3_*_j_*, where *γ*_3_*_j_* is the recovery rate of mild CoViD-19, and enter into immune class *I*. Another route of exit from class *Q*_2_*_j_* are being caught by a test at a rate *η*_2_*_j_* and enter into class I (assumption of adherence to isolation), or enter to class *Q*_3_*_y_* convinced by an education campaign at a rate *ε*_4_*_j_*, which is temporary, hence ξ*_j_* is the rate of abandonment of protective measures [32].

In the model, we consider pulse isolation and intermittent (series of pulses) release of persons. We assume that there is a unique pulse in isolation at time 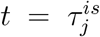 described by 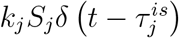, but there are *m* intermittent releases described by 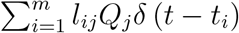, where 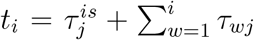 *j* = *y*, *o*, amd *δ*(*x*) is Dirac delta function, that is, *δ*(*x*) = ∞, if *x* = 0, otherwise, *δ*(*x*) = 0, with 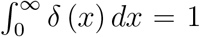. The fraction of persons in isolation is *k_j_*, and *l_ij_, i* = 1,2,…*,m*, is the fraction of *i*-th release of isolated persons, with *τ_wj_* being the period between successive releases.

Figure 1 shows the flowchart of the new coronavirus transmission model.

**Figure 1:**
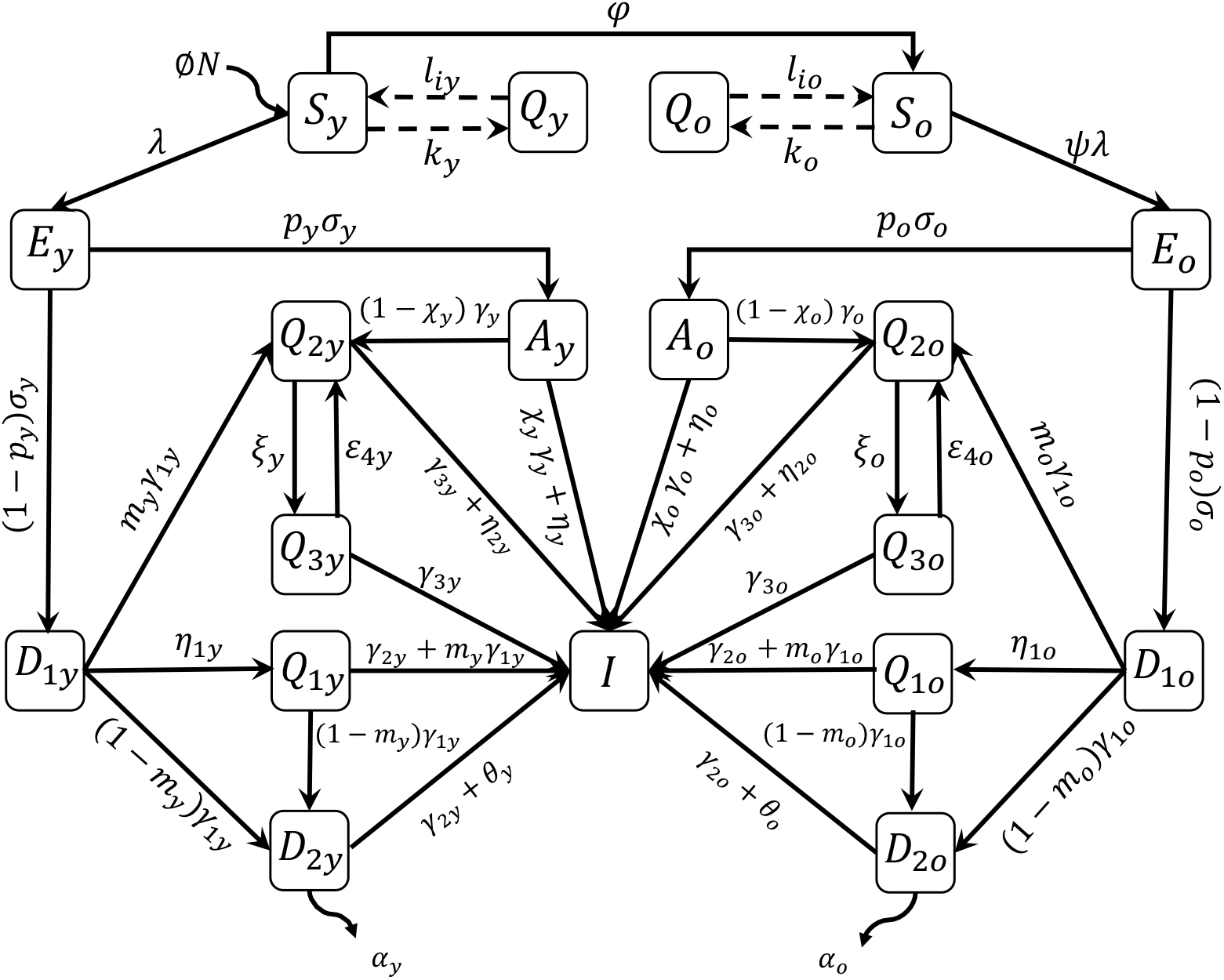
The flowchart of new coronavirus transmission model with variables and parameters.

The new coronavirus transmission model, based on the above descriptions summarized in Figure 1, is described by the system of ordinary differential equations, with *j = y, o*. Equations for susceptible persons are

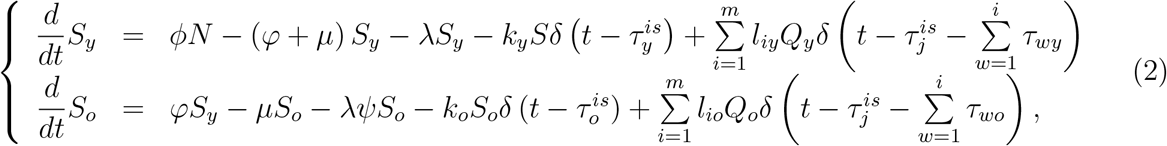

for infectious persons,

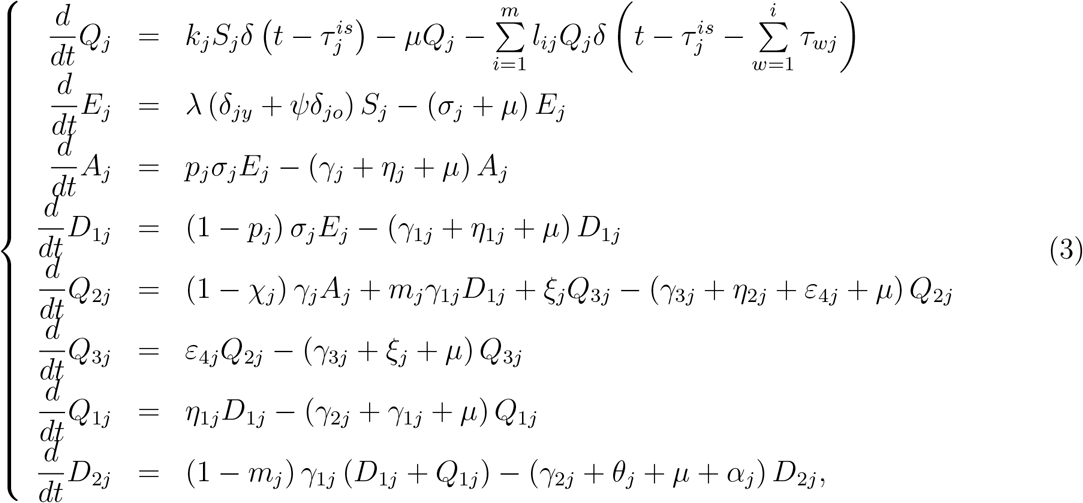

and for immune persons,

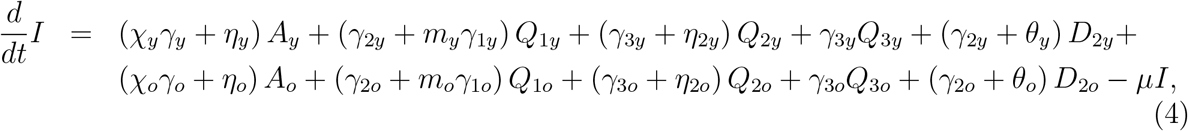

where *N_j_* = *S_j_* + *Q_j_* + *E_j_* + *A_j_* + *D*_1_*_j_* + *Q*_1_*_j_*, + *Q*_2_*_j_*, + *Q*_3_*_j_*, + *D*_2_*_j_*, and *N* = *N_y_* + *N_o_* + *I* obeys

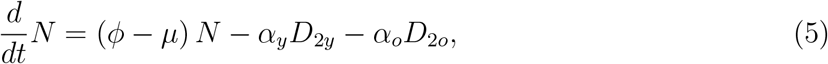

with the initial number of population at *t* = 0 being *N*(0) = *N*_0_ = *N*_0_*_y_* + *N*_0_*_o_*, where *N*_0_*_y_* and *N*_0_*_o_* are the size of young and elder subpopulations at *t* = 0. If *ϕ* = *μ* + (*α_y_D*_2_*_y_* + *α_o_D*_2_*_o_*) */N*, the total size of the population is constant.

Table 1 summarizes the model classes (or variables).

**Table 1:** Summary of the model variables (*j* = *y*, *o*).

| Symbol | Meaning |
| --- | --- |
| $S_j$ | Susceptible persons |
| $Q_j$ | Isolated among susceptible persons |
| $E_j$ | Exposed and incubating new coronavirus persons |
| $A_j$ | Asymptomatic persons |
| $D_{1j}$ | Pre-diseased (pre-symptomatic) persons |
| $Q_{1j}$ | Pre-diseased persons caught by test |
| $Q_{2j}$ | Mild (non-hospitalized) CoViD-19 persons |
| $Q_{3j}$ | Mild CoViD-19 persons adhered to isolation |
| $D_{2j}$ | Severe (hospitalized) CoViD-19 persons |
| $I$ | Immune (recovered) persons |

The non-autonomous system of equations (2), (3), and (4) is simulated permitting intermittent interventions to the boundary conditions. Hence, the equations for susceptible and isolated persons become

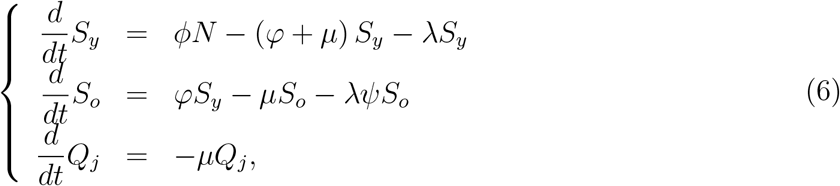

*j* = *y*, *o*, and other equations are the same.

For the system of equations (3), (4) and (6), the initial conditions (at *t* = 0) are, for *j* = *y*, *o*,

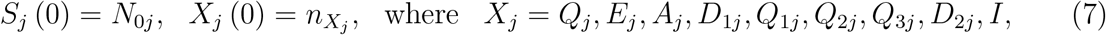

and is a non-negative number. For instance, 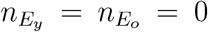 means that there is not any exposed person (young and elder) at the beginning of epidemics. We split the boundary conditions into isolation and release, and assume that 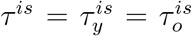 and *τ_i_* =*τ_iy_* = *τ_io_*, for *i* = 1, 2,…,*m*, then 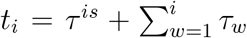. A unique isolation at *t* = *τ^i^*^s^ is described by the boundary conditions

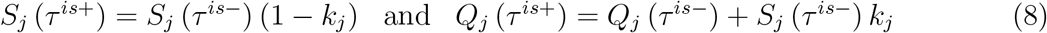

plus

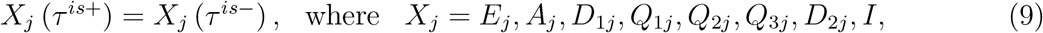

where we have 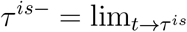 (for *t* < *τ^is^*), and 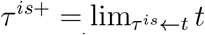 (for *t* > *τ^is^*). The boundary conditions for a series of pulses released at 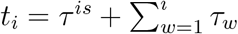 for *i* =1, 2,…,*m*, are

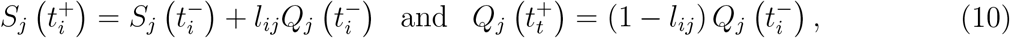

plus

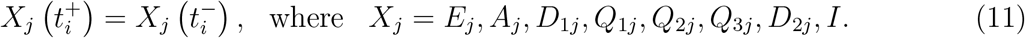

If *τ_i_* = *τ*, then *t_i_* = *τ^is^* + *iτ*. If isolation is applied to a completely susceptible population, at *t* = 0, there is not any infectious people, so *S*(0) = *N*_0_. If isolation is done at 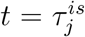 without screening of persons harbouring the virus, then many of the asymptomatic persons could be isolated with susceptible persons. In the Appendix A, we analyze the steady state attained by the fractions of persons in each class (compartment).

Table 2 summarizes the model parameters and values (for elder classes, values are between parentheses), see Appendix B.

From the solution of the system of equations (3), (4) and (6), we can derive some epidemiological parameters: susceptible persons, accumulated and daily cases of severe CoViD-19, number of occupied beds by inpatient, ICU and ICU/intubated persons, number of deaths due to CoViD-19, and number of cured persons. All initial conditions below are determined by the initial conditions (7) supplied to the system of equations.

**Table 2:** Summary of the model parameters (*j* = *y,o*) and values (rates in *days^−^*^1^, time in *days* and proportions are dimensionless). Some values are calculated (#), or assumed (*), or estimated (**) or not available yet (***).

| Symbol | Meaning | Value |
| --- | --- | --- |
| $\mu$ | Natural mortality rate | $1/(75 \times 365)[15]$ |
| $\phi$ | Birth rate | $1/(75 \times 365)^*$ |
| $\varphi$ | Aging rate | $6.7 \times 10^{-6}\#$ |
| $\sigma_y (\sigma_o)$ | Incubation rate | $1/5.8 (1/5.8)^\#$ |
| $\gamma_y (\gamma_o)$ | Recovery rate of asymptomatic persons | $1/12 (1/14)[19]$ |
| $\gamma_{1y} (\gamma_{1o})$ | Infection rate of pre-diseased persons | $1/4 (1/4)[2]$ |
| $\gamma_{2y} (\gamma_{2o})$ | Recovery rate of severe CoViD-19 | $1/14 (1/21)[19]$ |
| $\gamma_{3y} (\gamma_{3o})$ | Infection rate of mild CoViD-19 persons | $1/13 (1/16)^*$ |
| $\alpha_y (\alpha_o)$ | Additional mortality rate | $0.00053 (0.0053)^{**}$ |
| $\eta_y (\eta_o)$ | Testing rate among asymptomatic persons | $0 (0)^{***}$ |
| $\eta_{1y} (\eta_{1o})$ | Testing rate among pre-diseased persons | $0 (0)^{***}$ |
| $\eta_{2y} (\eta_{2o})$ | Testing rate among mild CoViD-19 persons | $0 (0)^{***}$ |
| $\varepsilon_{4y} (\varepsilon_{4o})$ | Adherence to protection behavior rate | $0.5 (0.5)^{**}$ |
| $\xi_y (\xi_o)$ | Loss of protection behavior rate | $0 (0)^{***}$ |
| $k_y (k_o)$ | Proportion of isolated susceptible persons | $0.53 (0.53)^{**}$ |
| $l_{1y} (l_{1o})$ | Proportion released at first time $t_1$ | $0.33 (0.33)^*$ |
| $\tau^{is}$ | Time of the introduction of isolation | 24 March |
| $\tau_{1y} (\tau_{1o})$ | Time of the first ($i$-th) releasing | 1 June(1 June)* |
| $\theta_y (\theta_o)$ | Treatment rate | $0(0)^{***}$ |
| $\beta_{1y} (\beta_{1o})$ | Transmission rate due to asymptomatic persons | $0.81 (0.93)^{**}$ |
| $\beta_{2y} (\beta_{2o})$ | Transmission rate due to pre-diseased persons | $0.81 (0.93)^{**}$ |
| $\beta_{2y} (\beta_{2o})$ | Transmission rate due to mild CoViD-19 persons | $0.81 (0.93)^{**}$ |
| $z_y (z_o)$ | Proportion circulating of mild CoViD-19 persons | $0.5 (0.2)^*$ |
| $\psi$ | Scaling factor of transmission among elder persons | $1.15^{**}$ |
| $\chi_y (\chi_o)$ | Proportion of remaining as asymptomatic persons | $0.98 (0.95)^{***}$ |
| $p_y (p_o)$ | Proportion of asymptomatic persons | $0.8(0.8)^*$ |
| $m_y (m_o)$ | Proportion of mild (non-hospitalized) CoViD-19 | $0.92^\# (0.75[6])$ |

The number of non-isolated (circulating) persons *S_j_* is obtained from equation (6), and the number of circulating plus isolated susceptible persons *S^tot^* is obtained by

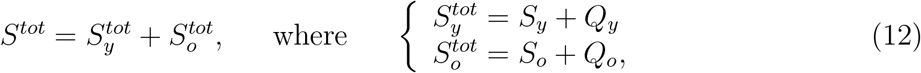

where 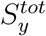 and 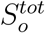 are the numbers of susceptible, respectively, young and elder persons.

The numbers of accumulated severe CoViD-19 cases Ω*_y_* and Ω*_o_* are given by the exits from *D*_1_*_y_*, *Q*_1_*_y_*, *D*_1_*_o_*, and *Q*_1_*_o_*, and entering into classes *D*_2_*_y_* and *D*_2_*_o_*, that is,

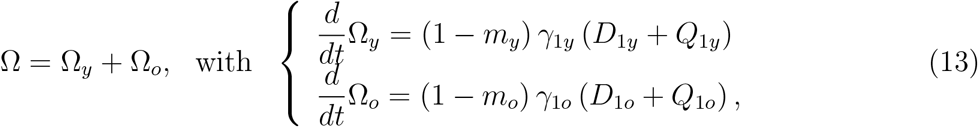

with *Ω_y_*(0) = *Ω_y_*_0_ and *Ω_o_*(0) = *Ω_o_*_0_. The number of deaths due to severe CoViD-19 is

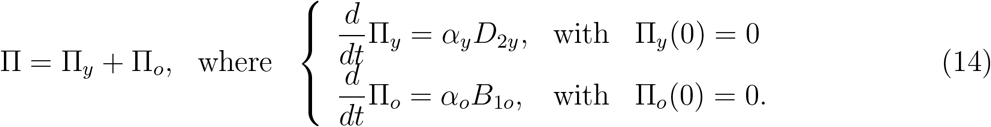

From these two variables Ω and Π, we estimate the model parameters, which are the transmission and additional mortality rates. In the estimation procedure of additional mortality rates, we must bear in mind that the time at which new cases and deaths were registered does not have direct correspondence, rather they are delayed by Δ_1_ days, that is, Π*_y_* (*t* + Δ_1_) = α*_y_D*_2_*_y_*(*t*), for instance.

The daily severe CoViD-19 cases *Q_d_* is, considering Δ*t* = *t_i_* − *t_i_*_−1_ = Δ*t* =1 day,

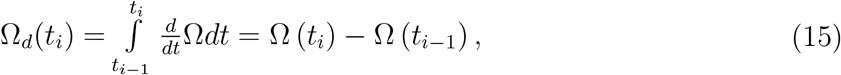

where Ω*_d_*(0) = Ω*_d_*_0_ is the first observed CoViD-19 case (*t*_0_ = 0), with *i* =1, 2,…, and *t*_1_ = 1 is the next day in the calendar time, and so on.

The number of beds occupied by inpatients during the evolving of epidemics is *B*_1_ = *B*_1_*_y_* + *B*_1_*_o_*, for *j* = *y*, *o*, where

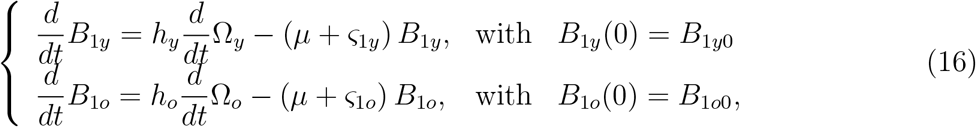

the number of beds occupied by ICU persons is *B*_2_ = *B*_2_*_y_* + *B*_2_*_o_*, where

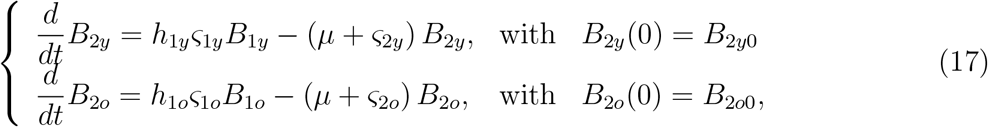

and the number of beds occupied by ICU/intubated persons is *B*_3_ = *B*_3_*_y_* + *B*_3_*_o_*, where

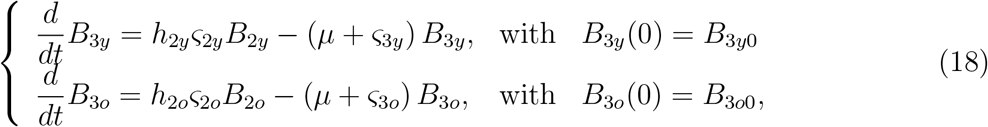

with the total number of occupied beds being *B* = *B*_1_ + *B*_2_ + *B*_3_. The number of deaths caused by severe CoViD-19 cases can be calculated from hospitalized cases. The number of deaths of inpatients is Π_1_ = Π_1_*_y_* + Π_1_*_o_*, where

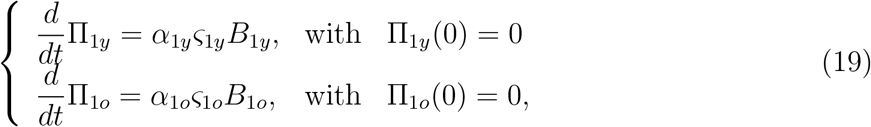

the number of ICU care persons is Π_2_ = Π_2_*_y_* + Π_2_*_o_*, where

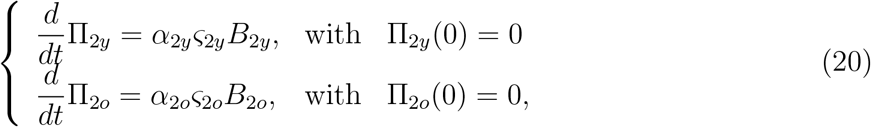

and the number of ICU/intubated persons is Π_3_ = Π_3_*_y_* + Π_3_*_o_*, where

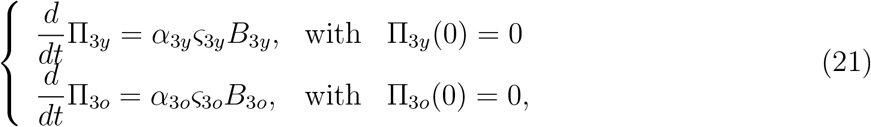

with the total number of deaths being Π*_s_* = Π_1_ + Π_2_ + Π_3_. The total number of persons being cured is *C* = *C_y_* + *C_o_*, where

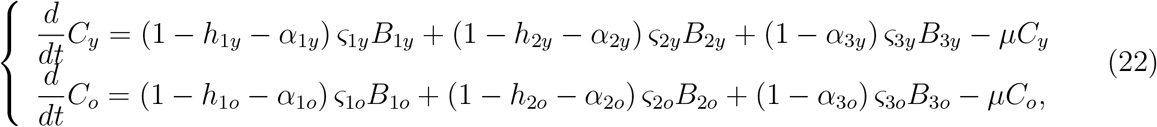

with *C_y_*(0) = *C_y_*(0) = 0.

With respect to the parameters of *B*, Π and *C*, for *j* = *y*, *o*, a proportion *h_j_* of severe CiViD-19 cases are hospitalized and enter into the class of bed *B*_1_, from which a proportion *h*_1_*_j_* needs ICU care (class of bed *B*_2_), and among them a proportion *h*_2_, needs ICU/intubated care (class of bed *B*_3_). The average occupying time of beds by inpatient, ICU and ICU/intubated persons are, respectively, 1/_ζ1_*_j_*-, 1/_ζ2_*_j_* and 1/_ζ3_*_j_*, where _ζ1_*_j_*, _ζ2_*_j_*, and _ζ3_*_j_*, are the discharging rates from hospital, ICU and ICU/intubated care; and *α*_1_*_j_*, *α*_2_*_j_*, and *α*_3_*_j_*, are the additional mortality (fatality) proportions among inpatient, ICU and ICU/intubated persons. The fraction 1 − *h_j_* is possibly non-hospitalization of severe CoViD-19, and 1 − *h*_1_*_j_* − *α*_1_*_j_*, 1 − *h*_2_*_j_* − *α*_2_*_j_*, and 1 − *h*_3_*_j_*, are the proportions of cure of inpatient, ICU and ICU/intubated persons.

As we have pointed out about additional mortality rates, in the estimation of fatality proportions, we must consider that the time of registering new cases and deaths is delayed by Δ_2_ days, that is, Π_1_*_y_* (*t* + Δ_2_) = *α*_1_*_y_*ζ_1_*_y_B*_1_*_y_*(*t*), for instance. The same argument is valid for the number of cured persons, that is, *C_y_*(*t* + Δ_3_) corresponds to *B*_1_*_y_*(*t*) occurred Δ_3_ days ago, for instance.

Table 3 summarizes parameters related to hospitalization and values (for elder classes, values are between parentheses), see Appendix B.

The system of equations (3), (4) and (6) is non-autonomous due to the additional mortality rates. Nevertheless, the fractions of persons in each compartment approach the steady state (see Appendix A), allowing the derivation of the basic reproduction number *R*_0_. Hence, at *t* = 0, the basic reproduction number *R*_0_ is obtained substituting 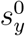 and 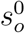 by *N*_0_*_y_*/*N*_0_ and *N*_0_*_o_*/*N*_0_ in equation (A.4), resulting in

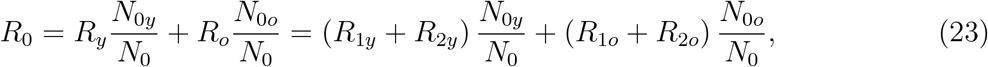

where *R_y_* and *R_o_* are given by equation (A.9), and *R*_1_*_y_*, *R*_2_*_y_*, *R*_1_*_o_* and *R*_2_*_o_* are given by equation (A.8).

**Table 3:** Summary of the parameters related to hospitalization (*j* = *y*, *o*) and values (rates in *days*^−1^ and proportions are dimensionless). Some values are assumed (*) or estimated (**).

| Symbol | Meaning | Value |
| --- | --- | --- |
| $\alpha_{1y} (\alpha_{1o})$ | Fatality proportion of inpatients | 0.01(0.05)** |
| $\alpha_{2y} (\alpha_{2o})$ | Fatality proportion of persons in ICU | 0.05(0.15)** |
| $\alpha_{3y} (\alpha_{3o})$ | Fatality proportion of persons in ICU/intubated | 0.15(0.35)** |
| $\varsigma_{1y} (\varsigma_{1o})$ | Discharge rate of inpatients from hospital | 1/5(1/6)* |
| $\varsigma_{2y} (\varsigma_{2o})$ | Discharge rate from ICU | 1/7 (1/8)* |
| $\varsigma_{3y} (\varsigma_{3o})$ | Discharge rate from ICU/intubated | 1/15 (1/20)* |
| $h_y (h_o)$ | Proportion needing long-stay in hospital | 1.0 (1.0)* |
| $h_{1y} (h_{1o})$ | Proportion needing ICU | 0.15 (0.25)** |
| $h_{2y} (h_{2o})$ | Proportion of ICU needing ICU/intubated | 0.2(0.4)** |

Let us use the approximated effective reproduction number *R_ef_* given by equation (A.11), that is,

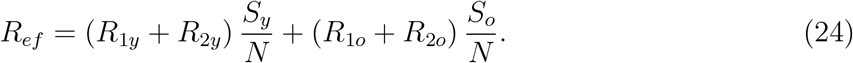

For *t* > 0, we have the effective reproduction number *R_ef_*, with *R_ef_*(0) = *R*_0_ at *t* = 0, which decreases as susceptible persons decrease. However, at *t* = *τ^is^* a pulse in isolation is introduced, hence we have *R_ef_* (*τ^is^*^+^) = *R_r_*, where the reduced reproduction number *R_r_* is given by

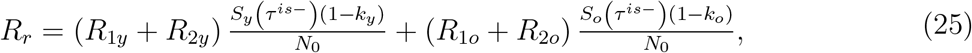

with *S_y_* (*τ^is^*^−^) and *S_o_* (*τ^is^*^−^) being the numbers of susceptible young and elder persons at the time just before the introduction of isolation. Notice that at *t* = *τ^is^*, *R_ef_*(*τ^is^*^−^) jumps down to *Ref*(*τ^is^*^+^). At *i*-th release time *t_i_*, we have *R_ef_*(*τ^is+^*) = *R_u_*(*i*), with the up (increased) reproduction number *R_u_*(*i*) being given by

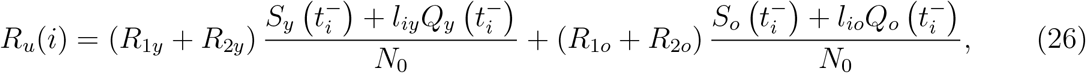

where 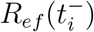 jumps up to 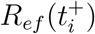. After *t > t_m_*, there is not release anymore, and *R_ef_* = 1 when *t* → ∞, and the new coronavirus returns to the original dynamics driven by *R*_0_.

The protective measures are incorporated in the modelling by the factor *ε*. At the time of introducing these measures, the effective reproduction number *R_ef_* jumps down to *R_p_* due to decrease in the transmission rates.

Given *N* and *R*_0_, let us evaluate the size of the population to trigger and maintain epidemics. Letting *R_ef_* = 1 and assuming that *R_y_* = *R_o_*, the critical size of population *N^th^* is

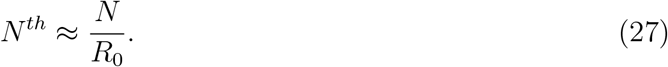

If *N* > *N^th^*, epidemic occurs and persists (*R*_0_ > 1, and epidemic is driven toward the non-trivial equilibrium point *P*\*), and the fraction of susceptible individuals is *s*\* = 1/*R*_0_, where 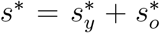; but if *N* < *N^th^*, the epidemics occurs but fades out (*R*_0_ < 1, and epidemics is driven toward the trivial equilibrium point *P*^0^), and the fractions of susceptible individuals *s_y_* and so at equilibrium are given by equation (A.4).

## 3 Results

The results presented in the foregoing section are applied to describe the new coronavirus infection in São Paulo State, Brazil. The first confirmed case of CoViD-19 occurred on 26 February 2020, and the first death on 16 March. On 24 March, the São Paulo State authorities implemented the isolation of population acting in non-essential activities until 31 May.

In this section, we present the parameters estimation, the description of current epidemiological status, and scenarios of release beginning on 1 June.

### 3.1 Parameters estimation

This section, using data collected in São Paulo State from 26 February until 7 May (see Appendix B), we fit transmission (*β_y_* and *β_o_*) and fatality (*α_y_* and *α_o_*) rates, the proportions in the isolated populations (*k_y_* and *k_o_*), and reduction in the transmission rates due to protective measures adopted by population (*ε*). Instead of using equation (B.2) in Appendix B (the least square estimation method), we vary the parameters and choose better fitting by evaluating the sum of squared distances between the curve and data.

#### 3.1.1 Fitting the transmission rates

To fit the transmission rates, we assume that all rates in young persons are equal, as well as in elder persons, that is, we assume that

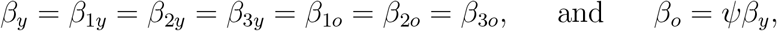

hence, the forces of infection are *λ_y_* = (*A_y_* + *D*_1_*_y_* + *z_y_Q*_2_*_y_* + *A_o_* + *D*_1_*_o_* + *z_o_Q*_2_*_o_*) *β_y_/N* and *λ_o_* = *ψλ_y_*, where *ε_ij_* = 1, with *i* = 1, 2, 3, and *j* = *y,o* (there are not any interventions in the beginning of epidemics). The reason to include factor *ψ* is the reduced capacity of defense mechanism by elder persons (physical barrier, innate and adaptive immune responses, etc.). The force of infection takes into account all virus released by infectious individuals (*A_y_*, *D*_1_*_y_*, *Q*_2_*_y_*, *A_o_*, *D*_1_*_o_* and *Q*_2_*_o_*), the rate of encounter with susceptible persons, and the capacity to infect them (see [21] [22]). Additionally, the amount inhaled by susceptible persons can be determinant in the chance of infection and in the prognosis of CoViD-19 [9].

We use the accumulated new cases (Figure B.1(b)) to estimate the transmission rates *β_y_* and *β_o_*, and the system of equations (3), (4) and (6), with initial conditions given by equation (B.1) in Appendix B, is evaluated to calculate

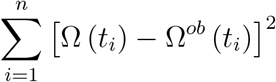

by varying *β_y_* and *β_o_*, where the number of accumulated CoViD-19 cases Ω is given by equation (13). The effects of isolation begun on 24 March are expected to appear later, around 2 April (the sum of incubation and pre-diseased infection periods (see Table 2) is 9.8 days). Hence, we estimate the transmission rates taking into account the confirmed cases from 26 February (*t*_1_) to 5 April (*t*_40_), totalizing *n* = 40 observations. Letting additional mortality rates equal to zero (*α_y_* = *α_o_* = 0), we estimate *β_y_* and *β_o_* = *ψβ_y_* using data collected in São Paulo State. The estimated values are Ψ*_y_* = 0.78 and *β_o_* = 0.90 (both in *days*^−1^), where *ψ* = 1.15, resulting in the basic reproduction number *R*_0_ = 9.24 (partials *R*_0_*_y_* = 7.73 and *R*_0_*_o_* = 1.51), according to equation (23).

Figure 2(a) shows the estimated curve of Ω and observed data, plus two curves with lower transmission rates: *β_y_* = 0.59 and *β_o_* = 0.68 (both in *days*^−1^), with *R*_0_ = 6.99 (partials *R*_0_*_y_* = 5.84 and *R*_0_*_o_* = 1.16); and *R*_0_*_y_* = 0.43 and *β_o_* = 0.50 (both in *days^−^*^1^), with *R*_0_ = 5.09 (partials *R*_0_*_y_* = 4.26 and *R*_0_*_o_* = 0.84). Figure 2(b) shows the extended curves of Ω, which approach asymptotes (or plateaus) indicating the end of the first wave of epidemics. For *R*_0_ = 9.24, 6.99 and 5.09, the curves Ω reach values on 13 September, respectively, 946400, 945700, and 941500. For *R*_0_ = 9.24, the curves for young (Ω*_y_*), elder (Ω*_o_*) and total (Ω) persons approach plateaus (figure not shown) on 13 September with values, respectively, 605300, 341100, and 946400.

We stress the fact that, if the observed data are fitted without caution about interventions, someone could estimate the basic reproduction number to be *R*_0_ = 5.09 or less (near the horizontal axis, the observed data seem to approach the curve of *R*_0_ = 5.09). The basic reproduction number estimated by our model is *R*_0_ = 9.24. However, if we estimate the basic reproduction number using SIR model with different infective persons at t = 0, we obtain *R*_0_ = 3.22 (for *I*(0) = 10), *R*_0_ = 2.66 (for *I*(0) = 25) and *R*_0_ = 2.38 (for *I*(0) = 50), with other initial conditions being *S*(0) = 44.6 million, and *R*(0) = 0.

Let us estimate the critical size of the population (all persons are susceptible) *N^th^* from equation (27). For *R*_0_ = 9.24, we have *N^th^* = 4.83 million. Hence, for São Paulo State, the isolation of 39.8 million (89%) persons or above is necessary to avoid the persistence of epidemics. The number of young persons is 1.97 million less than the threshold number of isolated persons to guarantee the eradication of CoViD-19.

#### 3.1.2 Fitting the additional mortality rates

We estimate taking into account confirmed deaths from 26 February (*t*_1_) to 31 March (*t*_35_), totalizing *n* = 35 observations, remembering that the first death occurred on 16 March.

**Figure 2:**
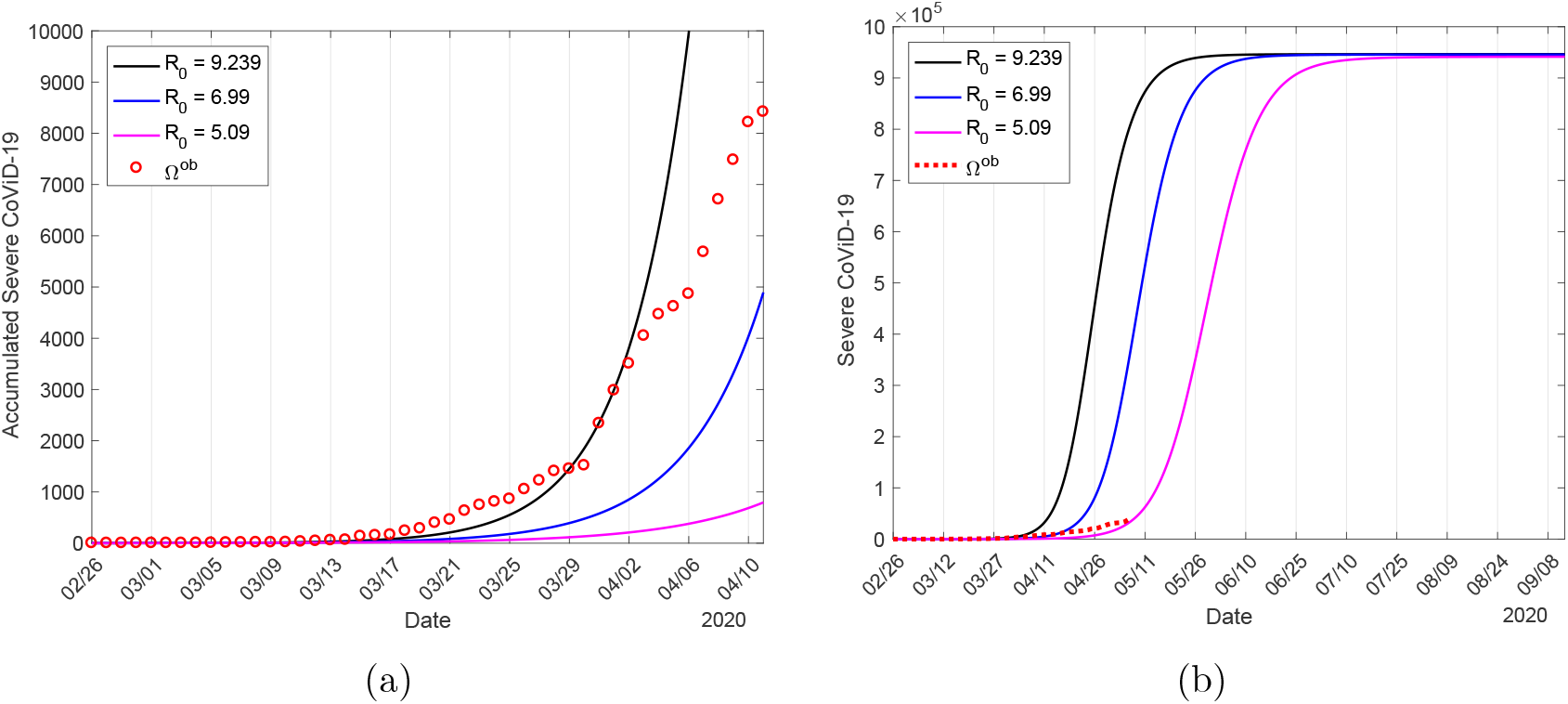
The estimated curve of Ω and observed data, plus two curves with lower transmission rates: *β_y_* = 0.59 and *β_o_* = 0.68 (*days^−^*^1^), with *R*_0_ = 6.99, and *β_y_* = 0.43 and *β_o_* = 0.50 (*days^−^*^1^), with *R*_0_ = 5.09 (a); and extended curves of accumulated number of severe CoViD-19 Ω (b).

To estimate the mortality rates *α_y_* and *α_o_*, we fix the previously estimated transmission rates *β_y_* = 0.78 and *β_o_* = 0.90 (both in *days^−^*^1^), resulting in *R*_0_ = 9.24, and evaluate the system of equations (3), (4) and (6), with initial conditions given by equation (B.1) in Appendix B, to calculate, by varying *α_y_* and *α_o_*,

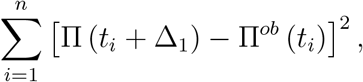

where Π is given by equation (14), with Π(0) = 0, and the time of death registration *t_i_* portraits the deaths of new cases Δ_1_ times ago, that is, *D*_2_(*t_i_ −* Δ_1_). We minimize the sum of differences to estimate the additional mortality rates *α_y_* and *α_o_* letting *α_y_* = 0.1*α_o_*, due to the fact that the lethality among young persons is lower than elder persons [6], and by varying Δ_1_ and *α_o_*. The estimated delay is Δ_1_ = 9 *days*, and the additional mortality rates are *α_y_* = 0.00053 and *α_o_* = 0.0053 (both in *days^−^*^1^).

Figure 3 shows the estimated curve of Π and the observed death data (a), and the extended curves of the number of CoViD-19 deaths (Π*_y_*, Π*_o_*, and Π = Π*_y_* + Π*_o_*) from equation (14) (b). The estimated curves Π*_y_*, Π*_o_*, and Π = Π*_y_* + Π*_o_* reach plateaus, and on 13 September the values are 3820 (0.6%), 34110 (10%) and 37930 (4.0%), respectively, for, young, elder and total persons. The percentage between parentheses is the ratio Π/Ω, Ω being given in Figure 2(b).

#### 3.1.3 Estimating the proportion of persons in isolation

We fix the transmission rates *β_y_* = 0.78 and *β_o_* = 0.90 (both in *days^−^*^1^), giving *R*_0_ = 9.24, and the additional mortality rates *α_y_* = 0.00053 and *α_o_* = 0.0053 (both in *days^−^*^1^), to estimate isolation (described by proportions *k_y_* and *k_o_*) of susceptible persons aiming the control of infection. Isolation was introduced on 24 March, and we will estimate taking into account the confirmed cases until 21 April, totalizing 29 observations. The mean proportion of persons in isolation from 24 March to 3 May is *k_mean_* = 0.53 (see Figure B.1).

**Figure 3:**
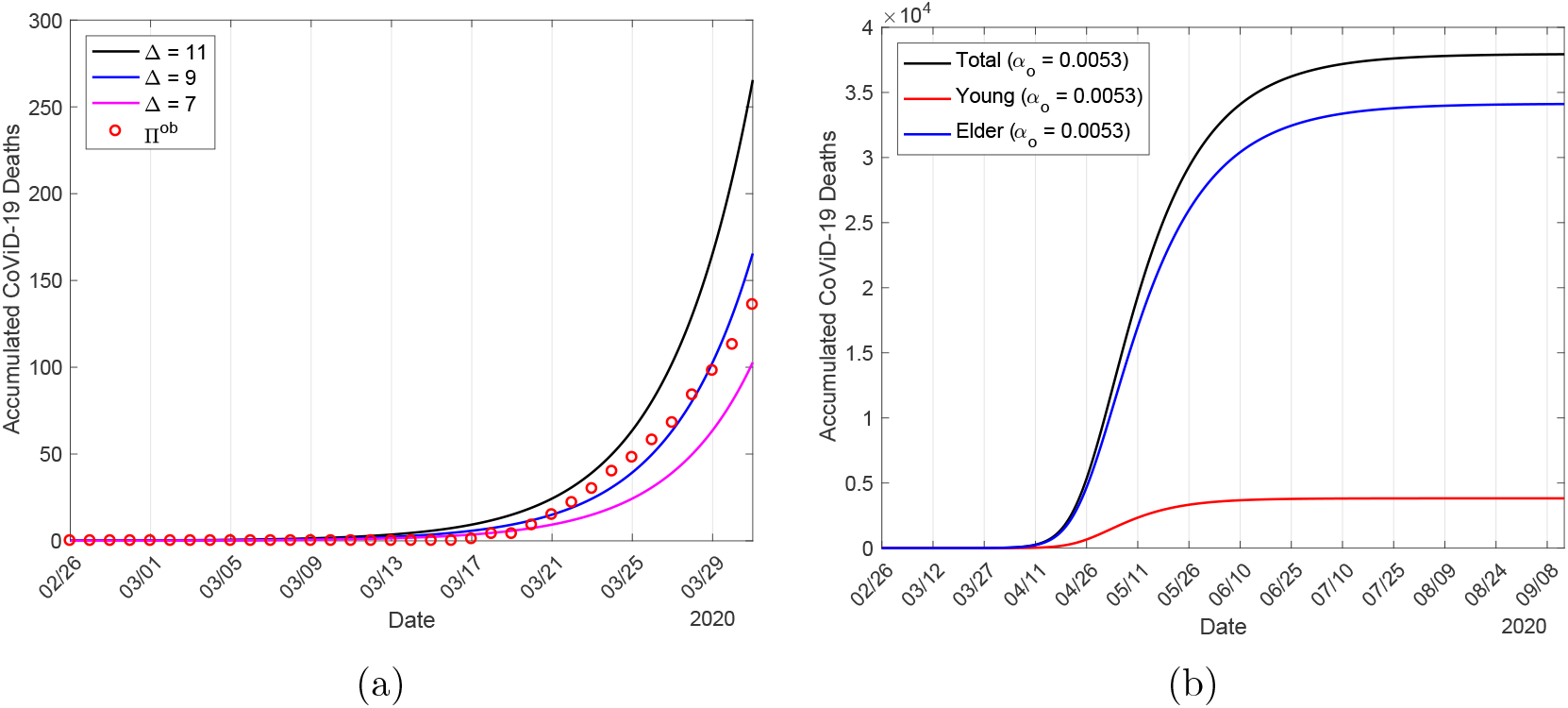
The estimated curve of Π and the observed data for *R*_0_ = 9.24 (a), and the accumulated number of CoViD-19 deaths (Π*_y_*, Π*_o_*, and Π = Π*_y_* + Π*_o_*) (b).

Here, we estimate the isolation parameter by varying *k_y_* and *k_o_*. The system of equations (3), (4) and (6) is evaluated with initial conditions (on 26 February) given by equation (B.1) in Appendix B, and boundary conditions (on 24 March) given by equations (8) and (9), and we calculate the sum of square differences

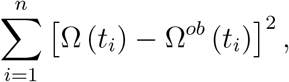

where *t*_1_ is 24 March and *t*_29_ is 21 April. We assume *k* = *k_y_* = *k_o_* and varied *k* = 0, 0.4, 0.53, 0.6, 0.7, and 0.8, from which we observe that *k* = 0.4 and 0.6 fit part of the observed data, while *k* = 0.7 does not. Hence, we chose *k* = *k_mean_* = 0.53 as the better estimated value. Figure 4(a) shows the curves of Ω for different proportions in isolation of São Paulo State and CoViD-19 observed data, and 4(b), the extended curves.

As we have pointed out in the description of the data (see Appendix B), the proportion in isolation delayed in approximately 9 days indeed affected the daily incidence of CoViD-19. Moreover, the estimated proportion agrees with the average proportion in isolation in São Paulo State. To incorporate the trend indicated by the data, we estimated taking into account the data from 24 March to 21 April. From Figure 4(b), Ω approaches plateau, and the asymptotic values on 13 September for *k* = 0, 0.4, 0.528, 0.6, 0.7, and 0.8 are, respectively, 964600, 567600 (60%), 444000 (47%), 372500 (40%), 268200 (28%) and 146000 (15%). The percentage between parentheses is the ratio Ω(*k*)/Ω(0).

Figure 5 shows the estimated curves of Ω*_y_*, Ω*_o_*, and Ω = Ω*_y_* + Ω*_o_* (a) and Π*_y_*, Π*_o_*, and Π = Π*_y_* + Π*_o_* (b), for *k* = 0.53. On 13 September, the curves of accumulated cases and deaths of CoViD-19 approach plateaus, attaining for Ω*_y_*, Ω*_o_*, and Ω the values, respectively, 283600 (47%), 160300 (47%) and 444000 (47%); and Π*_y_*, Π*_o_* and Π attain, respectively, 1791 (0.6%), 16010 (10%) and 17800 (4%). The percentages between parentheses are the ratios Ω(*k_mean_*)/Ω(0) and Π/Ω.

**Figure 4:**
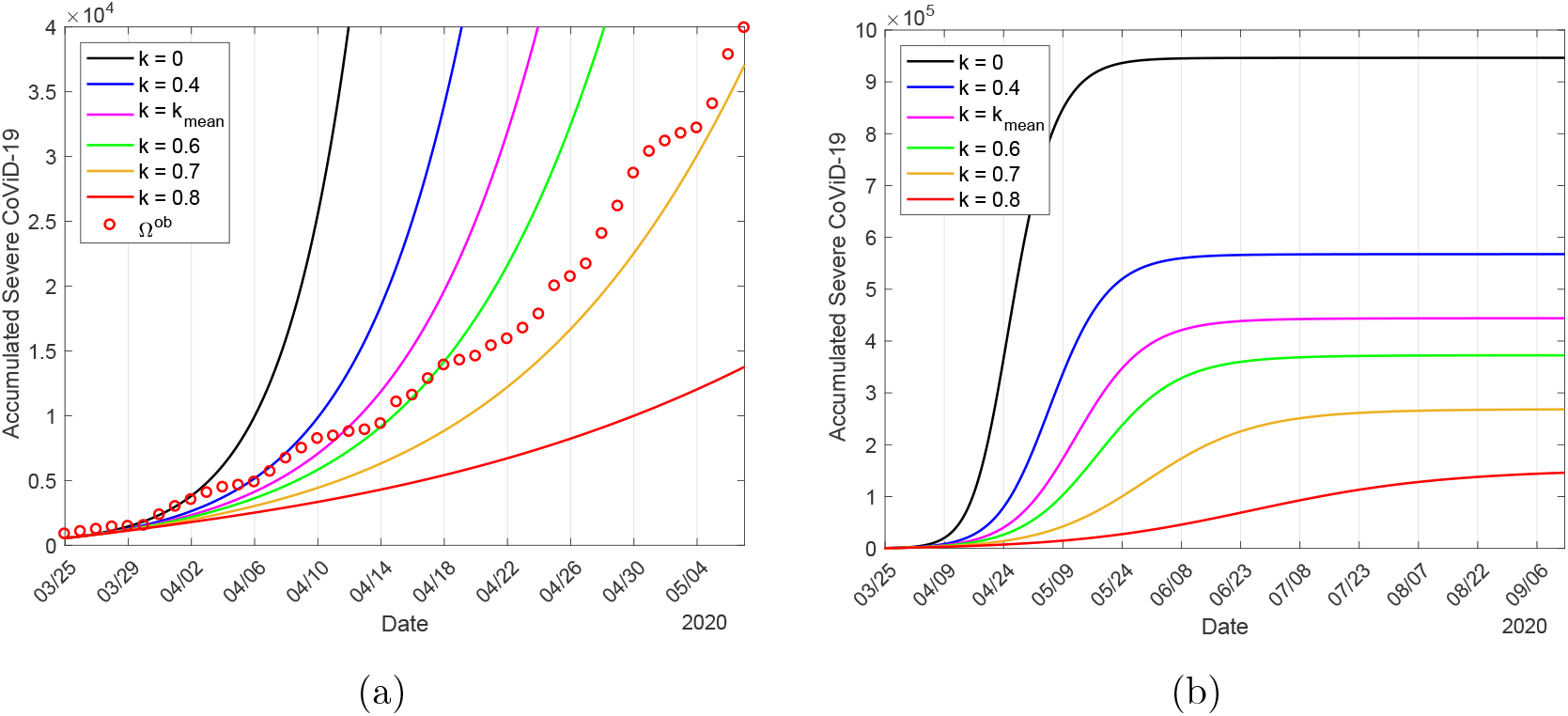
The proportions in isolation in São Paulo State with CoViD-19 observed data (a), and the extended curves (b).

**Figure 5:**
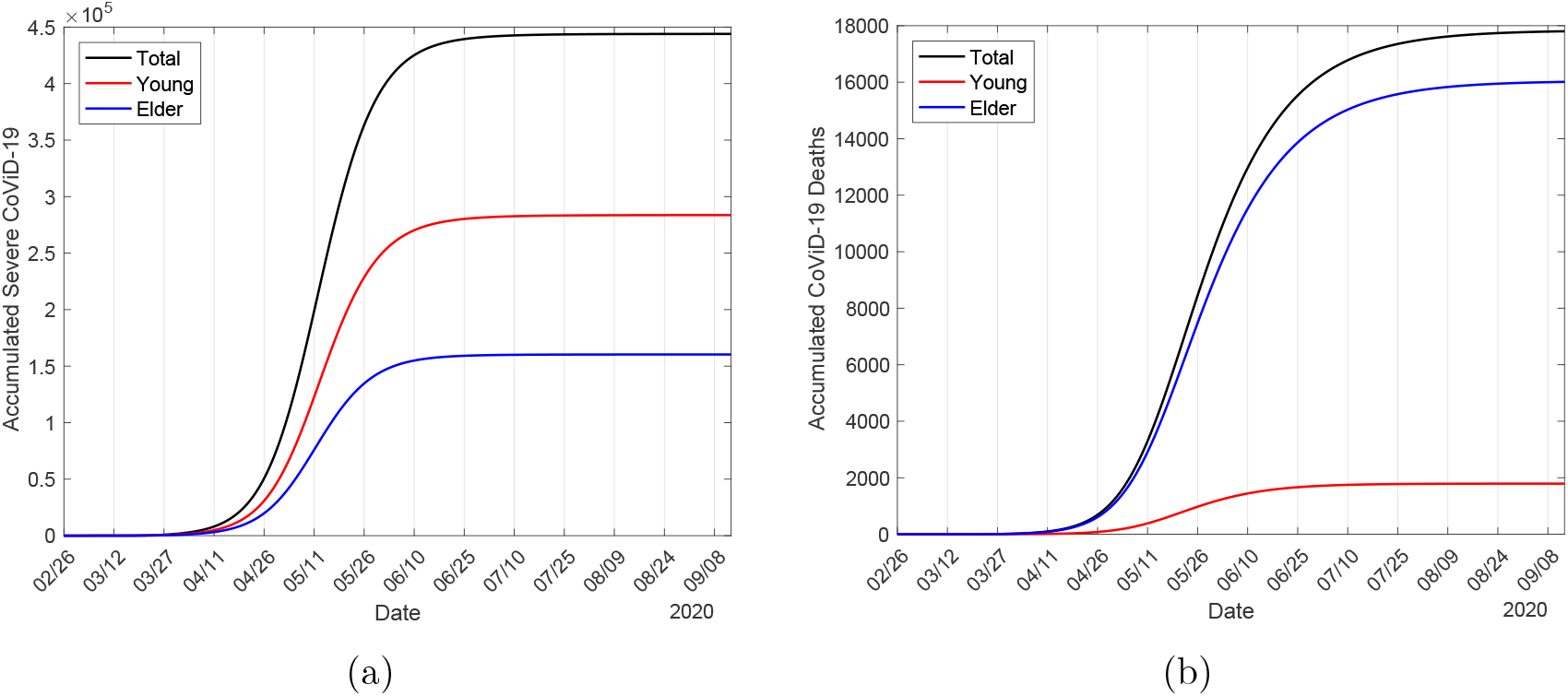
The estimated curves of Ω*_y_*, Ω*_o_*, and Ω = Ω*_y_* + Ω*_o_* (a), and Π*_y_*, Π*_o_*, and Π = Π*_y_* + Π*_o_* (b), for *k* = 0.53.

#### 3.1.4 Estimating the reduction in the transmission rates

On 13 April, 20 days after the beginning of isolation, we observe the first point leaving completely the curve. This new trend can not be explained by an increased proportion in isolation (see Figure 4). To take into account this new tendency of data, in [32] we hypothesized that the using of face mask, constant hygiene (washing hands with alcohol and gel, and protection of mouth, nose and eyes, etc.), social distancing and other protective measures may decrease the transmission of infection. Based on the literature [10], we include these protective measures in the model and we estimate the reduction in the transmission rates.

We fix the transmission rates *β_y_* = 0.78 and *β_o_* = 0.90 (both in *days*^−1^), giving *R*_0_ = 9.24, the additional mortality rates *α_y_* = 0.00053 and *α_o_* = 0.0053 (both in *days*^−1^), and the proportion in isolation of susceptible persons *k* = 0.53 to estimate the protective factor ε.

Let us assume that on 4 April (9 days before the observed points leaving consistently the estimated curve with isolation alone), 11 days after the beginning of isolation, protective measures were adopted by persons, which reduced the transmission rates from *β_y_* and *β_o_* to 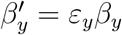 and 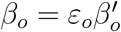, see equation (1). The system of equations (3), (4) and (6) is evaluated with the initial conditions (26 February) given by equation (B.1) in Appendix B, and boundary conditions (24 March) given by equations (8) and (9), and we calculate the sum of square differences

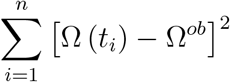

by varying *ε* = *ε_y_* = *ε_o_* between 4 April (*t*_1_) and 7 May (*t*34), totalizing 34 observations. We considered *ε* = 0.8, 0.7, 0.6, 0.5 and 0.4, and the better estimated value is *ε* = 0.5. Figure 6(a) shows the curves of Ω and observed data, where 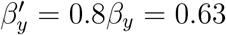, 0.7*β_y_* = 0.55, 0.6*β_y_* = 0.47, 0.5*β_y_* = 0.39 and 0.4*β_y_* = 0.31 (all in *days*^−1^), plus *ε* =1 and without isolation (*k* = 0). Figure 6(b) shows the extended curves of Q for 4 different decreasing values of transmission rates plus *ε* = 1.

**Figure 6:**
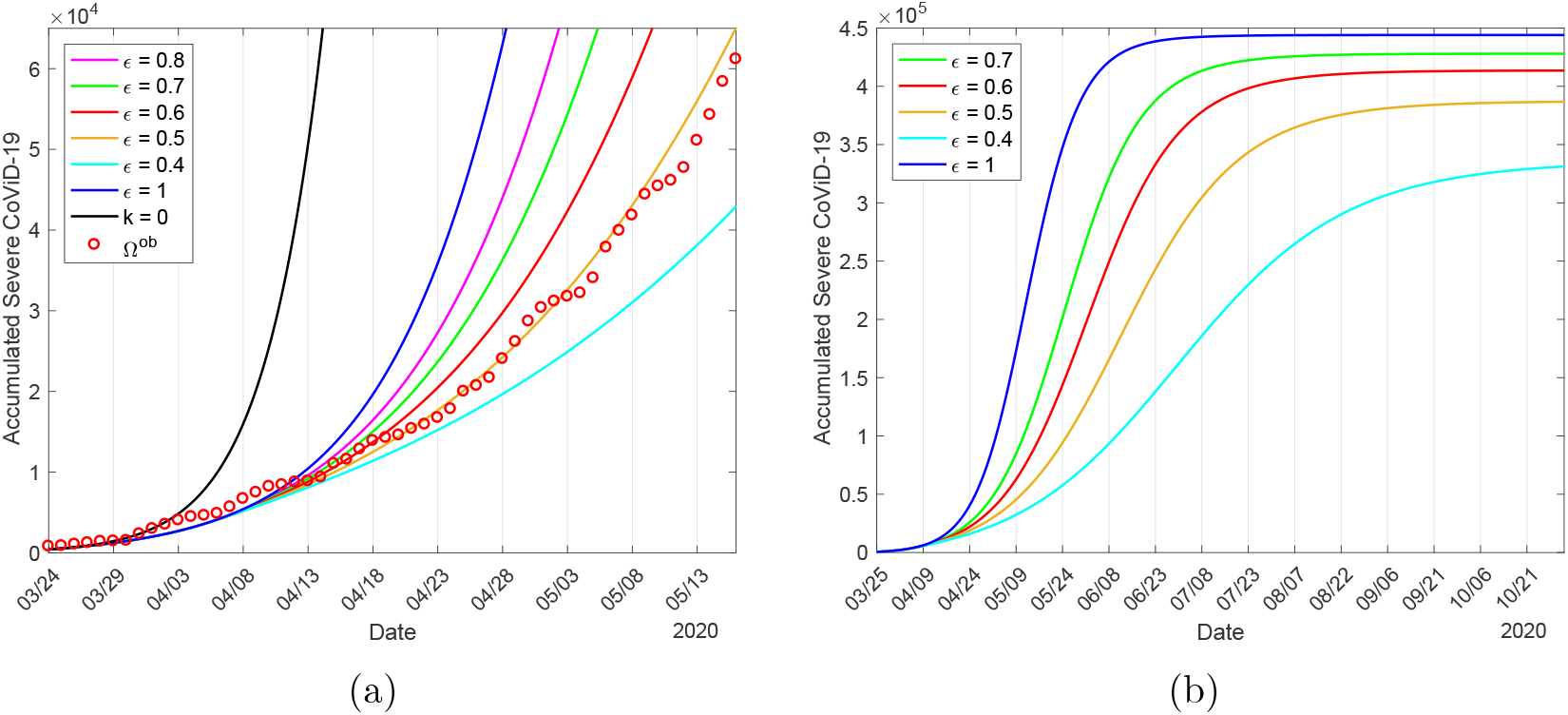
The curves of Ω = Ω*_y_*+Ω*_o_* and observed data, where 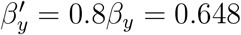, 0.7*β_y_* = 0.57, 0.6*β_y_* = 0.49, 0.5*β_y_* = 0.41 and 0.4*β_y_* = 0.32, plus 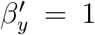 and without isolation (a), and extended curves of Ω for 4 different decreasing values of transmission rates plus 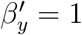 (b).

From Figure 6(b), Ω approaches plateau, and the asymptotic values on 2 November for *ε* = 1, 0.7, 0.6, 0.5 and 0.4 are, respectively, 444000, 427900 (97%), 413500 (93%), 386700 (87%) and 331200 (75%). The percentage between parentheses is the ratio Ω(*ε*)/Ω(0).

Figure 7 shows the estimated curves of Ω*_y_*, Ω*_o_*, and Ω = Ω*_y_* + Ω*_o_* (a) and Π*_y_*, Π*_o_*, and Π = Π*_y_* + Π*_o_* (b), for *k* = 0.53 and *ε* = 0.5, that is, 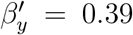 and 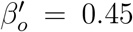 (both in *days*^−1^), reducing the basic reproduction number to *R*_0_ = 4.62. On 2 November, the curves of accumulated cases and deaths of CoViD-19 approach plateaus, with Ω*_y_*, Ω*_o_*, and Ω attaining asymptotic values, respectively, 243800 (86%), 142800 (89%) and 386700 (87%); and Π*_y_*, Π*_o_* and Π attain, respectively, 1540 (0.6%), 14240 (10%) and 15780 (4%). The percentages between parentheses are the ratios Ω (*ε* = 0.5) /Ω(*ε* = 1) and Π/Ω.

**Figure 7:**
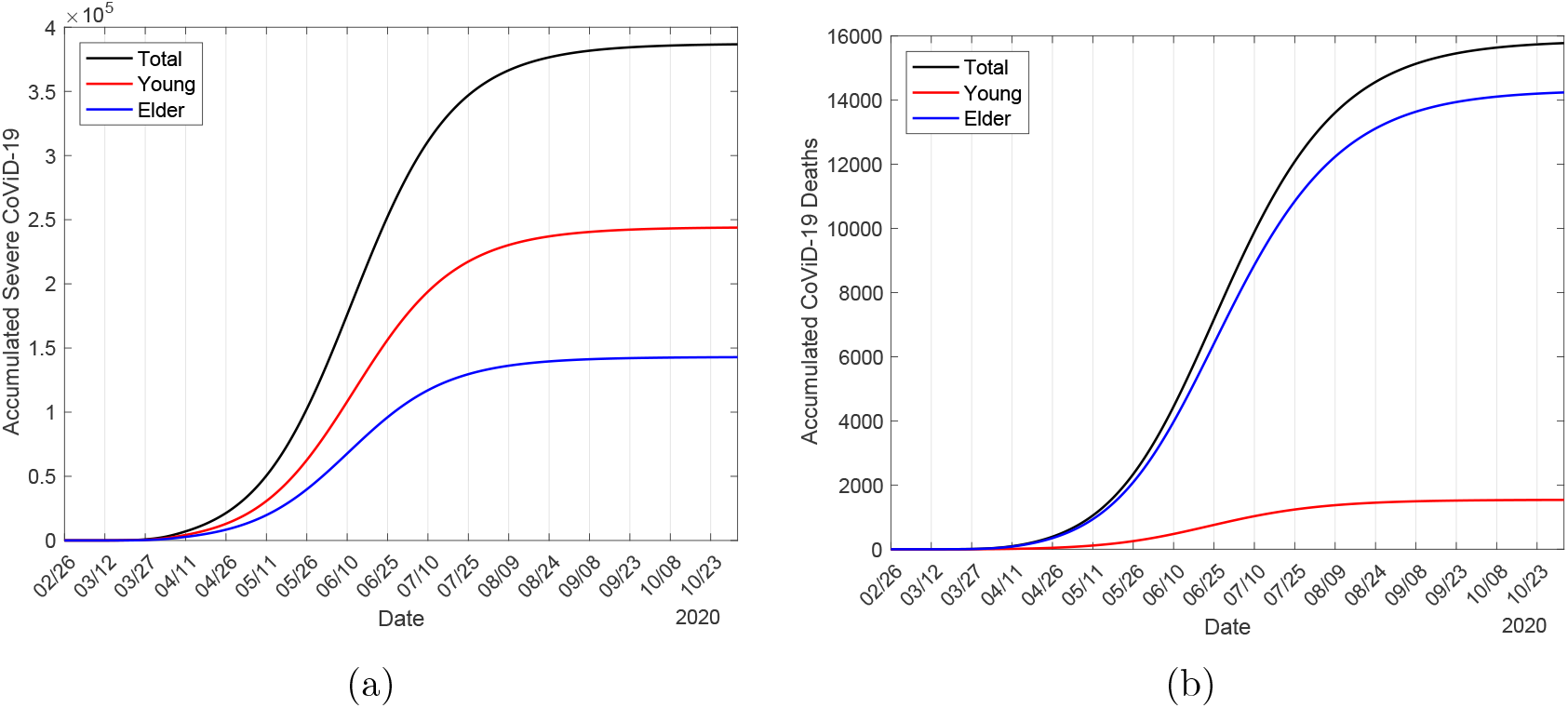
The estimated curves of Ω*_y_*, Ω*_o_*, and Ω = Ω*_y_* + Ω*_o_* (a) and Π*_y_*, Π*_o_*, and Π = Π*_y_* + Π*_o_* (b), for *k* = 0.53 and *ε* = 0.6. The reduced transmission rates are 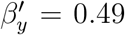 and 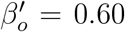, with *R*_0_ = 5.76.

### 3.2 Current epidemiological scenario

Before presenting epidemiological scenarios considering releasing strategies, we present the current epidemiological status using all previously estimated parameters: The transmission rates *β_y_* = 0.78 and *β_o_* = 0.90 (both in *days^−^*^1^), giving *R*_0_ = 9.24; the additional mortality rates *α_y_* = 0.00053 and *α_o_* = 0.0053 (both in *days^−^*^1^); the proportion in isolation of susceptible persons *k* = 0.53; and the protective factor ε = 0.5 reducing the transmission rates to 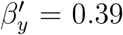 and 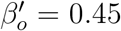 (both in *days*^−1^), giving *R*_0_ = 4.62. Hereafter, all these values are fixed, unless explicitly cited.

In Figure 8, we show the effects of interventions on the dynamics of the new coronavirus. As interventions are added (isolation followed by protective measures), we observe decreasing in the peaks of severe CoViD-19 *D*_2_, which move to the right. Figure 8(a) shows the curves representing *k* = 0 and *ε* = 1 (without interventions), *k* = 0.53 and *ε* = 1 (only isolation), and *k* = 0.53 and *ε* = 0.5 (isolation and protective measures). In Figure 8(b), we show the number of immune persons *I* corresponding to the three cases shown in Figure 8(a). The curves follow a sigmoid-shape.

In the absence of interventions (*k* = 0 and *ε* = 1), the numbers of immune persons *I_y_*, *I_o_*, and I increase from zero to, respectively, 35.3 million, 6.1 million and 41.4 million on 1 June (figure not shown). When interventions are adopted, the numbers are, on June 1, 3.7 million (10%), 700000 (11%) and 4.4 million (11%). Figure 8(b) shows only *I* with and without interventions. The percentage between parentheses is the ratio between with and without interventions *I*(*k*, *ε*)/*I*(0, 1) on 1 June (the isolation will end on 31 May).

**Figure 8:**
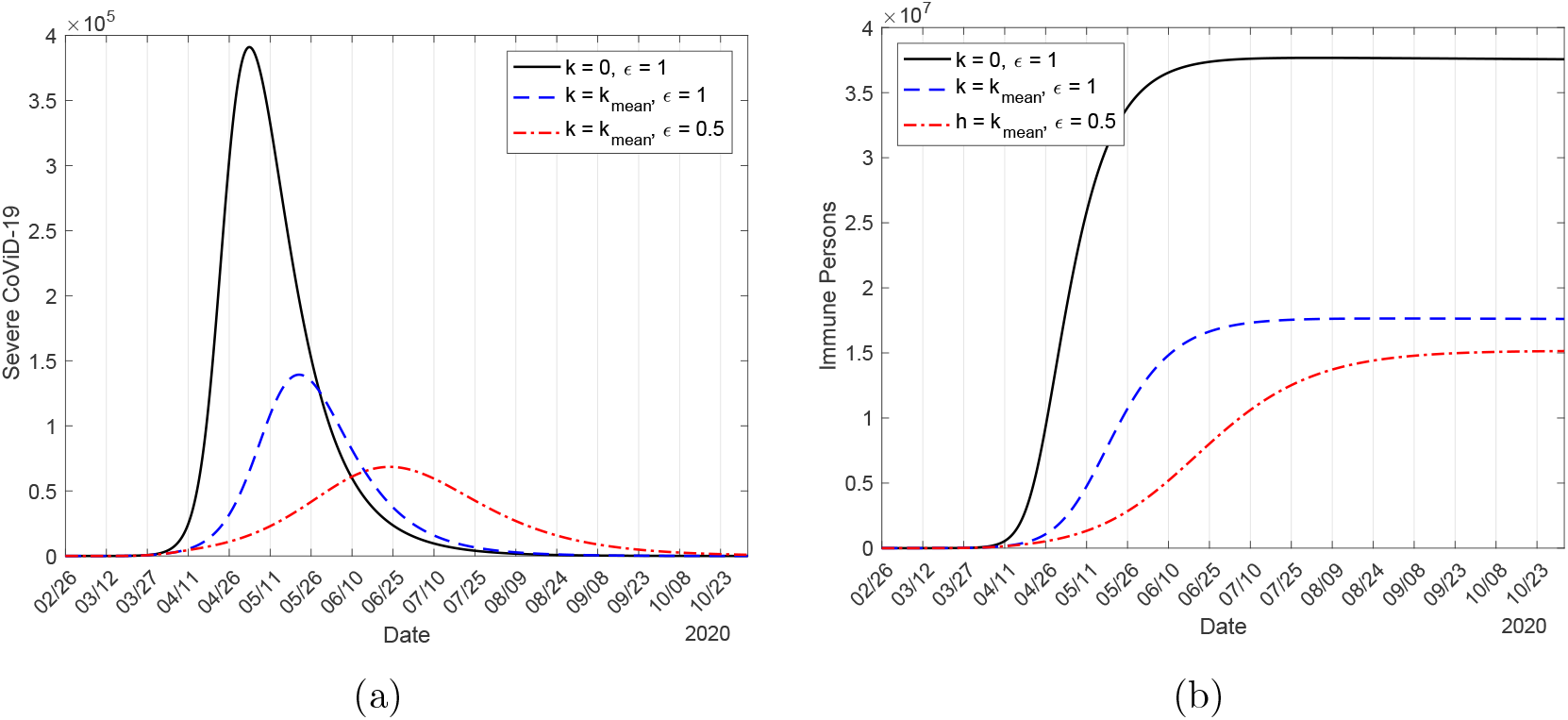
The effects of intervention on the dynamics of new coronavirus from without interventions (*k* = 0 and *ε* = 1), only isolation (*k* = 0.53 and *ε* =1), and isolation plus protection (*k* = 0.53 and *ε* = 0.6) (a), and number of immune persons corresponding to three cases.

Let us compare the peaks of *D*_2_. The peaks for young, elder and total persons without any interventions (*k* = 0 and *ε* =1) are, respectively, 226600, 164800, and 391000, occurring on 2, 4 and 3 May. Considering isolation alone (*k* = 0, 53 and *ε* =1), the peaks for young, elder and total persons are, respectively, 78190 (35%), 61230 (37%), and 139300 (36%), which occur on 21, 23 and 22 May. Considering interventions (*k* = 0, 53 and *ε* = 0.5), the peaks for young, elder and total persons are, respectively, 36510 (16%), 32000 (19%), and 68460 (18%), which occur on 23, 25 and 23 June. The percentage between parentheses is the ratio between with and without interventions *D*_2_(*k*,*ε*)/*D*_2_(0,1).

The isolation does not change the basic reproduction number, and just after the end of isolation, the dynamics is driven by the original epidemics, reaching the non-trivial equilibrium point *P*^*^ given by *R*_0_. However, the reduction in the transmission rates decreases the basic reproduction number. Hence, from 26 February to 3 April, the dynamics system is driven by *β_y_* = 0.78 and *β_o_* = 0.90 (both in *days*^−1^, *R*_0_ = 9.24), and since 4 April, the dynamics is driven by 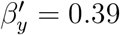 and 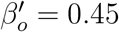 (both in *days*^−1^, *R*_0_ = 4.62). If the protective measures are abandoned, the dynamics is driven by the trend of *R*_0_ = 9.24 again.

Due to isolation and protective measures, so many people remain as susceptible. In Figure 9 we show circulating susceptible persons *S_y_*, *S_o_* and *S* = *S_y_* + *S_o_* (a), and circulating plus isolated susceptible persons 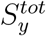, 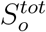 and 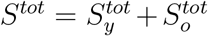 (b), using equation (12). Remember that 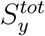 differs from *S_y_* just after the introduction of isolation (24 March).

Let us compare the number of susceptible persons before and after the introduction of interventions, which will end on 31 May. On 1 June, the numbers of susceptible persons *S_y_*, *S_o_* and *S* = *S_y_* + *S_o_* without any interventions (*k* = 0 and *ε* =1) are, respectively, 25260, 320 and 25580, and with interventions, the numbers of susceptible persons are 11.04 million (43705%), 1.85 million (590095%) and 12.9 million (50391%), for young, elder and total persons, respectively. On 1 June, for 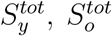, and 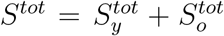, we have, respectively, 30.9 million (122328%), 5.4 million (1735783%) and 36.3 million (142025%). The percentage between parentheses is the ratio between with and without interventions *S*(*k*, *ε*)/*S*(0, 1) on 1 June. With interventions, at the end of isolation, there are more than 437-times the number of susceptible young and total persons, and for elder persons, 5900-times with respect to without interventions. However, if we add those in isolation to be released on 1 June, there are more than 1220-times the number of susceptible young and total persons, and for elder persons, 17357-times. Hence, if all persons are released without planning, the second wave will be intense, infecting much more elder persons. In the absence of vaccine and effective treatment, interventions aiming reduction in transmission must be continued for a long time to avoid rebounding of epidemics or a second wave.

**Figure 9:**
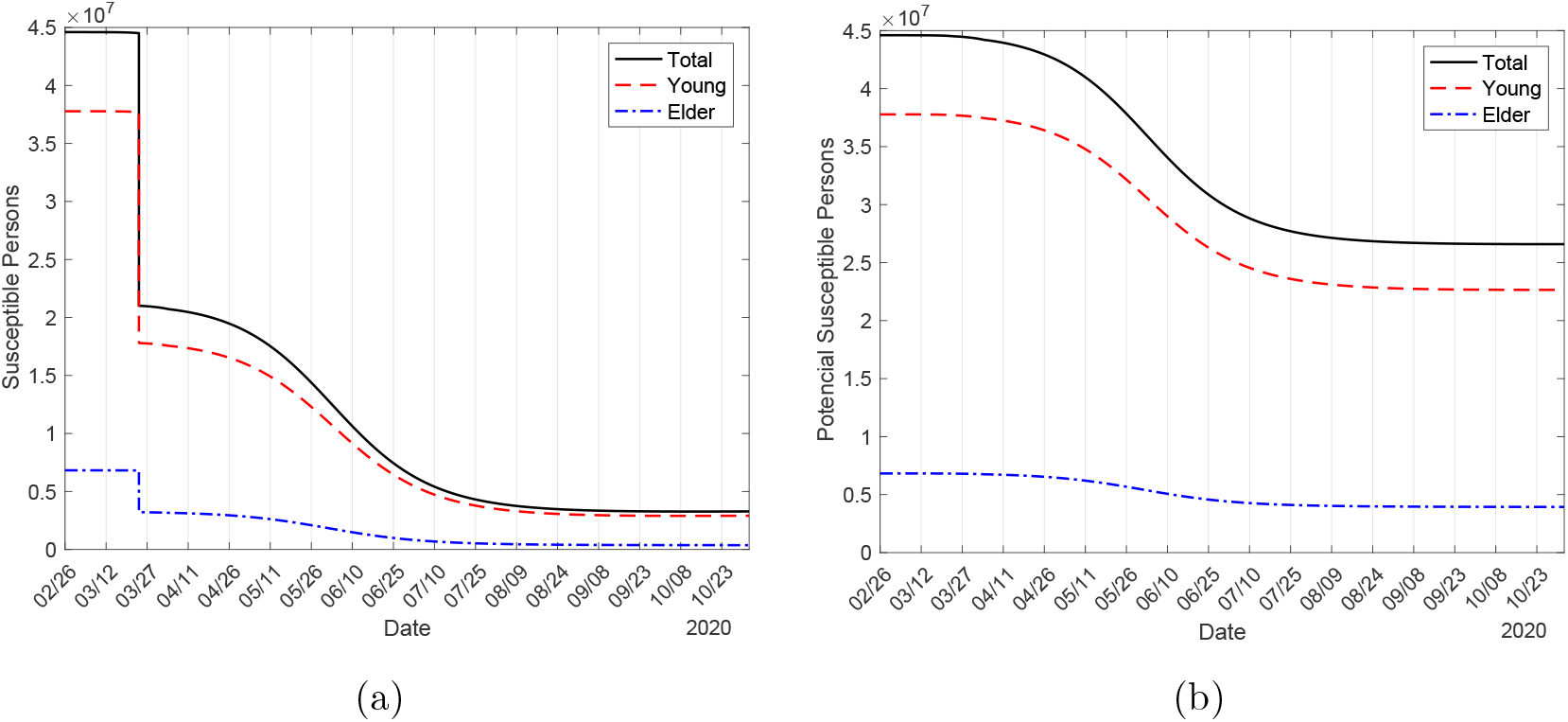
The circulating susceptible persons *S_y_*, *S_o_* and *S = S_y_* + *S_o_* (a), and potential susceptible persons to be infected 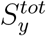, 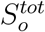 and 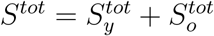 (b).

It is important to estimate the basic reproduction number, which portraits the beginning and ending phases of an epidemics [30]. During the evolving of epidemics, however, the effective reproduction number determines the intensity of epidemics. We use the approximate effective reproduction number *R_ef_*, given by equation (24), to follow the trend of dynamics, remembering that *R_ef_* > 1 implies epidemics in expansion, while *R_ef_* < 1, in contraction. Figure 10 illustrates the effective reproduction number *R_ef_* with (a) and without (b) interventions during the evolving of epidemics represented by *D*_2_. To be fitted together in the same frame with *R_ef_*, the curve of *D*_2_ is divided by 7000 (a) or 40000 (b). The curve of *R_ef_* follows the shape of susceptible persons as shown in Figure 9, as expected. As expected, *R_ef_* = 1 occurs before the peak of epidemics.

When epidemics evolves following its natural way (without interventions), on 1 June, the effective reproduction number is very low (*R_ef_* = 0.005) in the descending phase of epidemics, but epidemic is maintained due to the high number of infectious individuals infecting many susceptible persons. However, with interventions, on 1 June we have *R_ef_* = 1.33 in the ascending phase, but from Figure 10(a), *R_ef_* = 1 occurs just before the peak of epidemics. Hence, further release must take this fact into account and public health authorities must not assume that all descending phase is relatively safe - when *R_ef_* is lower but near than 1, there is a great possibility of rebounding of epidemics when release is initiated.

**Figure 10:**
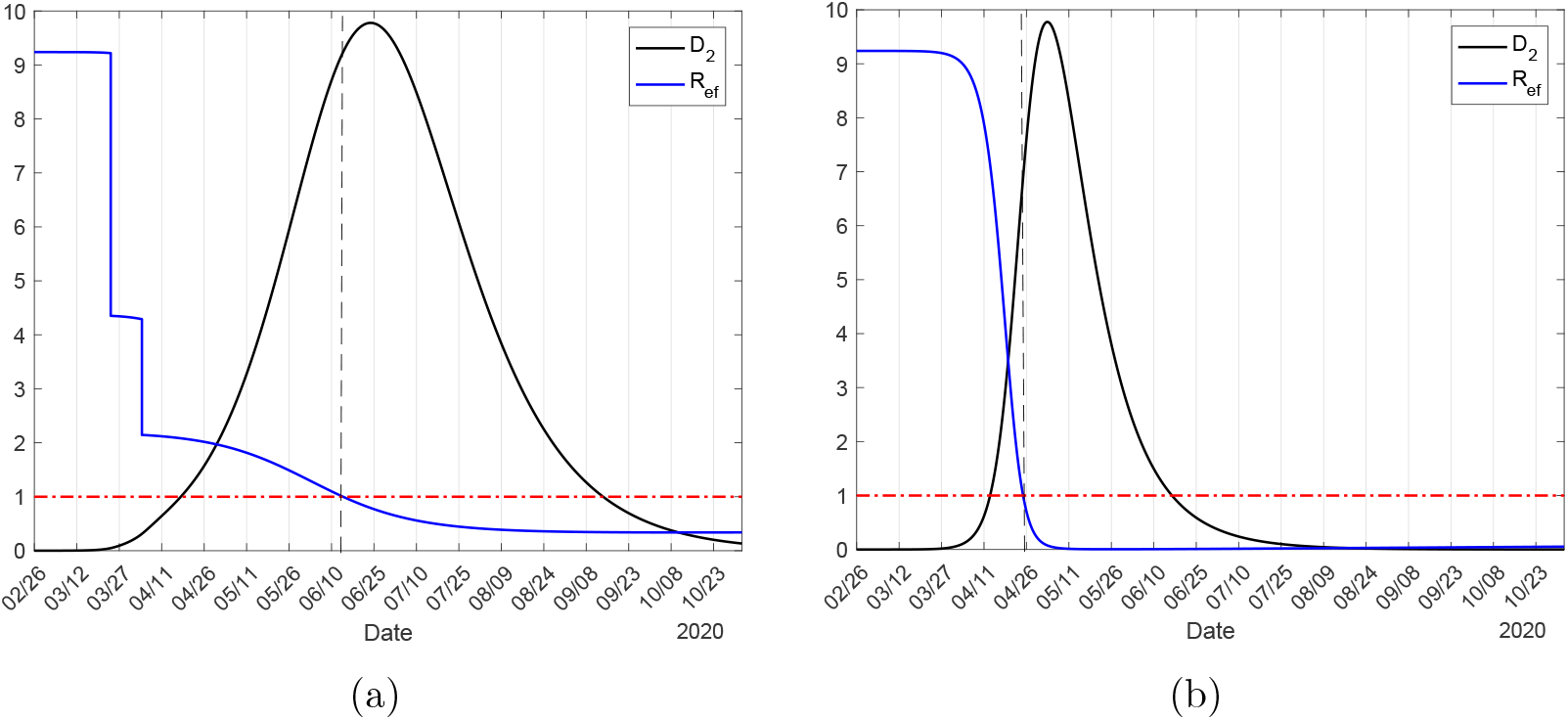
Illustration of the effective reproduction number *R_ef_* when isolation and protection are adopted (a) and without adoption (b), with the actual value of curve *D*_2_ must be multiplied by, respectively, 7000 (a) and 40000 (b).

#### 3.2.1 Calculating daily CoViD-19 from accumulated cases

We used the accumulated data shown in Figure B.1(b) in Appendix B and Ω given by equation (13) to estimate the transmission rates *β_y_* and *β_o_*, the proportion in isolation k, and the protective factor e. The curve labelled *ε* = 0.5 in Figure 6(b) is the estimated curve Ω, from which the curve of severe cases *D*_2_ was derived, corresponding to the most flattened curve shown in Figure 8(a). From the estimated curve of Ω we derive the daily cases Ω*_d_* calculated by equation (15). In Figure 11(a) we show the calculated curve Ω*_d_* and daily cases presented in Figure B.1(a). In Figure 11(b) we show the initial part of the estimated curve Ω with observed data Ω*^ob^*, the extended Ω*_d_* and daily observed cases 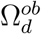, and severe cases *D*_2_. The peaks of *D*_2_ and Ω_d_ occur, respectively, on 23 and 12 June.

On 12 June, the peak of the daily cases of CoViD-19 predicted by the estimated values of parameters reaches 5287, remembering that the peak of *D*2 on 23 June is 68460. However, on 23 June, the number of accumulated cases is 243000, which is 355% of the peak of *D*_2_, and 63% of cases when the first wave of epidemics ends (386700).

**Figure 11:**
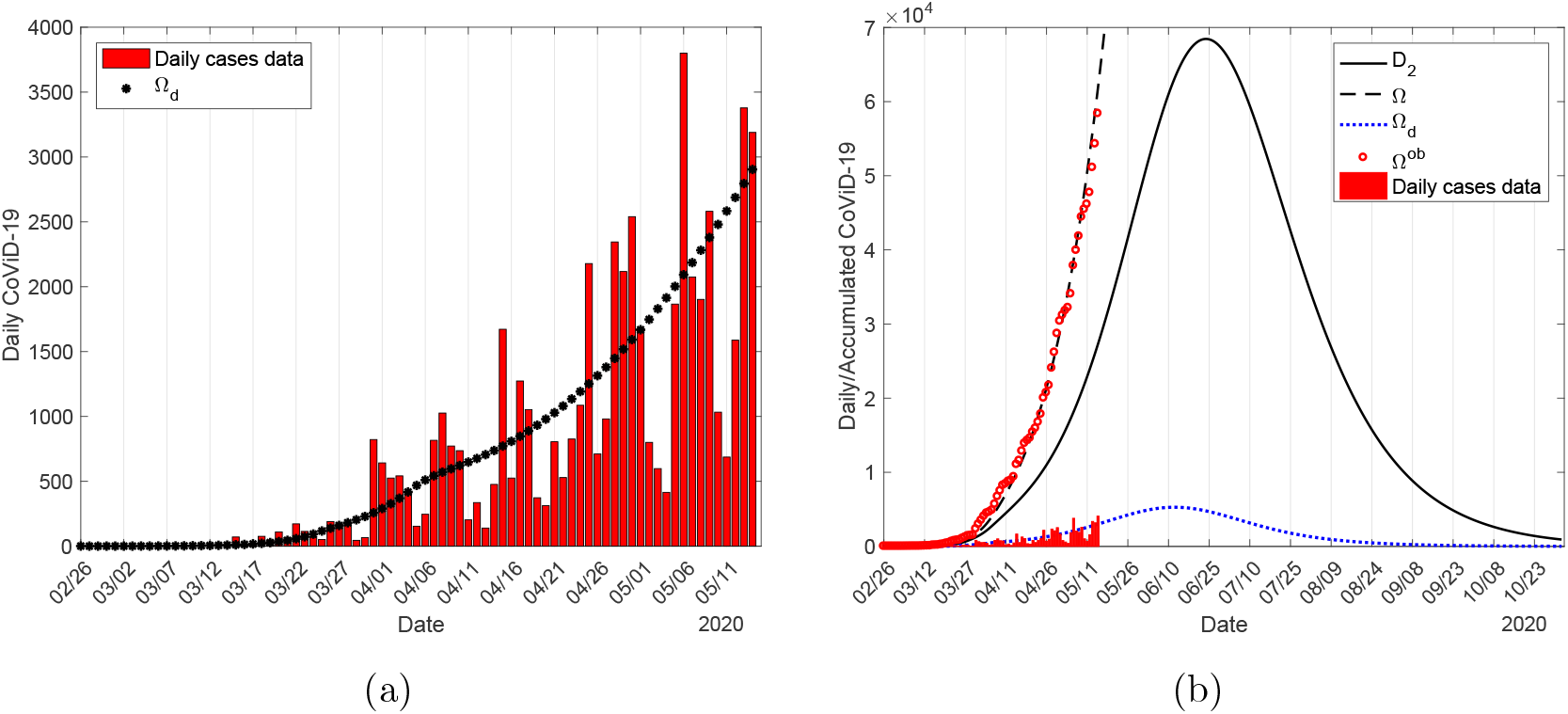
The fitted curve of Ω*_d_* with accumulated cases data (a), and the extended curve with previously adjusted *D*_2_ (b).

#### 3.2.2 Subnotification along epidemics

Let us discriminate the circulation of the new coronavirus in a community according to the infected classes. Figure 12 shows all persons harbouring this virus (*E_j_*, *A_j_*, *D*_1_*_j_*, *Q*_2_*_j_* and *D*_2_*_j_*), for young (*j* = *y*) (a) and elder (*j* = *o*) (b) persons. Notice that *Q*_1_*_j_* = *Q*_3_*_j_* = 0 (mass testing is not available yet).

**Figure 12:**
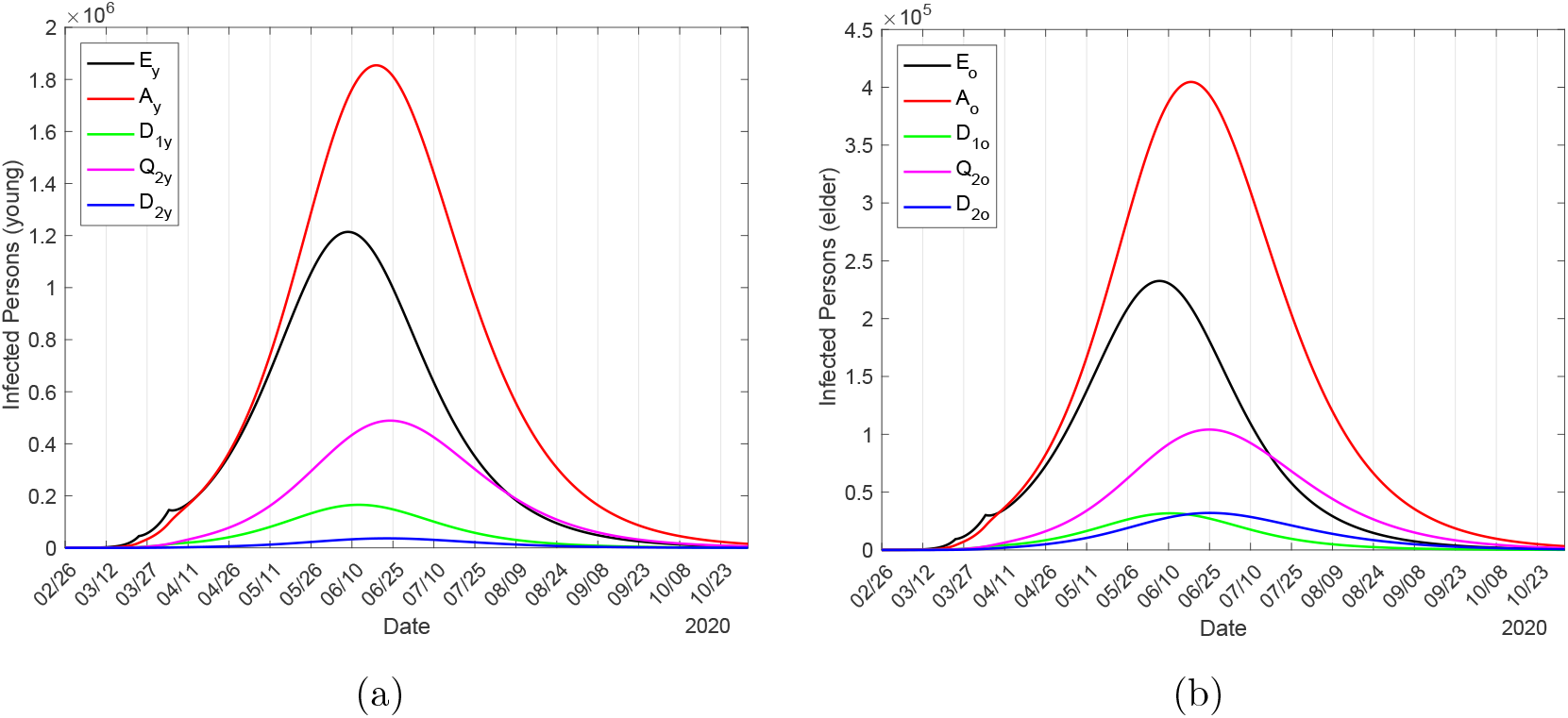
All persons harbouring the new coronavirus (*E_j_*, *A_j_*, *D*_1_*_j_*, *Q*_2_*_j_* and *D*_2_*_j_*), for young (*j* = *y*) (a) and elder (*j* = *o*) (b) persons.

Let us assess the subnotification of CoViD-19, considering the ratio hidden:apparent. We classify all those who harbour the new coronavirus (exposed, asymptomatic and not manifesting) in the hidden category, and in the apparent category, all those who manifest symptoms.

Hence, the ratio is calculated as (*E_j_* + *A_j_* + *D*_1_*_j_*)/(*Q*_2_*_j_* + *D*_2_*_j_*). In Figure 13(a) we show the ratio hidden:apparent based on Figure 12 for young (*j* = *y*), elder (*j* = *o*) and total persons. On 26 February (*t* = 0), the ratio was 10: 1 for young and elder persons due to initial conditions. Aiming comparison, Figure 13(b) shows the ratio hidden:apparent in the absence of interventions.

**Figure 13:**
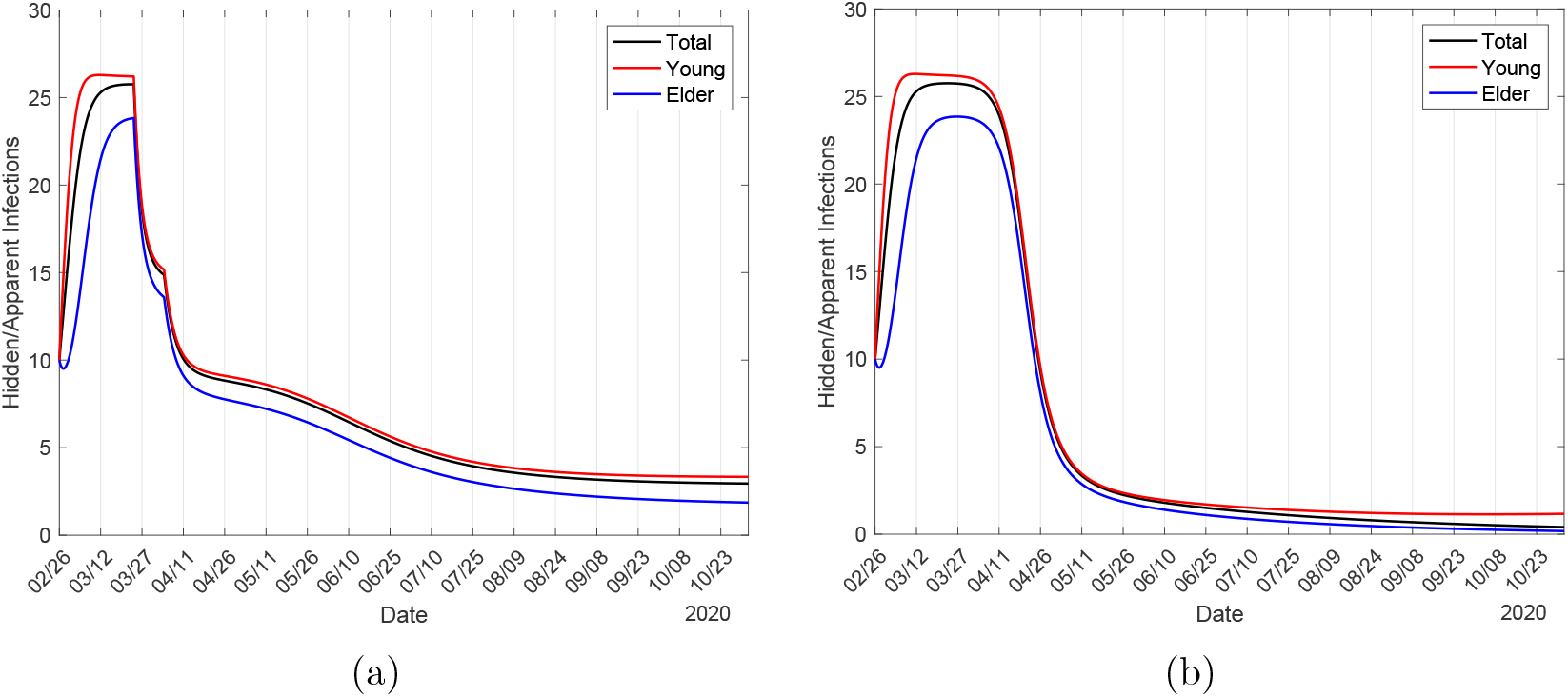
The ratio hidden:apparent based on Figure 12, calculated based on the ratio (*E* + *A* + *D*_1_): (*Q*_2_ + *D*_2_) (a). The ratio hidden:apparent when there is not any intervention is shown in (b).

Figure 13(b) portraits the tendency of subnotification understood as the ratio hidden:apparent in the absence of interventions. There is a quick increasing following the ascending phase of epidemics, reaching a plateau for a while, and then decreases quickly, reaching an asymptote. When interventions are introduced, subnotification changes behaviour following the sequential introduction of interventions: There is a first perturbation, which occurs at the moment when isolation is introduced, followed by a second perturbation, when protective measures are adopted by the circulating population. Comparing Figures 12 and 13(a), as the epidemic evolves, the ratio increases quickly in the beginning, reaches a plateau during the increasing phase, and decreases during the declining phase, finally reaching another plateau at the ending phase of the first wave. In the first plateau, the ratios are 24: 1, 27: 1, and 26: 1 for, respectively, elder, young, and total persons. At the end of the first wave, subnotification reaches asymptotically the ratio 1: 1 in the absence of interventions (2: 1 with interventions). Therefore, during epidemics, there are much more hidden than apparent persons, which makes any control mechanisms hard if mass testing could not be implemented. However, the estimation of the ratio between hidden and apparent cases can be helpful in designing mass testing aiming to isolate asymptomatic persons. If we calculate (*E_j_* + *A_j_* + *D*_1_*_j_* + *Q*_2_*_j_*)*/D*_2_*_j_*, the ratios in the plateaus are 360: 1, 105: 1, and 250: 1 for, respectively, elder, young, and total persons.

#### 3.2.3 Epidemics in isolated population

We evaluate the spread of the new coronavirus in isolated population, and the effects of lockdown implemented during the evolving of epidemics.

**Isolation – transmission in isolated population** As we have pointed out, when isolation is done during epidemics, in the isolated population harbouring the virus can be found. Just before the beginning of isolation on 24 March, the number of persons in each class, for *k* = 0.53, are

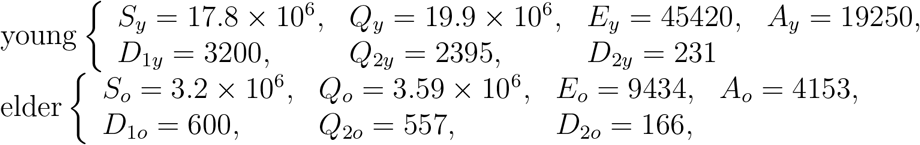

with *Q*_1_*_y_* = *Q*_1_*_o_* = 0 and *I* = 10032.

To estimate the circulation of virus in isolated population, we simulate the system of equations (3), (4) and (6) taking as initial conditions the number of persons in each class: *S_y_* = *Q_y_* = 19.9 million, *S_o_* = *Q_o_* = 3.6 million, *Q_y_* = *Q_o_* = 0, and for all other variables, we assume 53% of the corresponding values just before 24 March. Figure 14 shows the curves of *D*_2_*_y_*, *D*_2_*_o_*, and *D*_2_ = *D*_2_*_y_* + *D*_2_*_o_* for 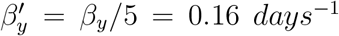, with *R*_0_ = 1.85 (a), and 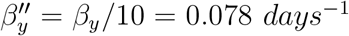, with *R*_0_ = 0.93 (b), from 24 March to 31 May, during the period of isolation. On 1 June, the numbers of severe CoViD-19 cases for *R*_0_ = 1.85 are 4632, 3990 and 8622, respectively, for young, elder, and total persons; and 316, 366 and 642, for *R*_0_ = 0.93. On 1 June, for *k* = 0.53 and *R*_0_ = 0.93, this 0.9% additional CoViD-19 cases compared to the peak of cases among circulating persons (68460) is negligible, however for *R*_0_ = 1.85, the 13% additional CoViD-19 cases is considerable.

**Figure 14:**
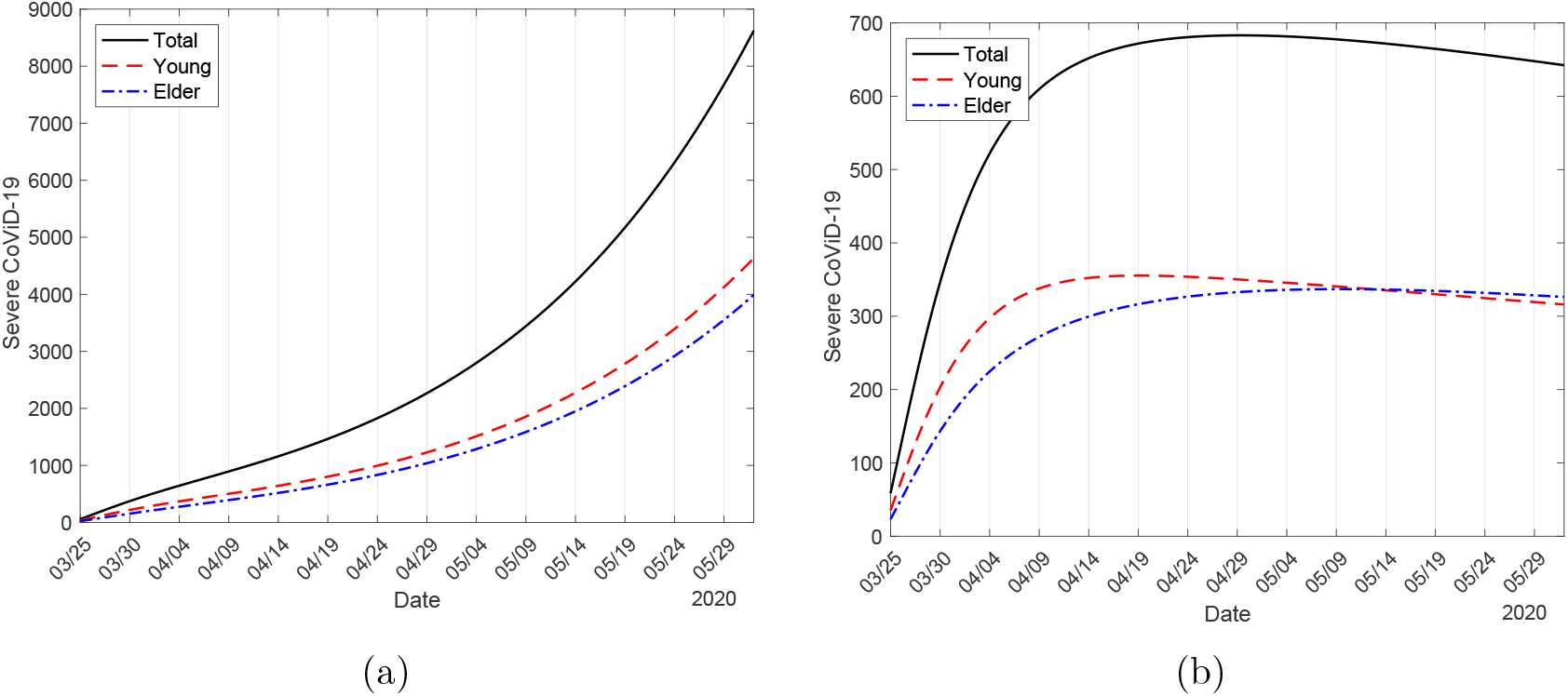
The curves of *D*_2_*_y_*, *D*_2_*_o_*, and *D*_2_ = *D*_2_*_y_* + *D*_2_*_o_* for 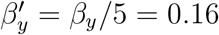, with *R*_0_ = 1.92 (a), and 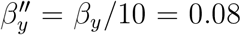, with *R*_0_ = 0.96 (b), from 24 March to 11 May, during the period of isolation.

The virus should be circulating in the isolated population through restricted contact occurring in the household and/or neighborhood. If asymptomatic persons are introduced in the isolated population, epidemics can be triggered if *R*_0_ is higher than 1.

**Lockdown – Transmission due to locked-down asymptomatic persons** As in isolation, when lockdown is implemented during the evolving of epidemics, persons harbouring the virus are gathered to the previously isolated population. Depending on the epidemiological status, a higher number of persons harbouring the virus will be in close contact with elder persons, which constitute a group under higher risk of infection and death. As we have pointed out in [31] [32], the presence of infectious young persons increases greatly the risk of infection in elder persons.

We illustrate the lockdown using epidemiological scenarios of São Paulo State with interventions (isolation *k* = 0.53 and protective factor *ε* = 0.5), and supposing that on 10 May lockdown is implemented. We analyze the lockdown lasting for a few days, for instance, 14 days, and a high proportion in isolation, for instance, *k* = 0.9. On 10 May, just before the beginning of lockdown, the number of circulating persons in each class is

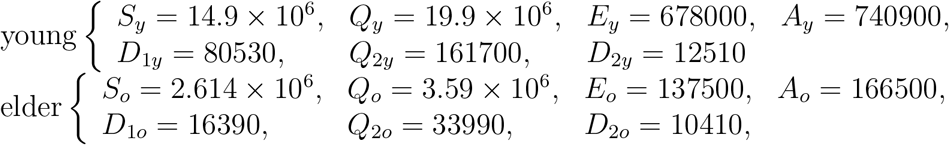

with *Q*_1_*_y_* = *Q*_1_*_o_* = 0 and *I* =1.6 million. However, the isolated susceptible persons on 24 March (*S_y_* = *Q_y_* = 19.9 million, *S_o_* = *Q_o_* = 3.59 million, and zero for all other classes) decreased by natural mortality *μ*, and on 10 May, they are *S_y_* = 19.8 million and *S_o_* = 3.58 million.

Our task is evaluating the transmission in the isolated population, now composed by locked-down on 10 May and previously isolated persons on 24 March, by simulating the system of equations (3), (4) and (6). The boundary conditions on 10 May (denoted by *x*) are given by equation (8) for all classes with *k* = 0.9, recalling that in the isolated population there are only susceptible persons. Hence, the boundary conditions supplied to the system of equations describing the transmission of infection in isolation and lockdown population are

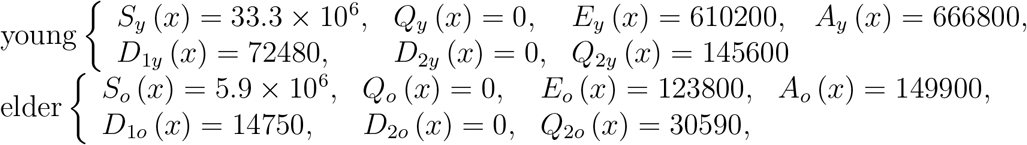

with *Q*_1_*_y_*(*x*) = *Q*_1_*_o_* (*x*) = 0 and *I* (*x*) = 1.42 × 10^6^. Remember that severe CoViD-19 is isolated or hospitalized, hence *D*_2_*_y_* (*x*) = *D*_2_*_o_* (*x*) = 0.

In Figure 15, we illustrate the implementation of 14 days lockdown. We assume that the protective factor is *ε* = 0.5 during isolation and lockdown, but the transmission rates are reduced by a factor 5, that is, 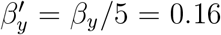 and 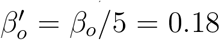 (both in *days*^−1^), with *R*_0_ = 0.93 (a), and reduced by 3, 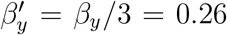 and 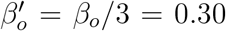 (both in *days*^−1^), with *R*_0_ = 1.54 (b). We present the curves of *D*_2_*_y_*, *D*_2_*_o_*, and *D*_2_ with isolation only (dashed) and with isolation and lockdown (continuous). The values for young, elder, and total persons in isolation alone on 10 May are, respectively, 12080, 10020 and 22120, and on 24 May, 21880, 18350 and 39980. Hence, the differences between 24 May and 10 May are 9800, 8330 and 17860, respectively, for young, elder, and total persons. In population composed by isolation followed by lockdown, on 24 May the number of severe CoViD-19 cases increases from 0 to the values given in Table 4 below.

**Figure 15:**
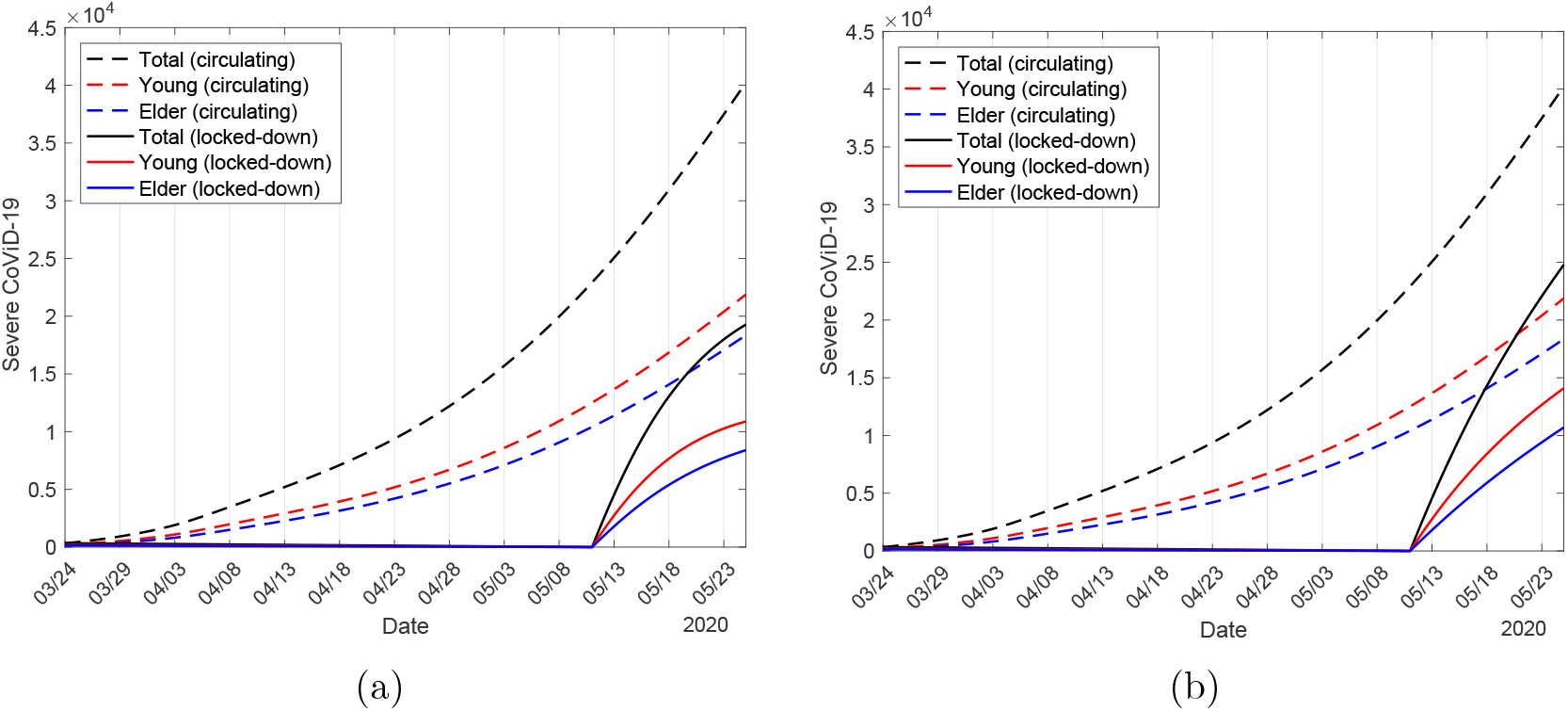
We present the curves of *D*_2_*_y_*, *D*_2_*_o_*, and *D*_2_ considering the protection factor *ε* = 0.5 during isolation and lockdown, and the transmission rates are reduced to 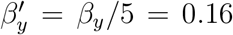 and 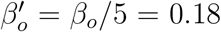, with *R*_0_ = 0.93 (a), and 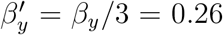 and 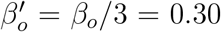, with *R*_0_ = 1.54 (b).

Let us define the reduction in transmission rates in the isolated and subsequent lockdown population. Among them, the transmission rates can be reduced by isolating factor ϖ, that is, they are reduced by 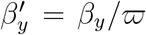 and 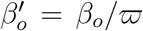, and by the protective factor *ε*, that is, 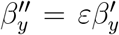 and 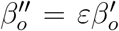, remembering that *β_y_* = 0.78 and *β_o_* = 0.90 (both in *days*^−1^), with *R*_0_ = 9.24. Table 4 shows the values obtained on 24 May for different factors of reduction in the transmission rates. With respect to ϖ =1 and *ε* = 1 (first row), *D*_2_*j, j* = *y*, *o*, and *D*_2_ = *D*_2_*_y_* + *D*_2_*_o_* are the difference between the values on 24 May and 10 May, that is, the number of new cases during this 14 days without lockdown. For other values of ϖ and *ε*, Δ*D*_2_ is the subtraction of new cases among locked-down persons on 24 May and the new cases among circulating persons without lockdown. Hence, − and + mean the advantage or disadvantage of lockdown.

The first row of Table 4 shows the dynamics of circulating persons driven by *R*_0_ = 9.24 from 26 February to 3 April, and by *R*_0_ = 4.62 since 4 April. For all other cases, there is a beginning of the dynamics of CoViD-19 transmission in the isolated population, for this reason *R_ef_* = *R*_0_ on 10 May. Observe that the lockdown lasting for 14 days is advantageous if the values of reduction factors w and e result in *R*_0_ < 1 (from Table 4, *R*_0_ = 0.93 is disadvantageous). We stress that expressive numbers of exposed (*E* = 734000), asymptomatic (*A* = 816700) and pre-diseased (*D*_1_ = 87230) persons are transferred to isolation with lockdown, showing that many people are getting sick in their home or being infected. For instance, from the last row, in the absence of transmission in the isolated population (*R*_0_ = 0), we have 12220 new cases among isolated persons, which is exactly the contribution of already infected persons isolated by lockdown. In this case, the avoiding of 5640 new cases due to lockdown (32% of 17860) is desirable, but may bring troubles about releasing strategies later. However, if there is transmission in the isolated population, for instance the last but one row *R*_0_ = 0.37, the number of new cases is 14870, and the difference 2990 is 17% of 17860, which increases as *R*_0_ increases. If the absence or low intensity of transmission could not be guaranteed, lockdown may bring another trouble besides the increased number of new cases, that is, many elder persons should be infected due to closeness with numerous asymptomatic especially young persons [31].

**Table 4:** The values attained on 24 May for different factors of reduction in the transmission rates: isolating factor ϖ, that is, 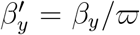 and 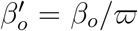, and protection actions factor *ε*, that is, 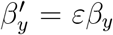 and 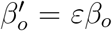, remembering that *β_y_* = 0.78 and *β_o_* = 0.90 (both in *days*^−1^), with *R*_0_ = 9.24.

| $\varpi$ | $\varepsilon$ | $R_0$ | $R_{ef}$ | $D_{2y}$ | $D_{2o}$ | $D_2$ | $\Delta D_{2y}$ | $\Delta D_{2o}$ | $\Delta D_2$ |
| --- | --- | --- | --- | --- | --- | --- | --- | --- | --- |
| 1 | 1 | 4.62 | 1.53 | 9800 | 8330 | 17860 | — | — | — |
| 2 | 0.6 | 2.77 | — | 21780 | 16130 | 37920 | −11980 | −7800 | −20060 |
| 2 | 0.5 | 2.31 | — | 18700 | 13960 | 32660 | −8900 | −5630 | −14800 |
| 2 | 0.4 | 1.85 | — | 15860 | 11950 | 27810 | −6060 | −3620 | −9950 |
| 3 | 0.6 | 1.85 | — | 15860 | 11950 | 27810 | −6060 | −3620 | −9950 |
| 3 | 0.5 | 1.54 | — | 14100 | 10700 | 24790 | −4300 | −2370 | −6930 |
| 3 | 0.4 | 1.23 | — | 12440 | 9510 | 21950 | −2640 | −1180 | −4090 |
| 5 | 0.6 | 1.11 | — | 11800 | 9056 | 20860 | −2000 | −726 | −3000 |
| 5 | 0.5 | 0.92 | — | 10880 | 8394 | 19270 | −1080 | −64 | −1410 |
| 5 | 0.4 | 0.74 | — | 9988 | 7756 | 17740 | −188 | +574 | +120 |
| 10 | 0.6 | 0.55 | — | 9135 | 7142 | 16280 | +665 | +1188 | +1580 |
| 10 | 0.5 | 0.46 | — | 8721 | 6844 | 15560 | +1079 | +1486 | +2300 |
| 10 | 0.4 | 0.37 | — | 8316 | 6844 | 14870 | +1484 | +1779 | +2990 |
| — | 0 | 0 | — | 6777 | 5438 | 12220 | +3023 | +2892 | +5640 |

In Figure 16, we present the new epidemics occurring in initially isolation plus lockdown population due to the transfer of a high number of asymptomatic persons to isolation, showing the curves of *D*_2_*_y_*, *D*_2_*_o_* and *D*_2_ among circulating (a) and isolated (b) persons. We consider low transmission of infection (ϖ = 10) and without protective measures (*ε* = 1) in the isolated population.

Let us compare the total number of cases without lockdown, the lower curve in Figure 8(a), and lockdown. We assume that there is a low transmission in the isolated population, hence the number of new cases is the sum of circulating and isolated persons since after the introduction of lockdown. In Figure 17, letting *ε* = 1 in the isolated population, we show the curves of *D*_2_*_y_*, *D*_2_*_o_* and *D*_2_ with (continuous) and without (dashed) lockdown for ϖ =10 (low transmission) (a), and ϖ = 5 (medium transmission) and (b). Figure 17(a) is the sum of cases for low transmission shown in Figure 16, with the peaks being 27280 (75%), 24780 (77%) and 52040 (76%) for young, elder and total persons, occurring on 1, 3 and 2 August. For medium transmission of infection, the peaks are 43760 (120%), 40270 (126%) and 83970 (123%), occurring on 8, 11 and 9 July, shown in Figure 17(b). If we assume that isolated persons are adopting the same protective measures (face mask, hygiene, etc.) adopted by circulating persons (*ε* = 0.5), then the peaks are 25330 (69%), 22700 (71%) and 48010 (70%), occurring on 14, 16 and 15 August (figure not shown). The percentage between parentheses is the ratio between with and without lockdown *D*_2_(0.9)/*D*_2_(0), where *D*_2_(0) for young, elder and total persons are 36510, 32000, and 68460.

**Figure 16:**
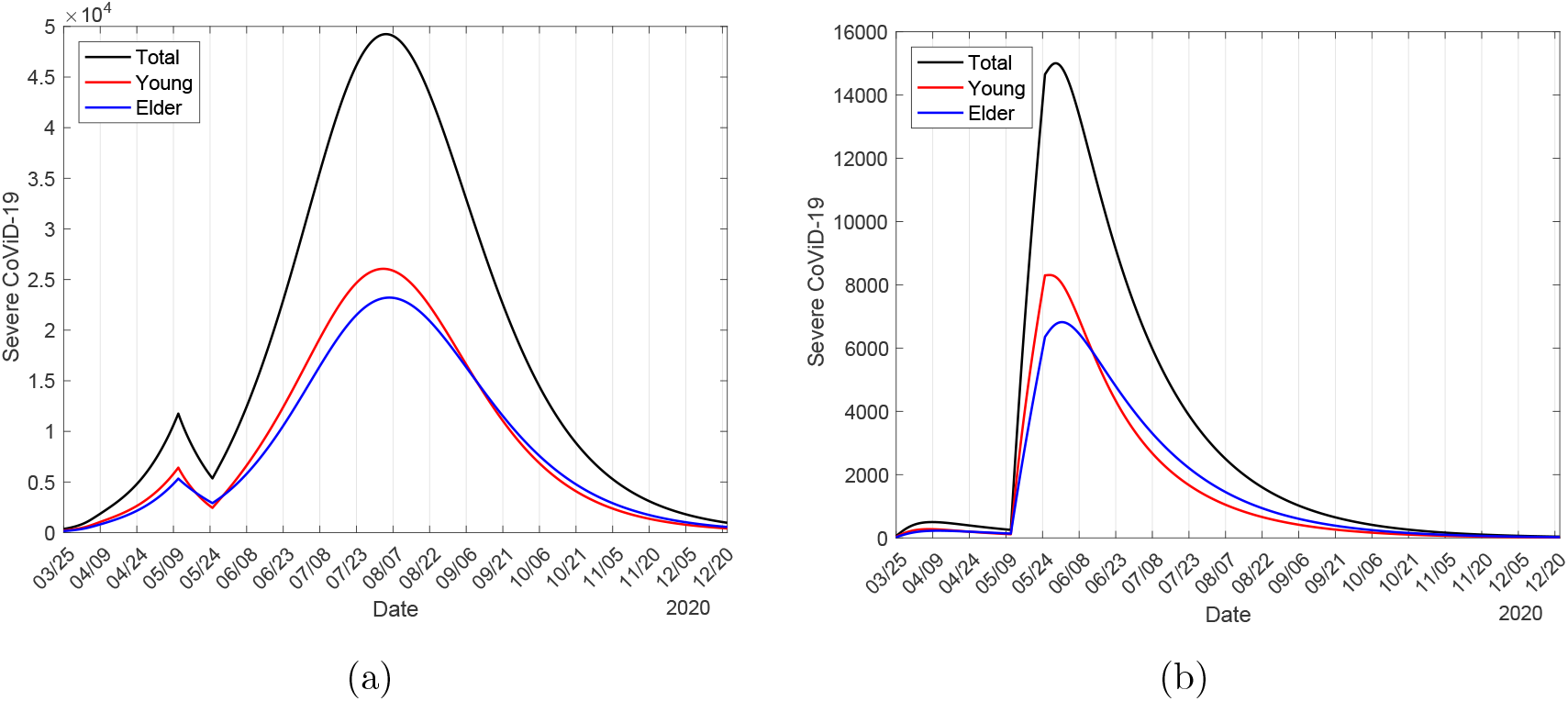
The curves of *D*_2_*<u>_y_</u>*, *D*_2_*_o_*, and *D*_2_ among circulating (a) and isolated (b) persons, assuming that among isolated persons, ϖ =10 (low transmission) and *ε* = 1.0 (without protection).

**Figure 17:**
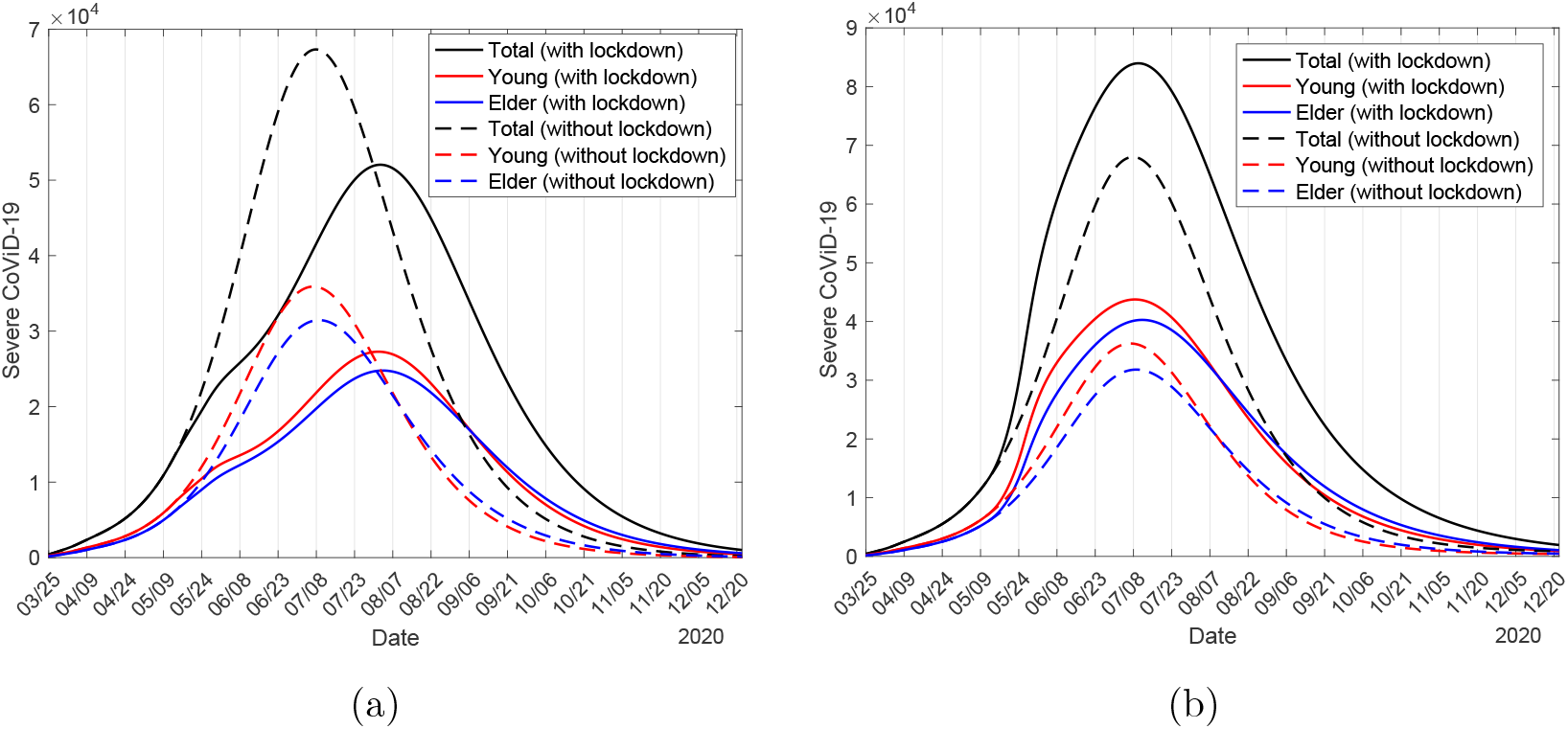
The curves of *D*_2_*_y_*, *D*_2_*_o_* and *D*_2_ with (continuous) and without (dashed) lockdown for w =10 (low transmission) (a), and ϖ = 5 (medium transmission) and (b), using *ε* = 0.5 for circulating and *ε* = 1 for isolated persons.

Therefore, considering the current epidemiological status of São Paulo State, lockdown seems conter-productive if *R*_0_ in the isolated population is relatively high. In this case, suggesting the implementation of lockdown when epidemic is growing rapidly to avoid collapse in the health system could not be a good strategy. However, lower transmission (*R*_0_ < 1) in the isolated population decreased the peak of new cases and the time of occurrence is moved to the right. Disregarding the intensity of transmission among isolated persons, the implementation of lockdown should be recommended in the beginning, or at most, at the early phase of epidemics.

Until now, we estimated the model parameters to describe the current epidemiological status of the new coronavirus in São Paulo State. Next, we evaluate the release of isolated persons, which will occur on 1 June. Let us summarize the values of the epidemiological scenario on 1 June provided by model: *S_y_*, *S_o_* and *S* are 11.0 million (43705%), 1.85 million (590095%) and 12.8 million (50391%); 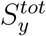, 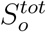 and *S^tot^* are 30.9 million (122328%), 5.4 million (1735783%) and 36.3 million (142025%); *I_y_*, *I_o_*, and *I* are 3.7 million (10%), 690300 (11%) and 4.4 million (11%); *R*_0_*_y_*, *R*_0_*_o_* and *R*_0_ are 3.86, 0.76 and 4.62; and *R_efy_*, *R_efo_* and *R_ef_* are 1.13, 0.21 and 1.34. The percentage between parentheses is the ratio between with (*k* = 0.53 and *ε* = 0.5) and without (*k* = 0 and *ε* = 1) interventions for all values. The basic reproduction number is that reduced on 4 April due to protective measures (*ε* = 0.5). There are a small number of immune persons (around 0.01% of population), but a higher number of susceptible persons (around 78% of population) at the moment of release on 1 June, indicating a rebounding of epidemics if isolation and protective measures are removed.

### 3.3 Epidemiological scenarios considering unique isolation followed by releases

To obtain epidemiological scenarios, we fix all estimated parameters: The transmission rates *β_y_* = 0.78 and *β_o_* = 0.90 (both in *days*^−1^), giving *R*_0_ = 9.24, the additional mortality rates *α_y_* = 0.00053 and *α_o_* = 0.0053 (both in *days*^−1^), the proportion in isolation of susceptible persons *k* = 0.53, and the protective factor *ε* = 0.5, which reduces the transmission rates to *α_y_* = 0.39 and *α_o_* = 0.45 (both in *days*^−1^), giving *R*_0_ = 4.62. Hereafter, all these values are fixed, unless explicitly cited.^2^

The isolation occurred on 24 March will be ended on 31 May. We consider three strategies of release, the first beginning on 1 June, the second on 23 June, and the third on 6 July. In each strategy, we consider three releases separated by 14 days, with the proportion of release at three consecutive times being *l*_1_*_j_*, *l*_2_*_j_* and *l*_3_*_j_*, for young (*j* = *y*) and elder (*j* = *o*) persons.

In strategy A, the releases occur on 1, 15 and 29 June, in strategy B, on 23 June, 7 and 21 July, and in strategy C, on 6, 20 July and 13 August. Remembering that the peaks without release for young, elder and total persons are, respectively, 36510, 32000, and 68460, which occur on 23, 25 and 23 June. Hence, in strategy A only the last release occurs after the peak (descending phase of epidemics), in strategy B, the first release situates at the peak, and in strategy C, all releases occur after the peak (descending phase) of epidemics. From Figure 10(a), the effective reproduction numbers on 1, 23 June and 6 July are, respectively, *R_ef_* = 1.33, 0.81 and 0.61.

To obtain the epidemiological scenarios of release, we solve numerically the system of equations (3), (4) and (6) with initial conditions on 26 February given by equation (B.1) in Appendix B, the boundary conditions in isolation occurred on 24 March given by equations (8) and (9), and boundary conditions of releases with first one occurring on 1 June 1 (or 23 June, or 6 July) and others, separated by 14 days, given by equations (10) and (11). There is a unique change of parameters in the system of equations, where the transmission rates *β_y_* = 0.78 and *β_o_* = 0.90 (both in *days*^−1^) were reduced to 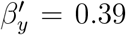 and 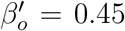 (both in *days*^−1^) on 4 April due to protective factor *ε* = 0. 5.

We evaluate the scheme where the isolated population is divided into three equal releases for the strategies A, B and C, fixing *l*_1_*_j_* = 0.33, *l*_2_*_j_* = 0.5 and *l*_3_*_j_* = 1, *j* = *y*, *o*, and varying protective factor *ε*.

#### 3.3.1 Strategy A – Release beginning on 1 June

In strategy A, the releases occur on 1, 15 and 29 June. On 1 June, *R_ef_* = 1.33 jumps up to a higher value *R_ef_* = *R_u_* = 2.14, a gap of 0.81, where the increased reproduction number *R_u_* is given by equation (26), and this increased *R_ef_* decreases until the next release, and so on.

In Table 5 we show the values and times of occurrence of peaks of *D*_2_, and the numbers of accumulated cases Ω, immune *I*, cured *C* and susceptible *S* persons at the end of the first wave of epidemics, on 10 February 2021, for *ε* = 0.2, 0.3, 0.4 and 0.5.

**Table 5:** The values and times of the occurrence of peaks of *D*_2_, and the numbers of accumulated cases Ω, immune *I*, cured *C* and susceptible S persons at the end of the first wave of epidemics, on 10 February 2021, for *ε* = 0.2, 0.3, 0.4 and 0.5. For Strategy A.

| | Peak 1 | Date | Peak 2 | Date | $\Omega$ ($10^5$) | $I$ ($10^7$) | $C$ ($10^5$) | $S$ ($10^7$) |
| --- | --- | --- | --- | --- | --- | --- | --- | --- |
| $\varepsilon = 0.5$ | 217600 | 17/7/20 | - | - | 9.30 | 4.34 | 8.68 | 0.12 |
| $\varepsilon = 0.4$ | 188800 | 22/7/20 | - | - | 9.10 | 4.24 | 8.50 | 0.21 |
| $\varepsilon = 0.3$ | 142600 | 30/7/20 | - | - | 8.59 | 3.99 | 8.02 | 0.46 |
| $\varepsilon = 0.2$ | 57740 | 12/6/20 | 74520 | 8/8/20 | 7.01 | 3.23 | 6.54 | 1.22 |

From Table 5, the peak 217600 (56% of without interventions) on 7 July, for *ε* = 0.5, decreases and moves to the right as *ε* increases. However, for *ε* = 0.2, we observe that the peak reaches 57740 (15% of without interventions) on 12 June, avoiding the peak on 23 June without release (68460, 18% of without interventions), but the reduced transmission rates are not sufficient to avoid the second peak 74520 (19% of without interventions) on 8 August due to further releases. We illustrate two protective factors in Figure 18.

In Figure 18, we show strategy A with the same protective factor ε = 0.5 being maintained after release (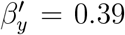and 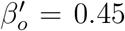 (both in *days*^−1^), giving *R*_0_ = 4.62) (a), and reduced *ε* = 0.2 adopted by all circulating persons after release (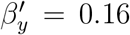 and 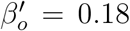 (both in *days*^−1^), giving *R*_0_ = 1.85) (b). For *ε* = 0.5, the peaks for young, elder and total persons are, respectively, 120400 (53%), 97300 (59%), and 217600 (56%), which occur on 16, 18 and 17 July, while for *ε* = 0.2, the higher second peak are 38660 (17%), 35860 (22%), and 74520 (19%), which occur on 7, 8 and 8 August. In Figure 18(a), we observe a small perturbation following the first release, which is not considered a peak. The percentage between parentheses is the ratio between with and without interventions *D*_2_(*k*, *ε*)/*D*_2_(0,1), remembering that the peaks of epidemics without interventions for young, elder and total persons are, respectively, 226600, 164800, and 391000.

**Figure 18:**
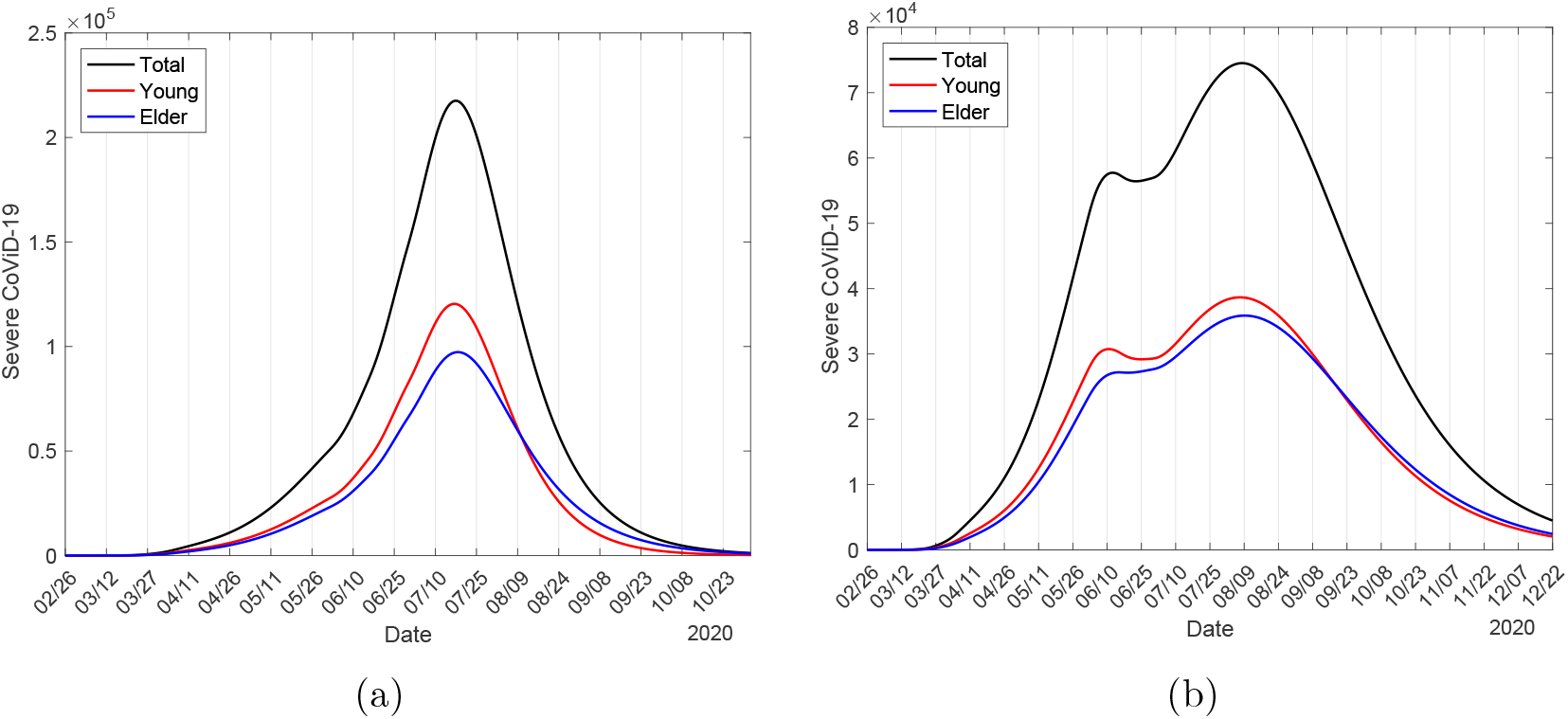
Strategy A with the same protection factor *ε* = 0.5 in isolation being maintained after release (a), and reduced *ε* = 0.2 adopted by all circulating persons after release (b).

From Table 5 and Figure 18, we observe that the decreasing in transmission rates due to increased protective measures (factor *ε* is decreased) becomes the epidemiological scenario with release less harmful, decreasing by around 34% when *ε* decreases from 0.5 to 0.2. Lower *ε* decreases the transmission rates, reducing *R_ef_*.

We present the curve of the approximated effective reproduction number *R_ef_* using equation (24). When the isolation begins, on 25 March, this number jumps down and decreases to *R_r_*, given by equation (25), but *R*_0_ = 9.24 does not change. On 4 April, the transmission rates decrease to 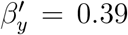 and 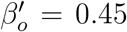 (both in *days*^−1^) due to the protective factor *ε* = 0.5, reducing the basic reproduction number to *R*_0_ = 4.62 and, consequently, *R_ef_* jumps down (see Figure 10). However, at the time of release, *R_ef_* jumps up due to the increased number of susceptible persons, given by equation (26), if ε is adopted by isolated persons after release. However, if protective measures are increased after release, then *R_ef_* can jump up or down depending on the factor *ε*. In Figure 19, we show the curves of *R_ef_* and *D*_2_ for the same protective factor when released *ε* = 0.5 (a) and increased protective measures after release to *ε* = 0.2 (b), where *D*_2_ was divided by 25000 and 10000, respectively, for *ε* = 0.5 and 0.2.

From Figure 19, on 26 February we have *R_ef_* = *R*_0_ = 9.24, on 24 March *R_ef_* = 9.22 jumps down to *R_r_* = 4.35, on 4 April *R_ef_* = 4.29 jumps down to = 2.15, and reaches on 1 June *R_ef_* = 1.33 just before the beginning of release. For release with *ε* = 0.5 (between parentheses, for *ε* = 0.2), on 1 June *R_ef_* = 1.33 jumps up to *R_u_* = 2.14 (jumps down to *R_r_* = 0.85), decreases to *R_ef_* = 1.48 (0.76) on 15 June, and jumps up to *R_u_* = 2.29 (1.01), and decreases to *R_ef_* = 1.20 (0.98) on 29 June, and jumps up to *R_u_* = 2.01 (1.30), and decreases continuously.

**Figure 19:**
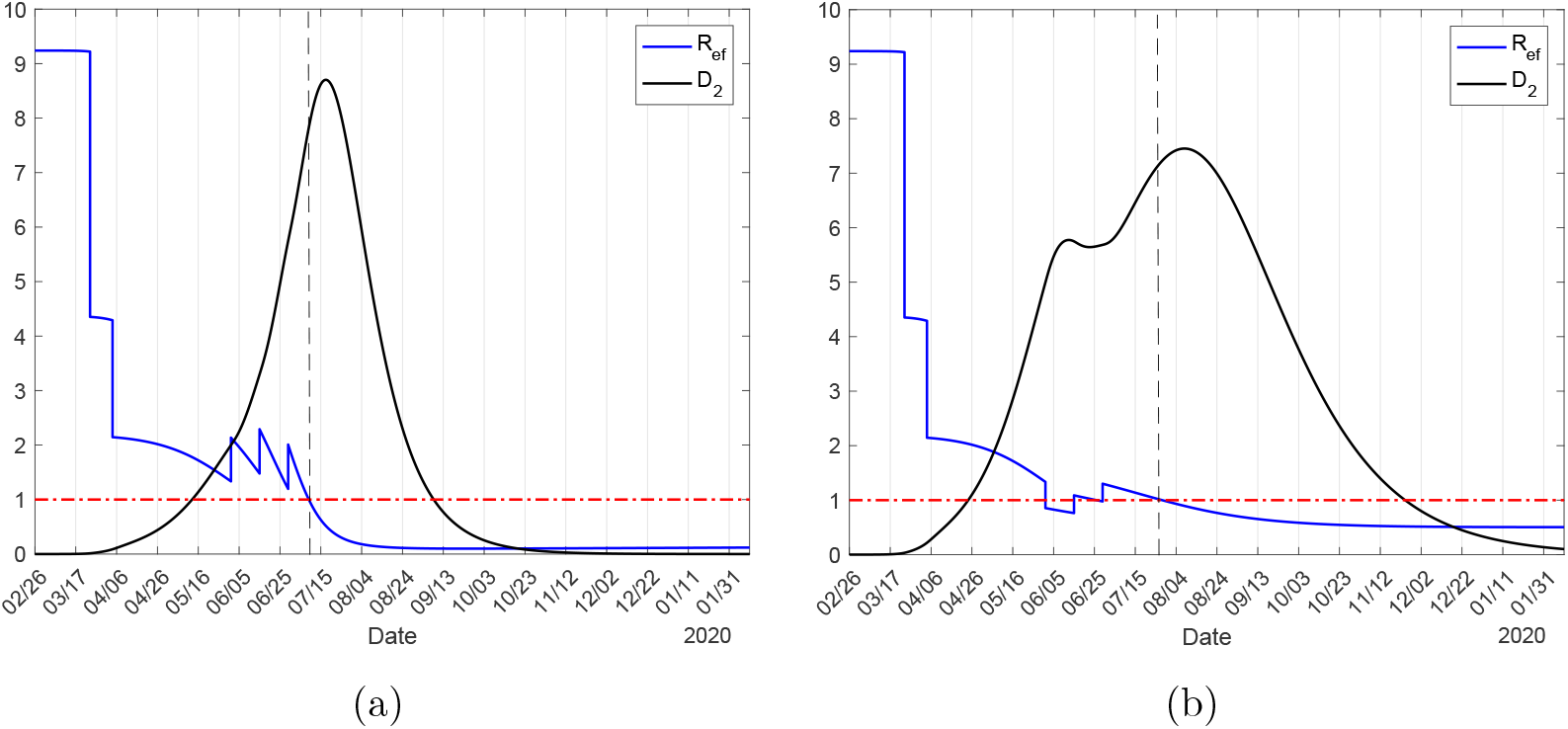
The curves of *R_ef_* and *D*_2_ for the same protection factor when released *ε* = 0.5 (a) and increased protection actions after release to *ε* = 0.2 (b), where *D*_2_ must be multiplied by 25, 000 and 10, 000, respectively, for *ε* = 0.5 and 0.2.

#### 3.3.2 Strategy B – Release beginning on 23 June

In strategy B, the releases occur on 23 June 23, 7 and 21 July. On 23 June, in the first release, *R_ef_* = 0.81 jumps up to *R_ef_* = *R_u_* = 1.61, a gap of 0.80. In Table 6, we show the values and times of occurrence of peaks of *D*_2_, and the numbers of accumulated cases Ω, immune *I*, cured *C* and susceptible *S* persons at the end of the first wave of epidemics, on 1 April 2021, for *ε* = 0.2, 0.3, 0.4 and 0.5.

**Table 6:** The values and times of the occurrence of peaks of *D*_2_, and the numbers of accumulated cases Ω, immune *I*, cured *C* and susceptible *S* persons at the end of the first wave of epidemics, on 1 April 2021, for *ε* = 0.2, 0.3, 0.4 and 0.5. For Strategy B.

| | Peak 1 | Date | Peak 2 | Date | $\Omega$ ($10^5$) | $I$ ($10^7$) | $C$ ($10^5$) | $S$ ($10^7$) |
| --- | --- | --- | --- | --- | --- | --- | --- | --- |
| $\varepsilon = 0.5$ | 170100 | 12/8/20 | - | - | 9.22 | 4.30 | 8.60 | 0.16 |
| $\varepsilon = 0.4$ | 142700 | 16/8/20 | - | - | 8.96 | 4.17 | 8.36 | 0.29 |
| $\varepsilon = 0.3$ | 68460 | 24/8/20 | 102000 | 23/8/20 | 8.34 | 3.87 | 7.78 | 0.59 |
| $\varepsilon = 0.2$ | 68450 | 23/8/20 | 50290 | 20/8/20 | 6.63 | 3.06 | 6.18 | 1.40 |

From Table 6, the peak 170100 (44% of without interventions) on 12 August for *ε* = 0.5 decreases and moves to the right as *ε* increases. However, for *ε* = 0.3, we observe that the first peak reaches the same value for that without release 68460 (18% of without interventions) on 24 June, but the reduced transmission rates are not sufficient to avoid the second peak 102000 (26% of without interventions) due to further release. However, for *ε* = 0.2, the second peak 50290 (13% of without interventions) occurring on 20 August is lower than the first peak, and the goal of isolation in reducing the peak of severe CoViD-19 cases is achieved.

The curves of epidemics in strategy B follow similar than those shown in strategy A. For *ε* = 0.5, the curves are more spread and the small perturbation in the first release, higher than strategy C, is not considered as a peak again. For *ε* = 0.2, the second peak is lower than the first in comparison with strategy A.

#### 3.3.3 Strategy C – Release beginning on 6 July

In strategy C, the releases occur on 6, 20 July and 3 August. On 6 July, in the first release, *R_ef_* = 0.61 jumps up to *R_ef_* = *R_u_* = 1.41, a gap of 0.80. In Table 7, we show the values and times of occurrence of peaks of *D*_2_, and the numbers of accumulated cases Ω, immune *I*, cured C and susceptible S persons at the end of the first wave of epidemics, on 10 February 2021, for *ε* = 0.2, 0.3, 0.4 and 0.5.

**Table 7:**
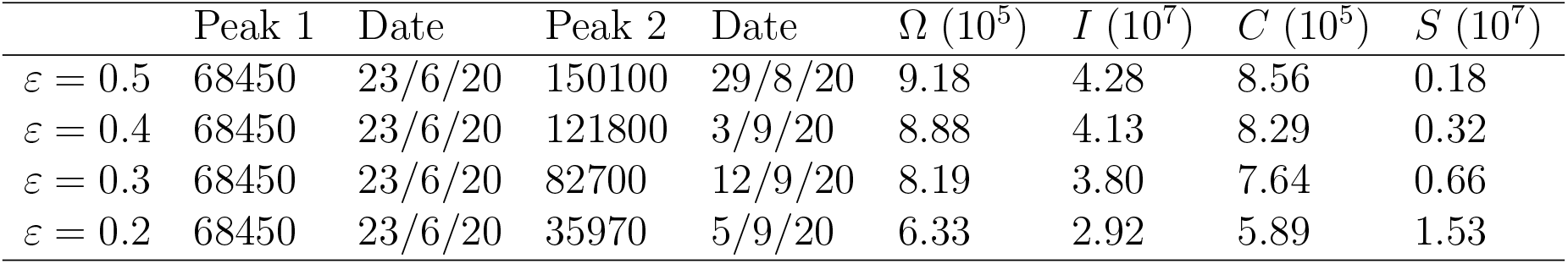
The values and times of the occurrence of peaks of *D*_2_, and the numbers of accumulated cases Ω, immune *I*, cured *C* and susceptible *S* persons at the end of the first wave of epidemics, on 1 April 2021, for *ε* = 0.2, 0.3, 0.4 and 0.5. For Strategy C.

From Table 7, as expected, for all values of ε, the first peak reaches 68460 on 23 June, but the reduced transmission rates are not sufficient to avoid the second peak due to further release. The second peak is delayed and decreased as *ε* decreases, which is higher than the first. For *ε* = 0.5, the second peak 150100 (38% of without interventions) occurs on 29 August, while for *ε* = 0.3, the second peak 82700 (21% of without interventions) occurs on 12 September. However, when *ε* = 0.2, the second peak 34970 (9% of without interventions) is lower than the first, representing not a rebounding of epidemics but a perturbation of some magnitude, which disappears for more lower *ε*. Again, the goal of isolation in reducing the peak of severe CoViD-19 cases is achieved with *ε* = 0.2. For higher *ε*, as the number of severe CoViD-19 cases decreases slowly, the number of susceptible persons also decreases slowly as well as the number of immune persons increases slowly, showing that the second wave of epidemics can be triggered earlier and intense if protective measures are abandoned after release.

The curves of epidemics in strategy C follow similar than those shown in strategy B. For *ε* = 0.5, the curves are more spread and the small perturbation in the first release, higher than strategy C, can be considered as a peak now. For *ε* = 0.2, the second peak is lower than the first in comparison with strategy B, which can be considered as a small perturbation.

## 4 Discussion

The system of equations (3), (4) and (6) is simulated to provide epidemiological scenarios using parameters estimated from data collected in São Paulo State [3] from 26 February to 7 May 2020 (see Figure B.1 in Appendix B). Based on those data, we estimated the transmission rates *β_y_* = 0.78 and *β_o_* = 0.90 (both in *days*^−1^), giving *R*_0_ = 9.24 (partials *R*_0_*_y_* = 7.73 and *R*_0_*_o_* = 1.5), and the additional mortality rates *α_y_* = 0.00053 and *α_o_* = 0.0053 (both in *days*^−1^). Isolation was introduced on 24 March, hence, we estimated the proportion in isolation of susceptible persons *k* = 0.53, which was the average proportion of daily proportions observed in São Paulo State from 26 February to 3 May. Nevertheless, the observed data suggested an additional decrease in the transmission, and we estimated the protective factor ε = 0.5 since 4 April, which reduced the transmission rates to *β_y_* = 0.39 and *β_o_* = 0.45 (both in *days*^−1^), giving *R*_0_ = 4.62. Hence, from 26 February to 3 April, the epidemics was driven by the force of infection with *R*_0_ = 9.23, and since 4 April 4, by the force of infection with *R*_0_ = 4.62. Additionally, the daily registered cases of CoViD-19 were affected by the proportion in isolation around 9 days ago, which, together with the values found in the literature, confirmed our estimated about incubation period and the period of infectivity of pre-diseased persons. Moreover, this observed delay helped us in the choice of appropriate period of observed data to estimate the model parameters.

Based on the estimated parameters, we evaluated the current epidemiological status of São Paulo State. The model parameters were estimated using Ω, given by equation (13), and the accumulated data (see Figures 2, 3, 5 and 7). From this estimated curve Ω we retrieved the curve of severe CoViD-19 *D*_2_, indicating that the peak reaches 68460 on 23 June, and at this day, the accumulated cases Ω is 243000. From the curve of estimated Ω, we also retrieved daily cases of CoViD-19 (see Figure 11) using Ω*_d_* given by equation (15), indicating that on 12 June the peak reaches 5287 cases. We also estimated the ratio hidden:apparent cases of new coronavirus as epidemics evolves (see Figures 12 and 13), suggesting around 25 asymptomatic cases by one symptomatic person at the peak of epidemics.

In the absence of interventions, the peak of severe Covid-19 cases should reach 391000 on 3 May. However, the implementation of isolation decreased the peak to 139300 (36% of without interventions) and delayed to 22 May, and the addition of protective measures to avoid new coronavirus infections reduced the peak to 68460 (18% of without interventions), which will occur on 23 June. The isolation alone demonstrated some reduction in the transmission of infection, which was enhanced by the adoption of protective measures by population.

In [31] and [32] we estimated the additional mortality rates based on the observed data and concluded that their values did not provide long-term reliable values. For instance, this estimation resulted in deaths of 30% up to 80% of severe CoViD-19 cases at the end of the first wave of epidemics. For this reason, we estimated considering the accumulated deaths at the end of the first wave of epidemics to be around 10% of elder persons, disregarding the observed data. Here, we estimated the daily deaths data considering that these data corresponded to the number of new cases occurred 9 days ago (see Figure 3(a)). Indeed, the estimated additional mortality rates agreed with those estimated in [32] taking into account deaths at the end of the first wave.

As we had hypothesized in [32], the estimation of the protective factor *ε* = 0.5 not only fitted well the accumulated severe CoViD-19 cases, but also subsequent data followed closely the estimated curve. Hence, the use of face mask, hygiene behaviour and social distancing reduced substantially the number of cases, indicating a way to control new coronavirus epidemics. Moreover, the estimated proportion of population in isolation, *k* = 0.53 agreed with the average proportion observed in São Paulo State.

The number of new cases at each time is determined by *R_ef_*. In Figure 20, we show the curve of *R_ef_* in the epidemics with isolation and *R_u_* at the time of the first release for *ε* = 0.5 (a), and for *ε* = 0.5, 0.4, 0.3 and 0.2 (b). The curve *R_u_* is the jump up of the curve *R_ef_* at the first release (the distance of the vertical line intercepting curves *R_ef_* and *R_u_* is the gap of jump). Only *ε* = 0.5 does not reach value lower than 1 at any time of the first release.

**Figure 20:**
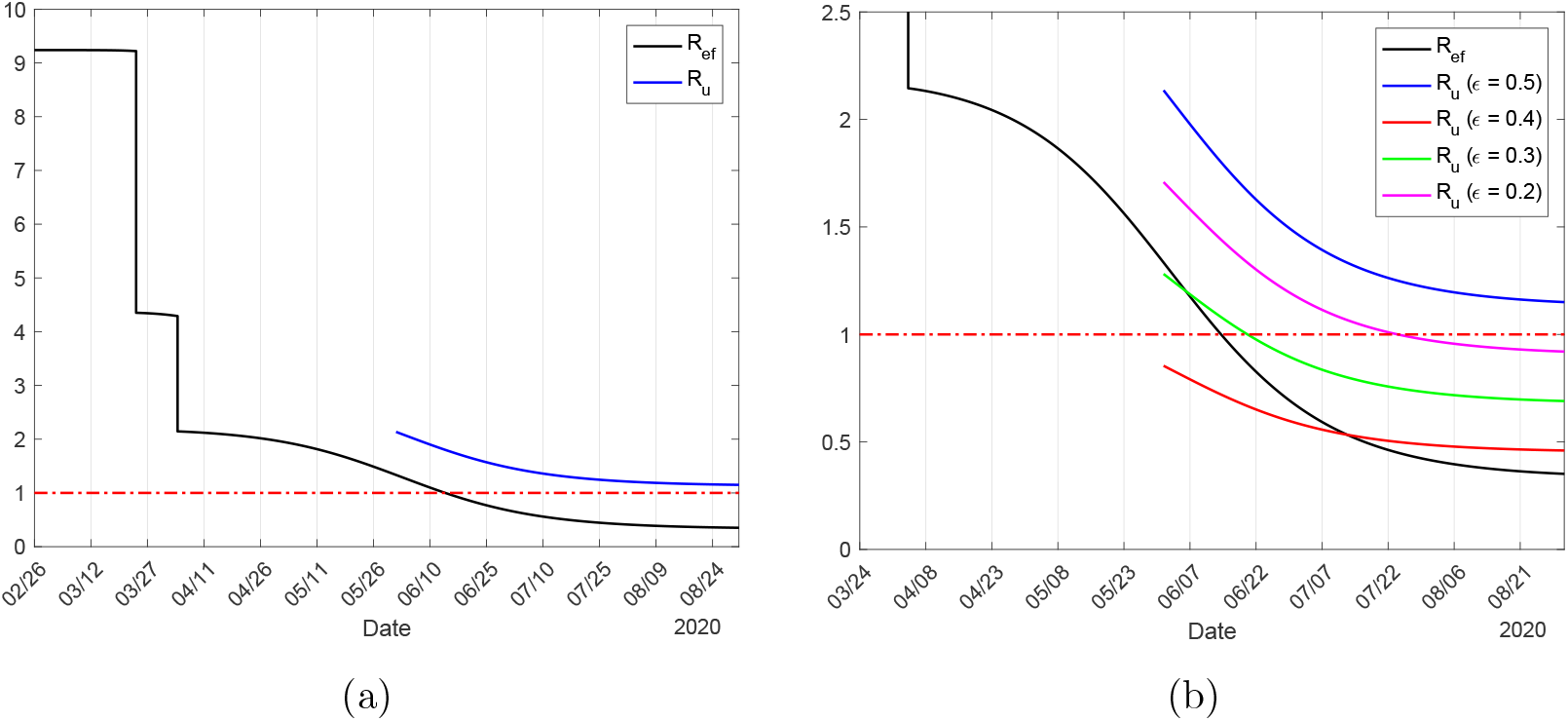
The curve of *R_ef_* in the epidemics with isolation, and *R_u_* at the time of the first release for *ε* = 0.5 (a), and for *ε* = 0.5, 0.4, 0.3 and 0.2 (b). The curve *R_u_* is the jumps of the curve *R_ef_* at the first release (vertical line intercepting curves *R_ef_* and *R_u_* is the gap of jump).

Using SIR model, we estimated the basic reproduction number using the data collected in São Paulo State, obtaining around *R*_0_ = 2.8, which is 3-times lower than our estimate. Additionally, from SIR model, the peak of 10 million occurred on 27 June, reaching 40 million of accumulated cases, and the number of deaths is 9.24 million, which is 23% of all cases, and 17-times higher than our estimate (55040).

The concept of herd immunity is associated with the protection provided by immune persons to a specific subpopulation under higher risk of comorbidity. For instance, in rubella infection, mass vaccination was planned to diminish infection among pregnant women aiming the decrease in the number of congenital rubella syndrome [29]. The misunderstanding of the concept of herd immunity associated with the lack of knowledge about the fatality of CoViD-19, and the rapid dissemination of underestimated *R*_0_ around 2, may have lead to misconducting public health policies. For instance, the United Kingdom (in the first moment) and Sweden adopted the idea of immunization by natural infection, believing that the “herd immunity” could protect the population. However, the concept of herd immunity must be understood not immunization by the natural course of epidemics but due to the vaccine, which decreases (jumping down) the effective reproduction number *R_ef_*. Figure 20 exemplifies “herd immunity”, that is, transitory protection of the population not from vaccine but external factors (isolation and protective measures), which we call “herd protection”. Contrarily to the herd immunity provided by vaccine, herd protection provided by isolation and protective measures increase *R_ef_* when external factors cease. If a reliable estimation of *R*_0_ (around 9) should be available at the beginning of epidemics, maybe the misconducting of public health policies had not been adopted. However, at the beginning of pandemics, the higher estimation of deaths by SIR-type model resulted in the adoption of isolation as a control mechanism, not the reliable estimation of *R*_0_.

We also studied the potential risk of introducing lockdown depending on the current epidemiological status. When, hypothetically, transmission occurs in non-interacting young and elder subpopulations, in long-term epidemics, they reach 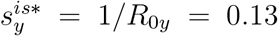 and 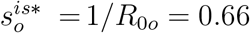. However, from numerical simulations without releasing, we obtained 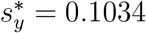 and 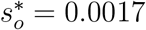 when these subpopulations are interacting. Notice that the difference 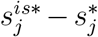, *j* = *y*, *o*, is the additional proportion of susceptible persons infected due to interaction, being 2.7% for young and 66% for elder persons, showing that elder persons are 24-times more risk than young persons when interacting. For this reason, if lockdown is implemented, assuming that there is low transmission (through restricted contact occurring in the household and/or neighborhood) in the isolated population due to asymptomatic persons locked-down, elder persons are under more risk, and consequently, deaths can increase.

Based on the accumulated severe CoViD-19 cases, we estimated the occupancy of beds in hospital, deaths and cured persons (see Figures C.1, C.2 and C.3 n Appendix C). To estimate the proportions of deaths occurring in severe CoViD-19 cases (see Appendix C), we considered that deaths correspond to new cases occurred 15 days ago. On 19 May, we observed a delay of around 15 days between high numbers of deaths and severe cases [1]. On 1 June, the beginning of release, the total number of deaths is around 8500 (see Table C.1 in Appendix C).

We studied possible scenarios of releasing by considering three different times for the first release, the strategies A (first release on 1 June, with *R_ef_* = 1.33), B (first release on 23 June, with *R_ef_* = 0.81) and C (first release on 6 July, with *R_ef_* = 0.61). The approximated effective reproduction number *R_ef_* was calculated using equation (24). The number of severe CoViD-19 cases in the peak of epidemics provided by the model (see Tables 5. 6 and 7) is important to support the choice of releasing strategy by decision-makers to avoid the collapse in the health care system.

Analyzing the releasing strategies, we observed that increase in the protective measures can manage the epidemics in terms of available hospital beds and the number of deaths, which decreased 75% when ε is decreased from 0.5 to 0.2. Maybe the massive educational campaign can achieve the goal by increasing the protective measures, that is, decreasing ε for at least 0.2. However, the three strategies (different time in the first release) presented very little difference (around 5%) among them, due to *R_ef_* being close to one. Hence, in all strategies where the protective factor is maintained at 50%, the health care system may collapse. However, if the protective factor could be increased to 80%, the health care system may not collapse. The question is: what kind and how intensity protective measures must be adopted by the population to decrease the force of infection?

In all strategies, the number of all deaths decreased around 97% (*ε* = 0.5) and 75% (*ε* = 0.2) in comparison with epidemics without interventions (55040). These numbers were obtained considering that all patients are receiving adequate treatment. However, the number of beds is limited, as well as health care workers. Hence, let us compare the peaks of occupancy of all beds with the maximum occupancy of beds 252200 in a epidemics without any interventions (*k* = 0 and *ε* =1). When isolation and protective measures are adopted by population (*k* = 0.53 and *ε* = 0.5), the maximum occupancy of beds is 38360, reduction by 15%, while the number of deaths was reduced by 42%. For instance, in strategy A with *ε* = 0.5, the maximum occupancy of bed 217600 is 50%, and 54240 deaths are 98.5% of that without any interventions. If São Paulo State has 38360 beds, hence 22892 will be the actual deaths due to lethality of CoViD-19, but with release, the surplus 179240 with respect to currently adopted isolation and protection (38360) will not be treated, and a high percentage should die. More drastic should be in the case without any interventions, when surplus 213840 will not be treated. The question is: how many people without hospital care will survive?

Due to the lack of a reliable number of occupied beds in hospital (inpatients and ICU), the scenarios of the occupancy of beds in the hospitals presented here may be over estimated. This over estimation could be due to, among many factors, improved protocols in the treatment of CoViD-19, and increased number of tests among severe CoViD-19 cases, which included individuals manifesting symptoms, but not needing long-stay hospital care. These new scenarios of CoViD-19 transmission are left to a future. Additionally, model parameters were estimated to describe the isolation and protective measures affecting on the current epidemiological status and subsequent release. However, from Figure 6(a), we observed a lowering in severe CoViD-19 data with respect to the estimated curve, and more data must be considered to decide whether this is a tendency or not, once the observed proportions in isolation in São Paulo State did not change in this period. There are many factors, among them we can cite the new coronavirus being irradiated to small cities from São Paulo City, and more increased protective measures.

## 5 Conclusion

We formulated a mathematical model considering two subpopulations comprised by young and elder persons to study CoViD-19 in São Paulo State, Brazil. The model considering pulses in isolation and release was simulated to describe the current epidemiological status in São Paulo State and future scenarios when release will occur on 1 June.

Isolation as well as lockdown are valuable measures to control epidemics with high lethality. However, lockdown with a short period would not be an appropriate control effort if epidemic is in the ascending phase, and non-negligible transmission in the isolated population can occur. The transferring of an elevated number of asymptomatic persons by lockdown to isolation may trigger new epidemic in a previously isolated population. A false feeling of absence of transmission in the isolated population could be prevailed due to the lower initial estimation of the basic reproduction number, around *R*_0_ = 2.5. However, our model estimated *R*_0_ = 9.24, which may increase the possibility of transmission in isolated population due to airborne transmission.

The comparison of the suitability and appropriateness of isolation and lockdown based on the number of deaths is not reasonable, rather, the peak of severe CoViD-19 must be taken into account. This number is proportional to the occupancy of beds (around 55% of severe cases need hospital care), but the numbers of beds and health care workers are limited, hence we must think in the surplus of severe CoViD-19 cases with respect to the limitation of the health care system.

From the epidemiological scenarios of release, we observed that the delay to implement the release (1, 23 June and 6 July) affected very little reducing by around 5% in the number of occupancy of beds and deaths. The effective reproduction number *R_ef_* in these three strategies was around 1, hence postponing the release of isolated persons when *R_ef_* decreases more could be a better strategy. However, the increase in protective measures, reducing *ε* from 0.5 to 0.2, reduced by around 15% the occupancy of beds and deaths (these numbers are proportional to severe CoViD-19 cases). Hence, the release beginning on 1 June could be implemented if accompanied by a massive educational campaign to adopt protective measures.

Finally, severe CoViD-19 data collected in the São Paulo State indicates that there is another lowering in those cases besides the diminishing resulted from isolation and protection. We hypothesized that this decreasing could be due to the spreading of new coronavirus to small cities irradiated from São Paulo City, which changes in demography (population density), but more data are needed to confirm or deny this hypothesis. The model considered homogeneous space (population density) and time (constant parameters) in São Paulo State. However, this model can be applied to cities taking into account their population densities, for instance. Notice that considering 44.6 million of inhabitants and *R*_0_ = 9.24 in São Paulo State, the epidemics will be established strongly due to the threshold to trigger epidemics is very low, *N*^th^ = 4.83 million of persons.

## Data Availability

The data that support the findings of this study are openly available in Ministry of Health (Brazil) at https://covid.saude.gov.br.

## Author contributions

Hyun Mo Yang: Conceptualization, Methodology, Formal analysis, Writing - Original draft preparation, Supervision. Luis Pedro Lombardi Junior: Software, Data Curation, Visualization. Fabio Fernandes Morato Castro: Validation, Investigation. Ariana Campos Yang: Validation, Investigation.

## A Trivial equilibrium and its stability

By the fact that *N* is varying, the system is non-autonomous non-linear differential equations.

To obtain autonomous system of equations we let *k_j_* = *l_ij_* = 0, *j* = *y*, *o*, and use fractions of individuals in each compartment, defined by, with *j* = *y* and *o*,

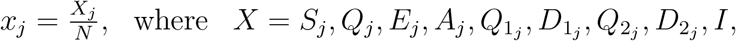

resulting in

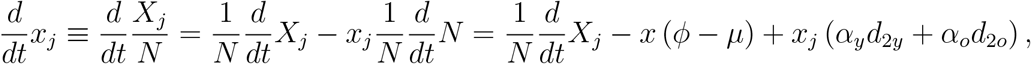

using equation (5) for *N*. Hence, equations (2), (3) and (4) in terms of fractions become, for susceptible persons,

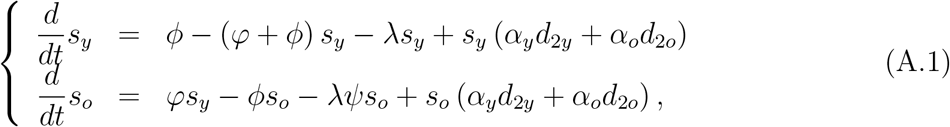

for infected persons,

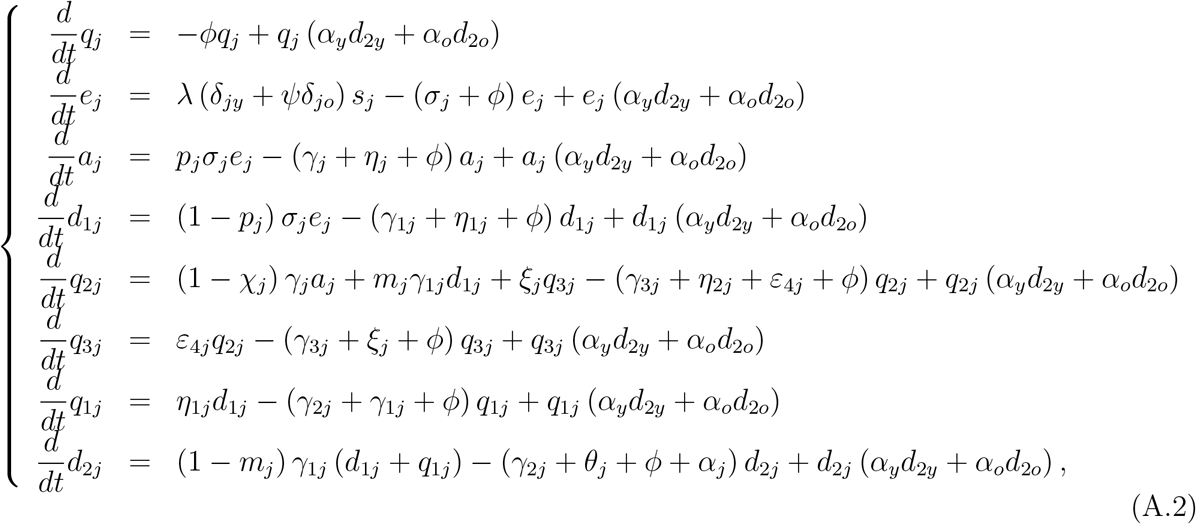

and for immune persons

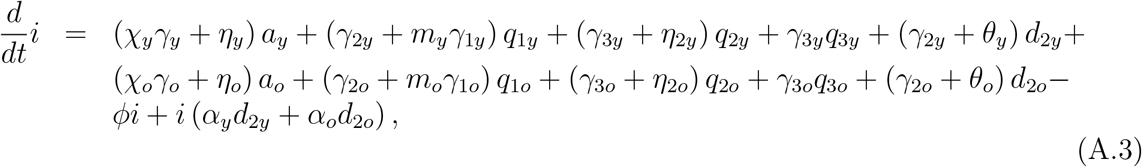

where λ is the force of infection given by equation (1) re-written as

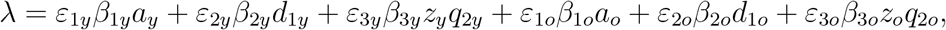

and

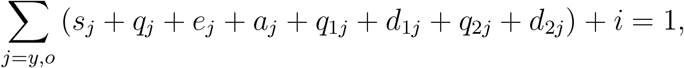

which is autonomous system of equations. We remember that all classes vary with time, however their fractions attain steady state (the sum of derivatives of all classes is zero). This system of equations is not easy to determine the non-trivial (endemic) equilibrium point *P*\*. Hence, we restrict our analysis with respect to the trivial (disease free) equilibrium point.

The trivial or disease free equilibrium *P*^0^ is given by

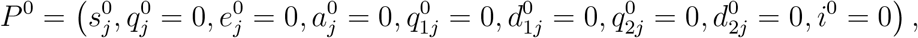

for *j* = *y* and *o*, where

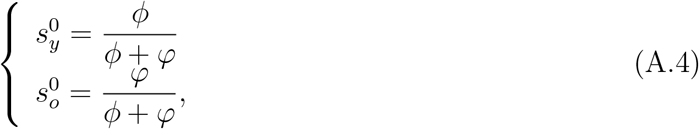

with 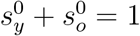.

Due to 17 equations, we do not deal with characteristic equation corresponding to Jacobian matrix evaluated at *P*^0^, but we apply the next generation matrix theory [7].

The next generation matrix, evaluated at the trivial equilibrium *P*^0^, is obtained considering the vector of variables *x* = (*e_y_*, *a_y_*, *d*_1_*_y_*, *e_o_*, *a_o_*, *d*_1_*_o_*). We apply method proposed in [25] and proved in [26]. There are control mechanisms (isolation), hence we obtain the reduced reproduction number *R_r_* by isolation.

In order to obtain the reduced reproduction number, diagonal matrix *V* is considered. Hence, the vectors *f* and *v* are

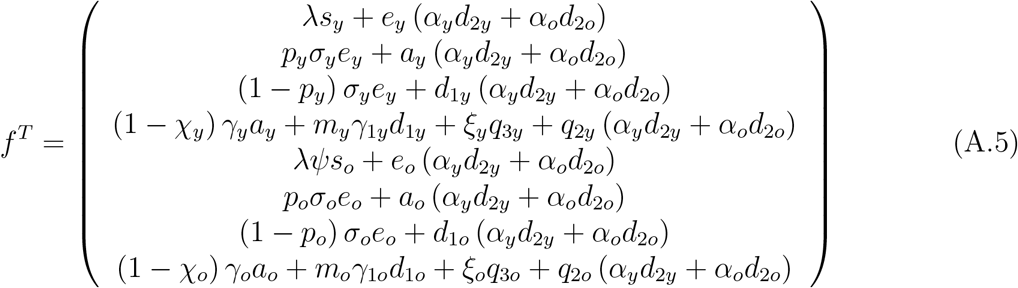

and

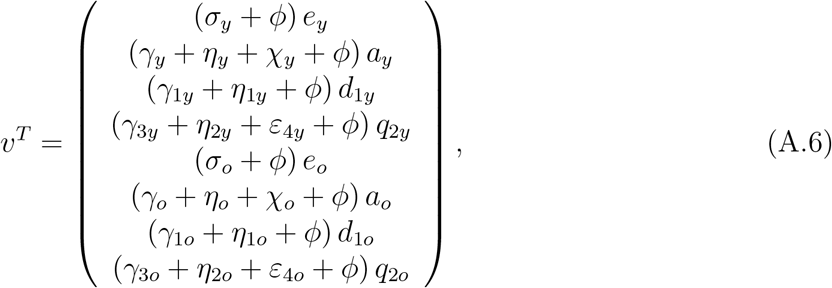

where the superscript *T* stands for the transposition of a matrix, from which we obtain the matrices *F* and *V* (see [7]) evaluated at the trivial equilibrium *P*^0^, which were omitted. The next generation matrix *FV*^−1^ is

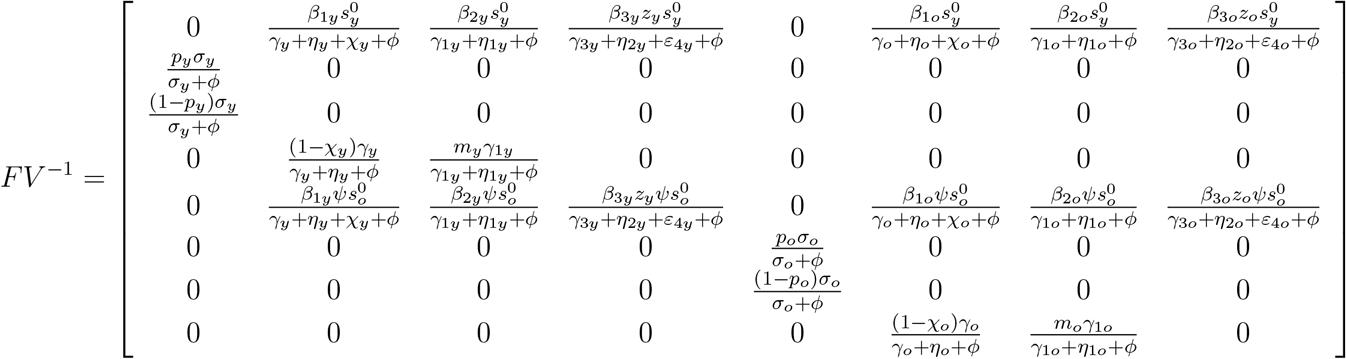

and the characteristic equation corresponding to *FV*^−1^ is

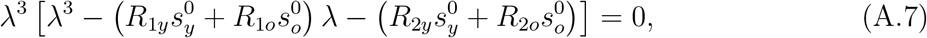

where we have

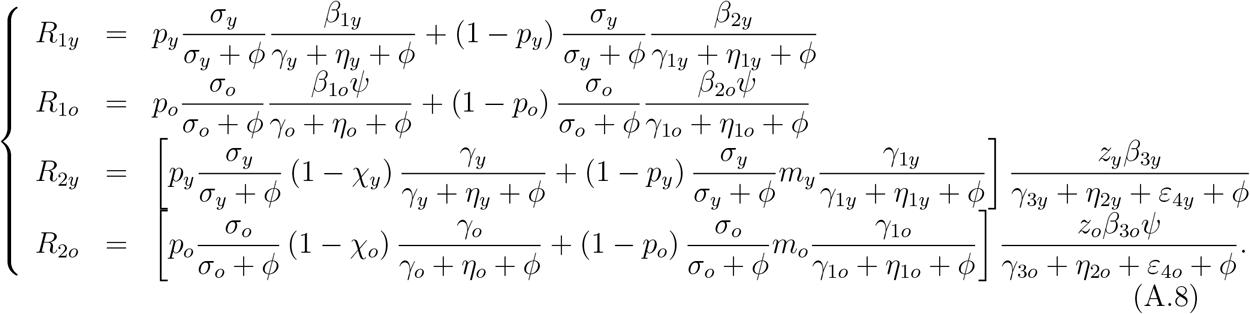

Instead of using the spectral radius ρ (*FV*^−1^), which is not easy to evaluate, we apply procedure in [25] (the sum of coefficients of characteristic equation), resulting the basic reproduction number *R*_0_ given by

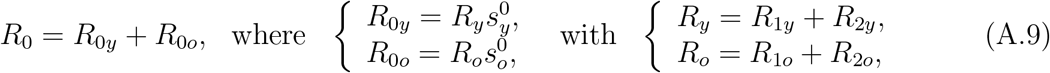

Hence, the trivial equilibrium point *P*^0^ is locally asymptotically stable if *R*_0_ < 1.

In [23] we showed, when *z_y_* = *z_o_* = 0, that the inverse of the basic reproduction number *R*_0_ is the fraction of susceptible persons in the steady state. But, according to [30], the inverse of the basic reproduction number *R*_0_ given by equation (A.9) is a function of the fraction of susceptible individuals at endemic equilibrium *s*\* through

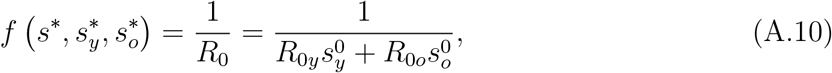

where 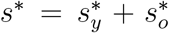 (see [28] [30]). For this reason, the effective reproduction number *R_ef_* [27], which varies with time, can not be defined neither by *R_ef_* = *R*_0_ (*s_y_* + *s_o_*), nor *R_ef_* = *R*_0_*_y_s_y_* + *R*_0_*_o_s_o_*. The function *f* (*ϰ*) is determined by calculating the coordinates of the nontrivial equilibrium point *P*\*. For instance, for dengue transmission model, 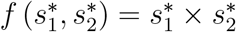, where 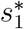 and 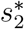 are the fractions at equilibrium of, respectively, humans and mosquitoes [28]. For tuberculosis model considering drug-sensitive and resistant strains, there is not *f* (*x*), but *s*\* is solution of a second degree polynomial [30]. From equation (A.10), let us assume that 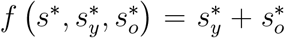. Then, we can define the approximated effective reproduction number *R_ef_* as

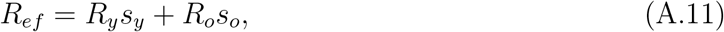

which depends on time, and when attains steady state (*R_ef_* = 1), we have *s*\* = 1/*R*_0_.

The basic reproduction number *R*_0_ is the secondary cases produced by one case of infectious person (could be anyone in one of classes harbouring virus) in a completely susceptible young and elder persons without control. Let us understand *R*_1_*_j_* and *R*_2_*_j_*, *j* = *y*, *o*, stressing that the interpretation is the same for both subpopulations, hence we drop out subscript *j*. To facilitate the interpretation, we consider this infectious person in exposed class *E*. This person enters into one of the infectious class composed by asymptomatic (*A*), pre-diseased (*D*_1_) and a fraction of mild CoViD-19 (*Q*_2_).

1. *R*_1_ takes into account the transmission by one person in asymptomatic A or pre-diseased *D*_1_ class. We interpret for asymptomatic person transmitting (between parentheses, for pre-diseased person) infection. One infectious person survives during the incubation period with probability *σ*/ (*σ* + *ϕ*) and enters into asymptomatic class with probability *p* (pre-diseased, with 1 − *p*) and generates, during the time 1/ (*γ* + *η* + *ϕ*) (pre-diseased, 1/ (*γ*_1_ + *η*_1_ + *ϕ*)) staying in this class, on average *β*_1_/ (*γ* + *η* + *ϕ*) (pre-diseased, *β*_2_/ (*γ*_1_ + *η*_1_ + *ϕ*)) secondary cases.
2. *R*_2_ takes into account the transmission by mild CoViD-19 person. An infectious persons has two routes to reach *Q*_2_: passing through *A* or *D*_1_ (this case is given between parentheses). One infectious person survives during the incubation period with probability *σ* (*σ* + *ϕ*) and enters into asymptomatic (pre-diseased) class with probability *p* (pre-diseased, with 1 − *p*); survives in this class and also is not caught by test with probability *γ*_1_/ (*γ* + *η* + *ϕ*) (pre-diseased, *γ*_1_/ (*γ*_1_ + *η*_1_ + *ϕ*)) and enters into mild CoViD-19 class *Q*_2_ with probability 1 − *χ* (pre-diseased, *m*); and generates, during the time 1/ (*γ*_3_ + *η*_2_ + *ε*_4_ + *ϕ*) staying in this class, on average *zβ*_3_/ (*γ*_3_ + *η*_2_ + *ε*_4_ + *ϕ*) secondary cases.

Hence, 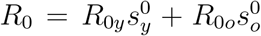 is the overall number of secondary cases generated form one primary case introduced into a completely susceptible subpopulations of young elder persons.

## B Initial conditions and parameters estimation

We present initial conditions supplied to dynamics system, and the estimation of model parameters.

### B.1 Initial conditions

The dynamics of the new coronavirus propagation is obtained by evaluating the system of equations (3), (4) and (6) numerically using the 4^th^ order Runge-Kutta method. Let us determine the initial conditions supplied to this system. In São Paulo State, the number of inhabitants is *N* (0) = *N*_0_ = 44.6 million [15]. The value of parameter *φ* given in Table 1 was calculated by rewriting the equation (A.4) as *φ* = *bϕ*/ (1 − *b*), where *b* is the proportion of elder persons. Using *b* = 0.153 in São Paulo State [15], we obtained *φ* = 6.7 × 10^−6^ *days*^−1^, hence, *N_y_*(0) = *N*_0_*_y_* = 37.8 million (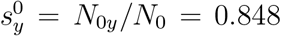) and *N_o_* (0) = *N*_0_*_o_* = 6.8 million (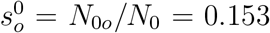). Hence, the initial conditions for susceptible persons are *S_y_* (0) = *N*_0_*_y_* and *S_o_* (0) = *N*_0_*_o_*.

The initial conditions for other variables are calculated based on Table 2. Using *p_y_* = *p_o_* = 0.8, the ratio asymptomatic:symptomatic is 4: 1 for young and elder persons; using *m_o_* = 0.75, the ratio mild:severe CoViD-19 is 3: 1 for elder persons, and for young persons, ratio is 12: 1 from *m_y_* = 0.92. Hence, for elder subpopulation, if we assume that there is one person in *D*_2_*_o_* (the first confirmed case), then there are 3 persons in *Q*_2_*_o_*; the sum (4) is the number of persons in class *D*_1_*_o_*, implying that there are 16 in class *A_o_*, hence, the sum (20) is the number of persons in class *E_o_*. Notice that, if there is 1 person in *D*_2_*_y_*, then there must be 12 persons in *Q*_2_*_y_*. For young subpopulation, we assume that there is not any person in *D*_2_*_y_*, but 6 persons in *Q*_2_*_y_*, then the sum (6) is the number of persons in class *D*_1_*_y_*, implying that there are 24 in class *A_y_*, hence, the sum (30) is the number of persons in class *E_y_*. Finally, we suppose that no one is isolated or tested, and immunized.

Therefore, the initial conditions supplied to the dynamic system (3), (4) and (6) are, for young and elder subpopulations,

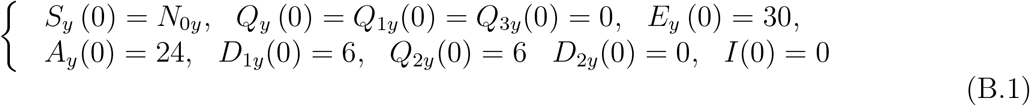

and

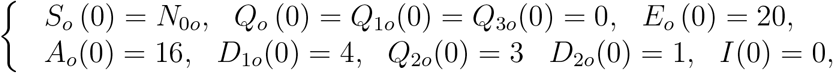

where the initial simulation time *t* = 0 corresponds to the calendar time 26 February 2020, when the first case was confirmed.

### B.2 Parameters estimation

Some values for model parameters are found in literature, other parameters are calculated or reasonable values are assigned, and others are estimated.

For incubation period, we use mean value between 5.2 [13] and 6.4 [5], that is, *σ* = *σ_y_* = *σ_o_* = 1/5.8 *days*^−1^. We use for the infectious rates of pre-diseased persons, *γ*_1_ = *γ*_1_*_y_* = *γ*_1_*_o_* =1/4 *days*^−1^ [2], which is indirectly confirmed by the delay observed in 9 days between low isolation and increase in CoViD-19 cases (see Figure B.1(a)). It was observed approximately 2 weeks for the duration of mild disease, then we use *γ_o_* = 1/14 and *γ_y_* = 1/12 (both in *days*^−1^), and critical disease lasts 2-6 weeks, then we use *γ*_2_*_o_* = 1/21 and *γ*_2_*_y_* = 1/12 (both in *days*^−1^) [19].

From observation that 81% of infections are mild and can recover at home [17], and assuming that the ratio between asymptomatic and symptomatic young and elder persons are equal, we let *p* = *p_y_* = *p_o_* = 4/5 = 0.8. From São Paulo State, 76% of deaths due to CoViD-19 are 60 years old or above, then the ratio of death is 1: 3 to young persons [3]. However, the ratio may be lower in severe CoViD-19 cases, then we assume 2: 3 (in São Bernardo de Campo City, São Paulo State, the ratio of hospitalized young and elder persons is 2: 3.3). We assume that the ratio between asymptomatic and symptomatic among elder persons is 3: 1, hence *m_o_* = 3/4 = 0.75. In order to have ratio 2: 3 between young and elder in hospitalized persons, we must have approximately 10: 1 in the ratio between asymptomatic and symptomatic among young persons. Gathering above information, we calculate approximately

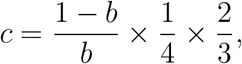

where the term (1 − *b*) /*b* is the populational ratio between young and elder persons, 1/4 is the proportion of severe CoViD-19 cases among elder persons, and 2/3 is the ratio between hospitalized young and elder persons. Using *b* = 0.153, we have *c* = 0.92. Hence, the ratio asymptomatic:symptomatic of young persons is approximately 12: 1, which results in my = 12/13 = 0.92, for *p_y_* = *p_o_*.

In the beginning of epidemics, severe CoViD-19 cases (*D*_2_) were confirmed during hospital care, hence we use *h_y_* = 1.0 and *h_o_* = 1.0. In Wuhan, China [17], 81% of infections did not need hospital care, 14% were severe (developing severe diseases including pneumonia and shortness of breath) and 4.7% were critical (respiratory failure, septic shock, and multi-organ failure). From 4.7%, we use *h*_1_ = 0.05, which is the proportion of hospitalized persons needing ICU care. However, we use higher values for elder and young persons, *h*_1_*_o_* = 0.25 and *h*_1_*_y_* = 0.15. For the ratio ICU:ICU/intubated, we use approximately 14% and 4.7%, resulting in 3: 1, and *h*_2_ = 1/4 = 0.25, which is the proportion of ICU persons needing ICU/intubated care. However, we use higher and lower values for elder and young persons, *h*_2_*_o_* = 0.3 and *h*_2_*_y_* = 0.2. Finally, we assume that proportions not surviving in ICU/intubated are *h*_3_*_y_* = 0.5 and *h*_3_*_o_* = 0.8.

Based on data shown in Figure B.1 (observed accumulated death due to CoViD-19 is shown in Figure 3(a) in the main text), the transmission and fatality rates, proportion in isolated persons, and reduction in the transmission are estimated. Figure B.1 shows the daily (a) and accumulated (b) CoViD-19 cases, plus observed proportions in isolation. In Figure B.1(a), we moved the proportions in isolation showed in Figure B.1(b) 9 days to the right (for instance, the number of cases registered on 10 April corresponds to proportion in isolation observed on 1 April). The horizontal line in Figure B.1(a) corresponds to the mean value *k_mean_* = 0.53, around which daily proportions vary impacting on the transmission. We use the observed proportions in isolated persons [1] and hospitalized CoViD-19 cases [3] in São Paulo State from 24 March to 7 May.

We discuss the collected data roughly (observing the trend of data, but not scientifically based). Interestingly, Figure B.1(a) shows that the daily data present weekly seasonality, with lower cases at the weekend [3], due maybe to registering the day at which occurred the confirmation by laboratory testing, not the beginning of symptoms.

A. The number of SARS in São Paulo State registered in the site of Ministry of Health (Brazil) [11] shows increasing beyond the average cases occurred in past years since 8 March 2020 (around 1000 cases in the 11*^th^* epidemiological week (hereafter, week), 8-14 March), and reach peak 2 weeks later (around 4000 cases in the 13*^th^* week, 22-8 March). After this epidemiological week, the notification as SARS initiates decreasing trend, maybe due to increased testing of severe CoViD-19 cases (on 31 March, there were 822 cases, but one day earlier, only 66 cases, and around 180 cases a day in the 13*^th^* week). Figure B.1(b) shows this jumps up on 31 March. This increased number of cases should be explained by more testing among SARS to identify CoViD-19, or by the exponential-like increasing of epidemics in the beginning, or probably by both. Figure B.1(a) shows an unusual jumps up when comparing 13^th^ week and 14^th^ week (29 March - 4 April), which is not observed in next weeks, suggesting that the isolation decreased the force of infection. Indeed, the isolation was introduced on 24 March, but after 10 days, on 3 April, there is a change in the exponential-like trend, becoming less abrupt. Figure B.1(b) shows increasing trend in blocks of week affected by weekly seasonality shown in Figure B.1(a) depending on the proportion in isolation occurred 9 or 10 days earlier. In a future work, we deal with the depending of the accumulated cases with proportion in isolation delayed in Δ days, that is, Ω (*t* + Δ) = Ω (*t*, *k* (*t* + Δ)).
B. Let us compare roughly severe CoViD-19 cases and isolation week by week. Notice that there is a jumping up from the 13^th^ to 14^th^ week, showing exponential-like increase. However, there is not jumping to the 15^th^ week (5-11 April), possibly showing the effects of isolation. In the 16^th^ week (12-18 April) there was strong variations in the CoViD- 19 cases, maybe due to huge variation in the proportions in isolation 9 days earlier. In the 17^th^ week (19-25 April), the increased number of cases corresponds to decreased proportions in isolation including weekend (on 25 April, Sunday, there was the highest number of cases). This increased trend continued in the next 18^th^ week (26 April - 2 May), when the proportions in isolation fluctuated, but relatively small number of cases was registered during the extended holiday (1-3 May). The behaviour observed in the 17^th^ and 18^th^ weeks may be the effects of manifestation against isolation occurred on 18 April: the peak on 25 April and high number of cases lasting until 30 April, that is, 7 to 12 days after the manifestation is the interval with median 9.5 and variation 2.5 (sum of incubation and pre-diseased periods is 9.8). Should this behaviour be the prolonged effect of one day crowding?

**Figure B.1:**
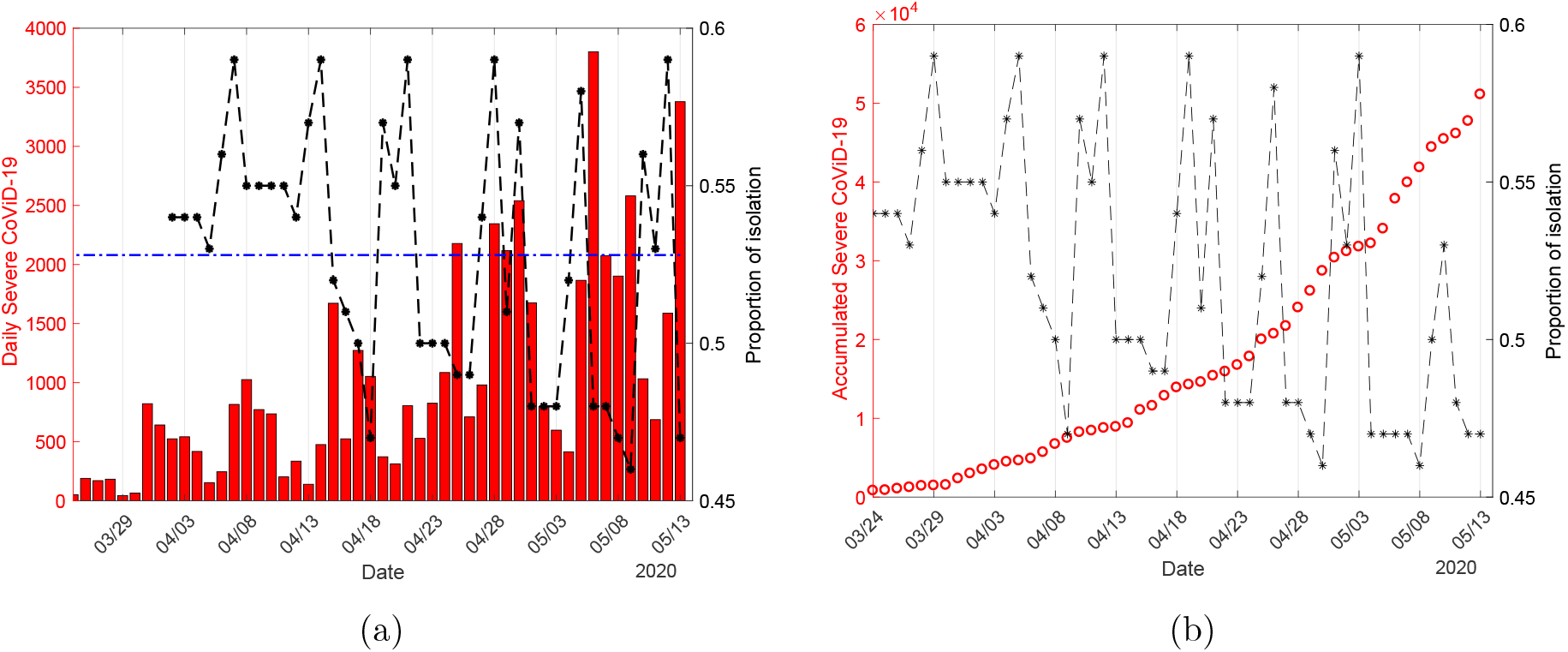
The daily (a) and accumulated (b) CoViD-19 cases collected from 24 March to 3 May [3], plus observed proportion in isolation moved 9 days to the right in (a).

Currently, there is not a sufficient number of kits to detect infection by the new coronavirus. For this reason, test to confirm infection by this virus is done only in hospitalized persons, and also in persons who died manifesting symptoms of CoViD-19. Hence, we have only data of accumulated severe CoViD-19 cases (Ω = Ω*_y_* + Ω*_o_*) and those who died (Π = Π*_y_* + Π*_o_*). We fit the transmission and additional mortality rates taking into account, respectively, severe CoViD-19 and those who died due to CoViD-19. These rates are fitted applying the least square method (see [14]), that is,

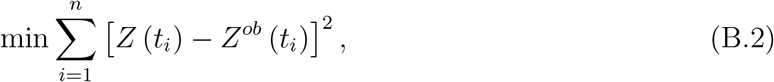

where min stands for the minimum value, *n* is the number of observations, *t_i_* is *i*-th observation time, *Z* stands for Ω given by equation (13), or Π given by equation (14); and *Z^ob^* stands for the observed number of hospitalized persons Ω*^ob^* or number of died persons Π*^ob^*. The fitted parameters are those minimizing the sum of squared differences.

## C Estimating occupancy of beds and deaths

We calculate using values given in Table 3 the occupancy of beds in hospitals, using equations (16), (17) and (18), the number of deaths, using equations (19), (20) and (21), and number of cured persons using equation (22). We present these calculations for current epidemiological status and scenarios of release.

### C.1 Current epidemiological status

The proportions of deaths due to hospitalized CoViD-19 cases are estimated from data collected in São Paulo State using equations (19), (20) and (21), and minimizing the sum

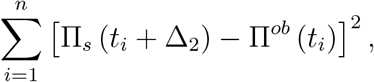

where Π*_s_* = Π_1_ + Π_2_ + Π_3_ is the sum of all deaths in hospital. The better estimation was obtained with Δ_2_ = 15 days using values given in Table 3. This delay is corroborate by data. On 18 May, São Paulo State registered 41 deaths due to CoViD-19, and on 19 May, 324. On 3 and 4 May, 15 and 14 days before 18 May, there were 598 and 415 CoViD-19 cases. On 5 and 6 May, 14 and 13 days before 19 May, there were 1866 and 3800 CoViD-19 cases. We can infer that the number of deaths corresponds to cases of CoViD-19 occurred around 14 to 15 days earlier. For instance, on 13, 14, 15 and 16 May, the daily registered CoViD-19 cases were 3378, 3189, 4092 and 2805, and it is expected elevate number of deaths after 15 days. The estimated curve of deaths using Π*_s_* = Π_1_ + Π + Π_3_ is not shown (similar to that shown in Figure 4(a)).

Figure C.1 shows the estimated curve of Π*_s_* and observed data (a), and extended curve (b), using the delay Δ_2_ = 15 days.

**Figure C.1:**
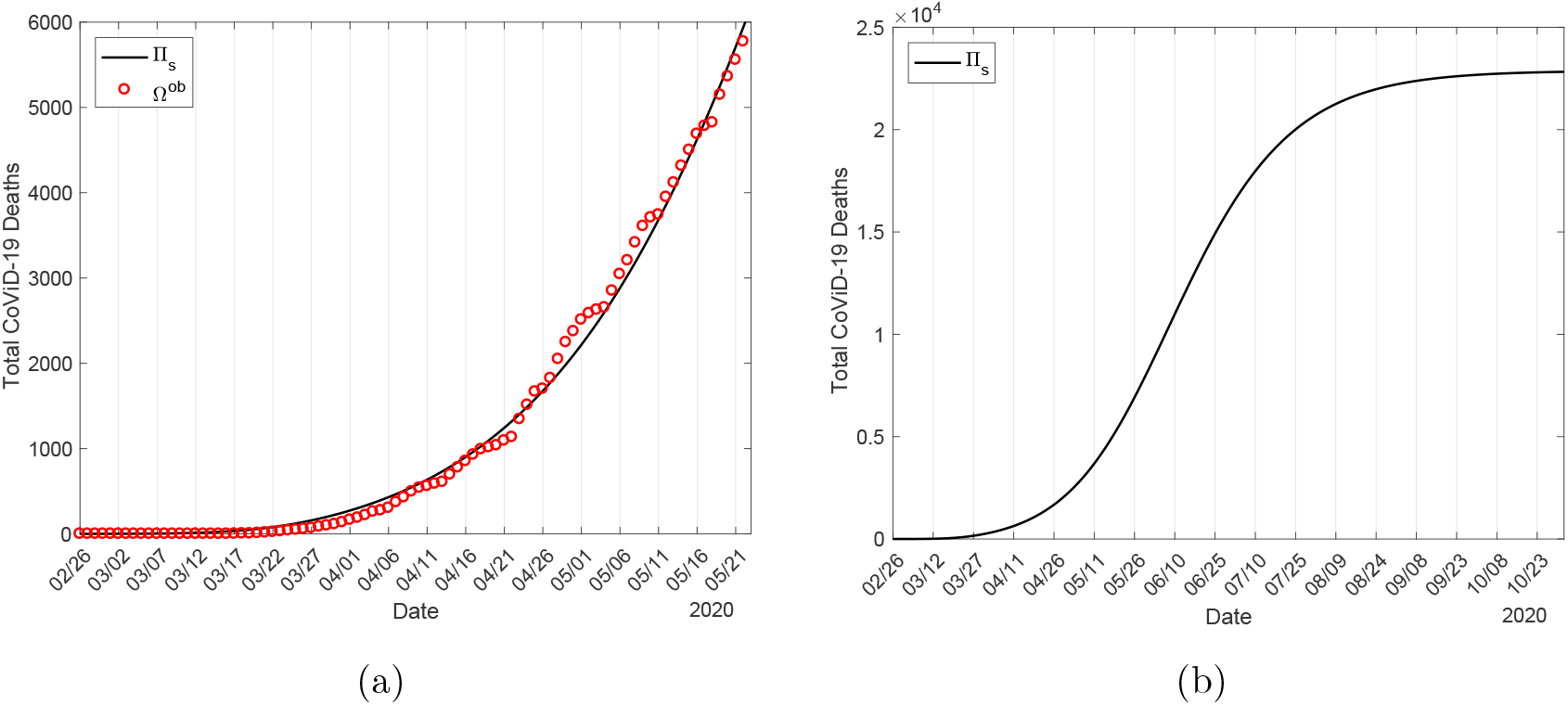
The estimated curve of Π*_s_* and observed data (a), and extended curve (b), using the delay Δ_2_ = 15 days.

Figure C.2 shows the number of beds occupied by young (*j* = *y*) (a) and elder (*j* = *o*) (b) for inpatients (*B*_1_*_j_*), ICU (*B*_2_*_j_*) and ICU/intubated (*B*_3_*_j_*) persons.

**Figure C.2:**
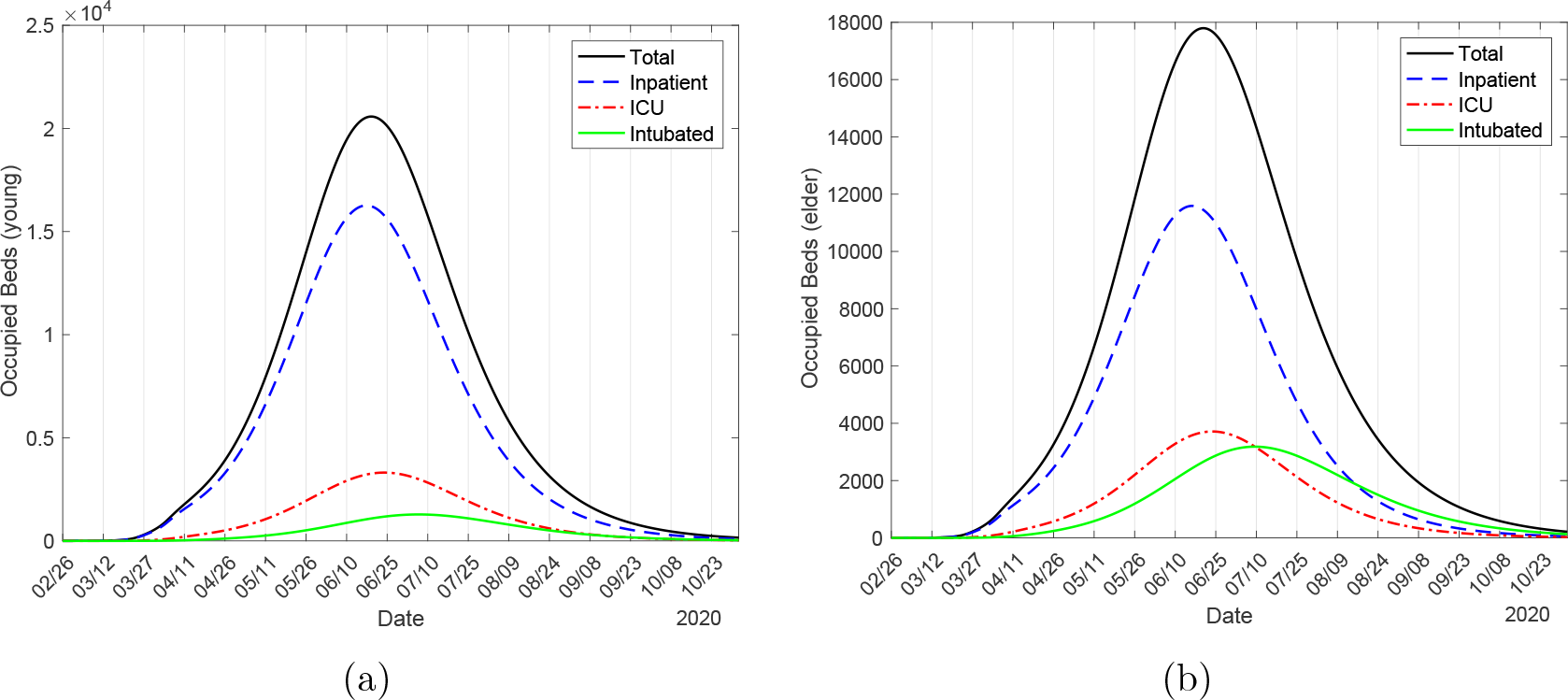
The number of beds occupied by young (*j* = *y*) (a) and elder (*j* = *o*) (b) persons in hospital (*B*_1_*_j_*), ICU (*B*_2_*_j_*) and ICU/intubated (*B*_3_*_j_*).

The peak of numbers of occupied beds for young inpatient, ICU, ICU/intubated and total persons are, respectively, 16260, 3312, 1282 and 20570, which occur on 18 and 23 June, 7 July and 19 June. For elder persons, we have, respectively, 11590, 3714, 3184 and 17790, which occur on 16 and 24 June, 9 July and 20 June. The numbers of occupied beds for all persons is 38360. From Figure 8(a), the peaks of severe CoViD-19 cases *D*_2_ for young, elder and total persons were, respectively, 36510 (56%), 32000 (56%), and 68460 (56%), which occurred on 23, 25 and 23 June. The percentage between parentheses is the ratio between the peaks of occupied beds and severe CoViD-19 cases peak*B*/peak*D*_2_. In comparison with *D*_2_, the peaks of occupied beds for inpatients are anticipated by 5 (young) and 9 (elder) days; for ICU, anticipated by 1 day only for elder, and for ICU/intubated, delayed by 14 (young and elder) days. For all young and elder persons, the peaks of occupied beds are anticipated by, respectively, 4 and 5 days.

Figure C.3 shows the number of deaths due to CoViD-19 for young (*j* = *y*) (a) and elder (*j* = *o*) (b) for inpatient (Π_1_*_j_*), ICU (Π_2_*_j_*) and ICU/intubated (Π_3_*_j_*) persons. At the end of the first wave of epidemics, the numbers of deaths for young persons occurred in hospital, ICU, ICU/intubated and total are, respectively, 2482, 1835, 1094 and 5412. For elder persons, we have, respectively, 7169, 5358, 4953 and 17480. The number of all deaths is 22892.

**Figure C.3:**
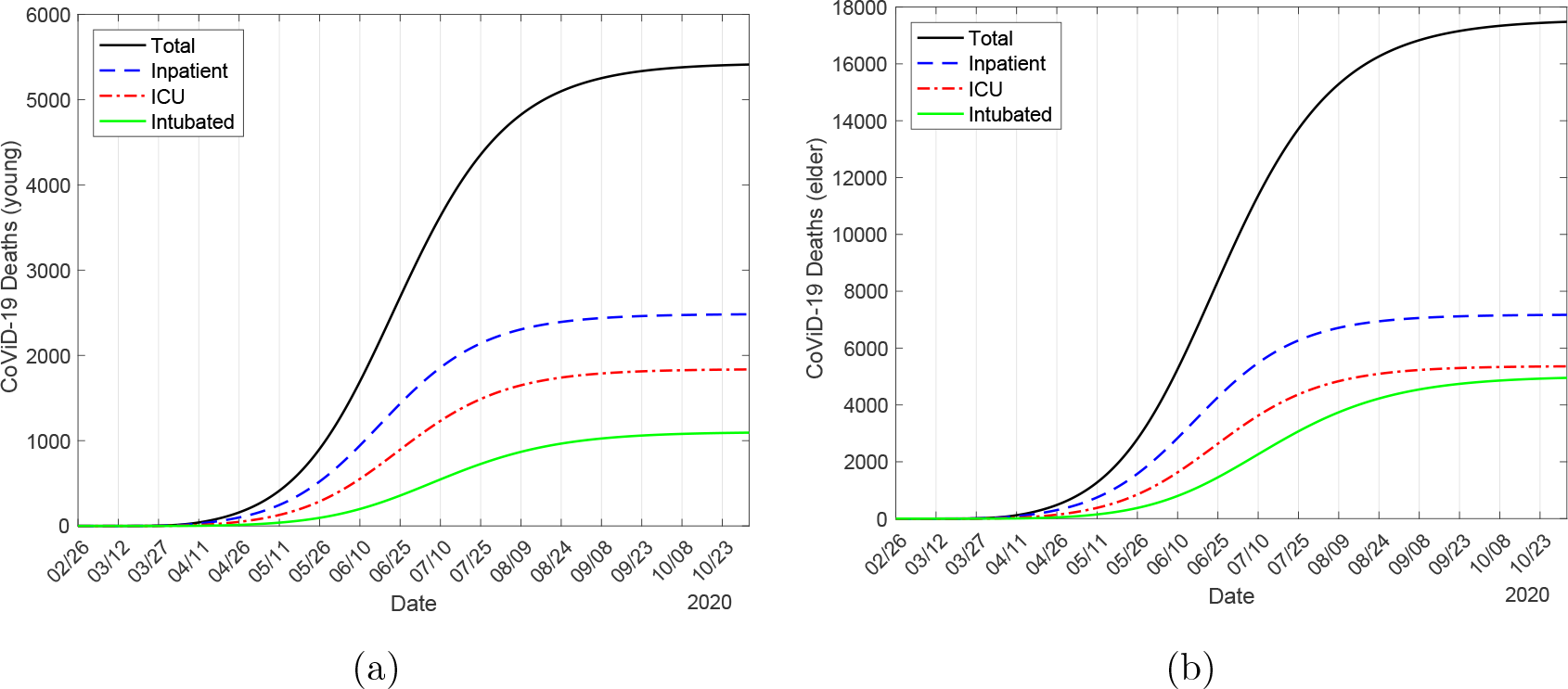
The number of deaths due to CoViD-19 by young (*j* = *y*) (a) and elder (*j* = *o*) (b) persons in hospital (Π_1_*_j_*), ICU (Π_2_*_j_*) and ICU/intubated (Π_3_*_j_*).

At the end of the first wave of epidemics, the numbers of deaths for young persons occurred in hospital, ICU, ICU/intubated and total without any interventions (*k* = 0 and *ε* =1) are, respectively, 6052, 4538, 2720 and 13310. For elder persons, we have, respectively, 17050, 12780, 11890 and 41730. The number of all deaths is 55040. The reduction in the number of deaths by isolation and protective measures in inpatient, ICU, ICU/intubated and total persons are around 40% for young and 42% for elder persons.

We used Δ_1_ = 9 days to estimate the fatality rates. However, if we use Δ_1_ = 15 days, the estimated additional mortality rates are *α_y_* = 0.0009 and *α_o_* = 0.00009 (both in *days*^−1^), resulting at the end of the first wave of epidemics the numbers of deaths for young, elder and total persons, respectively, 263, 2641 and 2904. Hence, the delayed time Δ_1_ for estimating fatality rates is different to the fatality expressed in proportions Δ_2_.

Figure C.4 shows the numbers of cured from CoViD-19 for young *C_y_*, elder *C_o_*, and all persons *C* = *C_y_* + *C_o_* (a) in the beginning of epidemics, and extended curves (b). At the end of the first wave of epidemics, the numbers of cured from CoViD-19 by young, elder, and all persons are, respectively, 237100, 124500, and 361600. The number of cured persons is 94% of all severe CoViD-19 cases Ω = 386700 (Figure 6(b)), and 1580% of deaths at the end of the first wave of epidemics.

**Figure C.4:**
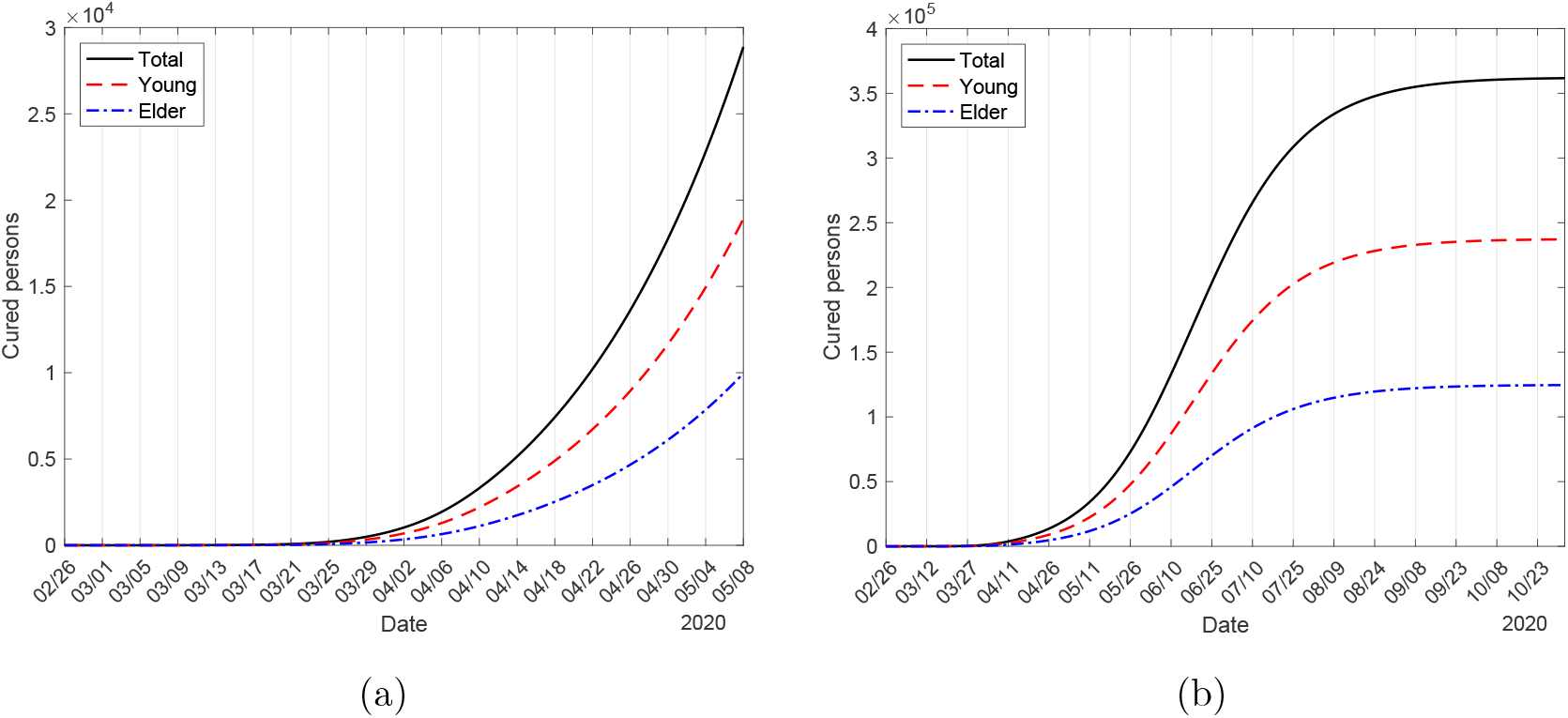
The numbers of cured young, elder and total persons from CoViD-19 (a), and extended curves (b).

On 1 June, the São Paulo State authorities will release isolated persons. Based on estimated parameters, we calculate the numbers of occupied beds and deaths due to CoVid-19 considering three scenarios: without any interventions (*k* = 0 and *ε* =1), isolation alone (k = 0.53 and *ε* =1), and isolation plus protective measures (*k* = 0.53 and *ε*= 0.5). On 1 June, the curves of epidemics without any intervention (peak of the occupancy of beds 252200 on 30 April, 64% of all cases) and with isolation alone (peak of the occupancy of beds 83860 on 18 May, 60% of all cases, 139300) are in descending phase, while curve of isolation and protective measures is in ascending phase (peak of the occupancy of beds 38360 on 20 June, 56% of all cases). In Table C.1 we summarize the decreasing in the number of occupied beds and deaths due to CoVid-19 on 1 June. The percentage between parentheses is the ratio between with and without interventions *B*(*k*, *ε*)/*B*(0, 1) and Π(*k*, *ε*)/Π(0, 1).

Observe that isolation alone decreased occupancy of beds and deaths to 30-45%, while the addition of protective measures decreased to 10-20%. However, comparing with the peaks of occupancy, isolation alone decreased occupancy of beds to 33%, while the addition of protective measures decreased to 15% with respect to without interventions. Comparing the number of deaths at the end of the first wave, isolation alone decreased number of deaths to 47%, while the addition of protective measures decreased to 42%. Hence, we must look not only in the number of deaths, but especially in the capacity of the health care system. However, the surplus of the occupancy of beds in the peak is 213840 for without interventions and 45500 for isolation alone with respect to epidemics with isolation and protection.

**Table C.1:** The summary of the decreasing in the number of occupied beds and deaths due to CoVid-19 on 1 June, with *k_m_* = 0.53. The percentage between parentheses is the ratio between with and without interventions *B*(*k*, *ε*)/*B*(0,1) and Π(*k*, *ε*)/Π(0, 1).

| | $B_{1y}$ | $B_{2y}$ | $B_{3y}$ | $B_y$ | $\Pi_{1y}$ | $\Pi_{2y}$ | $\Pi_{3y}$ | $\Pi_y$ |
| --- | --- | --- | --- | --- | --- | --- | --- | --- |
| $k = 0$<br>$\varepsilon = 1$ | 123700 | 21600 | 6418 | 145500 | 6154 | 4518 | 2397 | 13069 |
| $k = k_m$<br>$\varepsilon = 1$ | 38570<br>(31.2%) | 7331<br>(33.9%) | 2440<br>(33%) | 46970<br>(32.3%) | 2744<br>(44.6%) | 1894<br>(42%) | 831<br>(28%) | 5469<br>(42.1%) |
| $k = k_m$<br>$\varepsilon = 0.5$ | 13490<br>(10.9%) | 2356<br>(10.9%) | 650<br>(10.1%) | 16500<br>(11.3%) | 1139<br>(18.5%) | 683<br>(15%) | 256<br>(10.7%) | 2078<br>(15.9%) |
| | $B_{1o}$ | $B_{2o}$ | $B_{3o}$ | $B_o$ | $\Pi_{1o}$ | $\Pi_{2o}$ | $\Pi_{3o}$ | $\Pi_o$ |
| $k = 0$<br>$\varepsilon = 1$ | 81640 | 21960 | 13760 | 106700 | 17100 | 12620 | 9494 | 39214 |
| $k = k_m$<br>$\varepsilon = 1$ | 26330<br>(32.3%) | 7768<br>(35.4%) | 5401<br>(39.3%) | 36890<br>(34.6%) | 7696<br>(45%) | 5300<br>(42%) | 3201<br>(33.7%) | 16197<br>(41.3%) |
| $k = k_m$<br>$\varepsilon = 0.5$ | 9810<br>(12%) | 2661<br>(12.1%) | 1515<br>(11%) | 13990<br>(13.1%) | 3413<br>(20%) | 2016<br>(16%) | 1034<br>(10.9%) | 6463<br>(16.5%) |

### C.2 Scenarios of release

The occupancy of beds and deaths are evaluated considering releasing of population in isolation divided in three equal releases for the strategies A, B and C, fixing *l*_1_*_j_* = 0.33, *l*_2_*_j_* = 0.5 and *l*_3_*_j_*= 1, *j* = *y*, *o*, and two protective factors, *ε* = 0.5 and 0.2. The shape of figures of the occupancy of beds is quite similar than that shown in Figure 18, as well as the figure of deaths with Figure C.3. Hence we illustrate only the strategy C. For all three strategies, we describe the number of occupied beds and deaths for young (*j* = *y*) and elder (*j* = *o*) persons.

#### C.2.1 Strategy A – Release beginning on 1 June

In strategy A the releases occur on 1, 15 and 29 June with the same protective factor *ε* = 0.5 in isolation being maintained after release.

The peak of numbers of occupied beds by young inpatient (*B*_1_*_j_*), ICU (*B*_2_*_j_*), ICU/intubated (*B*_3_*_j_*) and total (*B_j_*) persons are, respectively, 56360, 11050, 3987 and 70050, which occur on 13, 18, 29 and 14 July. For elder persons, we have, respectively, 37940, 11660, 9215 and 56150, which occur on 13, 19, 31 and 15 July. The peak of all occupied beds, 126200, is 58% of the peak of severe CoViD-19 cases (217600), and 50% of the peak of occupied beds without interventions (252200), but insufficient to avoid the collapse in the health care system.

The number of deaths due to CoViD-19 for young inpatient (Π_1_*_j_*), ICU (Π_2_*_j_*), ICU/intubated (Π_3_*_j_*) and total (Π*_j_*) persons at the end of the first wave of epidemics are, respectively, 5930, 4446, 2666 and 13040. For elder persons, we have, respectively, 16820, 12610, 11760 and 41200. When *ε* = 0.5, all deaths will reach 54240, which is 239% of that without release (22892), and 98.5% of that without any interventions (55040). Notice that the number of deaths due to isolation followed by three releases reduced in only 1.5% in comparison with epidemics without interventions, however, the peak of total number of occupied beds was reduced by around 50%. We stress the fact that the number of deaths considers unlimited health care system, that is, offering treatment for all diseased persons.

Now, we present strategy A with reduced protective factor *ε* = 0.2 adopted by all circulating persons after release.

The peak of numbers of occupied beds by young inpatient, ICU, ICU/intubated and total persons are, respectively, 16680, 3450, 1412 and 21410, which occur on 1, 7, 22 and 3 August. For elder persons, we have, respectively, 12070, 3947, 3697 and 19390, which occur on 4, 8, 23 and 4 August. The peak of all occupied beds (40800), is 55% of the peak of severe CoViD-19 cases (74520). Maybe the health care system does not collapse.

The number of deaths due to CoViD-19 for young inpatient, ICU, ICU/intubated and total persons are, respectively, 4379, 3282, 1963 and 9625. For elder persons, we have, respectively, 13110, 9825, 9128 and 32070. The total number of deaths (41695) is 77% of protective factor *ε* = 0.5, 182% of that without release, and 76% of that without any interventions. From Table 5 and Figure 19, we observe that the decreasing in transmission rates due to increased protective measures (factor ε is decreased) becomes the epidemiological scenarios with releases less harmful, decreasing by around 77% when ε decreases from 0.5 to 0.2. Lower ε decreases the transmission rates, reducing *R_ef_*.

#### C.2.2 Strategy B – Release beginning on 23 June

In strategy B the releases occur on 23 June 23, 7 and 21 July with the same protective factor *ε* = 0.5 in isolation being maintained after release.

The peak of numbers of occupied beds for young inpatient, ICU, ICU/intubated and total persons are, respectively, 43180, 8528, 3163 and 53950, which occur on 7, 12, 23 and 8 August. For elder persons, we have, respectively, 29130, 9017, 7418 and 43780, which occur on 7, 13, 25 and 8 August. The peak of all occupied beds (97730) is 57% of the peak of severe CoViD-19 cases (170100), which is 77% of the peak observed in strategy A, but insufficient to avoid the collapse in the health care system.

The number of deaths due to CoViD-19 for young inpatient, ICU, ICU/intubated and total persons are, respectively, 5876, 4406, 2642 and 12920. For elder persons, we have, respectively, 16700, 12520, 11680 and 40900. The total number of deaths 53820 is 99% of strategy A, 235% of that without release (22892), and 98% of that without any interventions (55040).

Now, we present strategy B with reduced protective factor *ε* = 0.2 adopted by all circulating persons after release.

The peak of numbers of occupied beds by young inpatient, ICU, ICU/intubated and total persons are, respectively, 16260, 3312, 1282 and 20570, which occur on 18, 23 June, 7 July and 19 June. For elder persons, we have, respectively, 11590, 3714, 3184 and 17790, which occur on 16, 24 June, 9 July and 20 June. The peak of all occupied beds (38360) is 56% of the peak of severe CoViD-19 cases (68450), which is 94% of the peak observed in strategy A. Maybe the health care system does not collapse.

The number of deaths due to CoViD-19 for young inpatient, ICU, ICU/intubated and total persons are, respectively, 4147, 3108, 1861 and 9115. For elder persons, we have, respectively, 12410, 9298, 8650 and 30350. The total number of deaths (39465) is 73% of protective factor *ε* = 0.5, 95% of strategy A, 172% of that without release, and 72% of that without any interventions.

The number of deaths decreased by around 72% when *ε* decreases from 0.5 to 0.2, while in comparison with strategy A, very small decreasing by around 95%.

#### C.2.3 Strategy C – Release beginning on 6 July

In strategy C the releases occur on 6, 20 July and 3 August with the same protective factor *ε* = 0.5 in isolation is maintained after release.

Figure C.5 shows the number of occupied beds due to CoViD-19 by young (*j* = *y*) (a) and elder (*j* = *o*) (b) for inpatients (*B*_1_*_j_*), ICU (*B*_2_*_j_*) and ICU/intubated (*B*_3_*_j_*) persons.

**Figure C.5:**
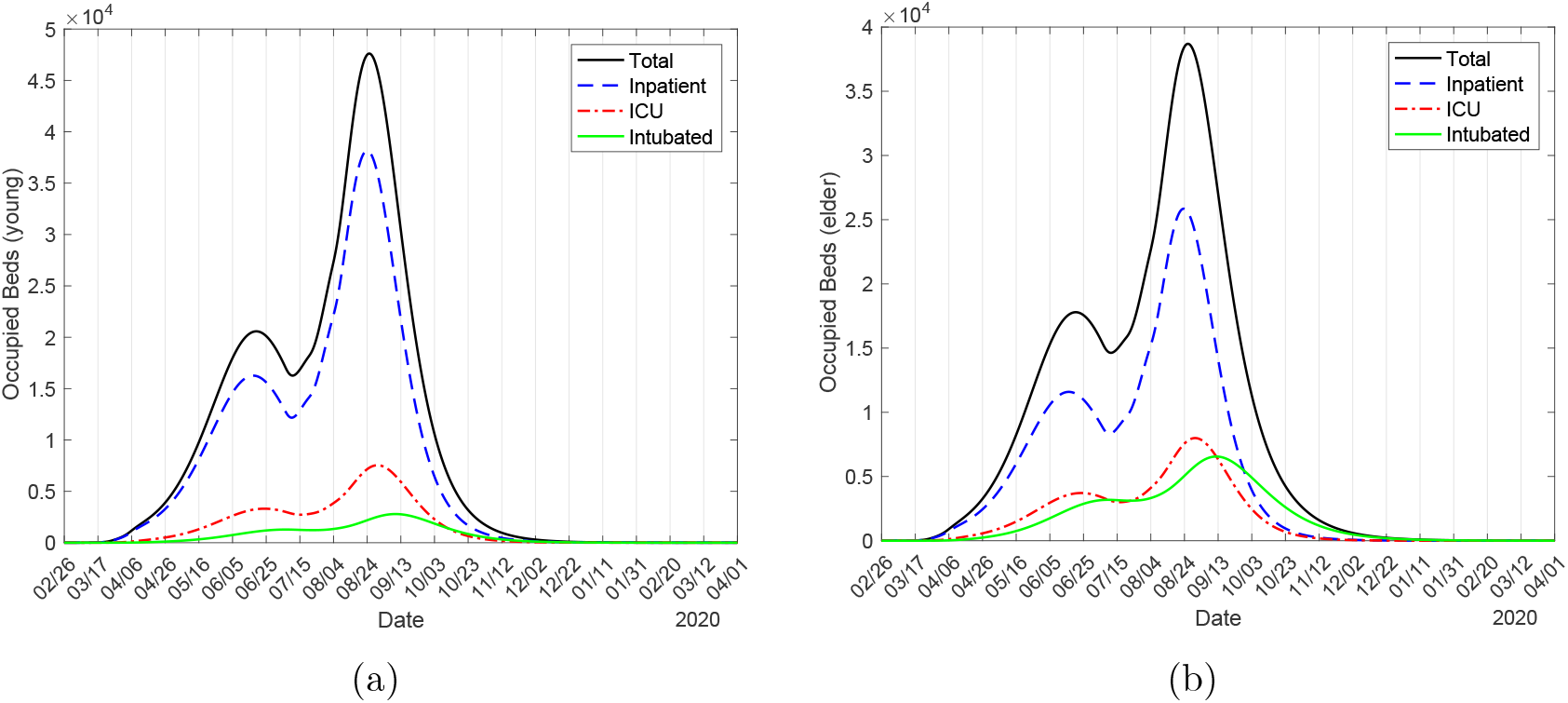
The number of occupied beds due to CoViD-19 by young (*j* = *y*) (a) and elder (*j* = *o*) (b) for inpatients (*B*_1_*_j_*), ICU (*B*_2_*_j_*) and ICU/intubated (*B*_3_*_j_*) persons. Strategy C with *ε* = 0.5, release beginning on 6 July.

The peak of numbers of occupied beds for young inpatient, ICU, ICU/intubated and total persons are, respectively, 38150, 7531, 2790 and 47610, which occur on 23, 30 August, 9 September and 25 August. For elder persons, we have, respectively, 25850, 7993, 6555 and 38690, which occur on 23, 30 August, 11 September and 26 August. The peak of all occupied beds (86300) is 58% of the peak of severe CoViD-19 cases (150100), and this peak of all occupied beds is 63% and 88% of the peaks observed in strategies A and B, respectively, but insufficient to avoid the collapse in the health care system.

Figure C.6 shows the number of deaths due to CoViD-19 for young (*j* = *y*) (a) and elder (*j* = *o*) (b) inpatient (Π_1_*_j_*), ICU (Π_2_*_j_*) and ICU/intubated (Π_3_*_j_*) persons. At the end of the first wave of epidemics, the numbers of deaths for young persons occurred in hospital, ICU, ICU/intubated and total are, respectively, 5846, 4383, 2629 and 12860. For elder persons, we have, respectively, 16630, 12470, 11630 and 40740. The total number of deaths (53600) is 98.8% and 99.6% of strategies A and B, respectively, 234% of that without release (22892), and 97% of that without any interventions (55040).

**Figure C.6:**
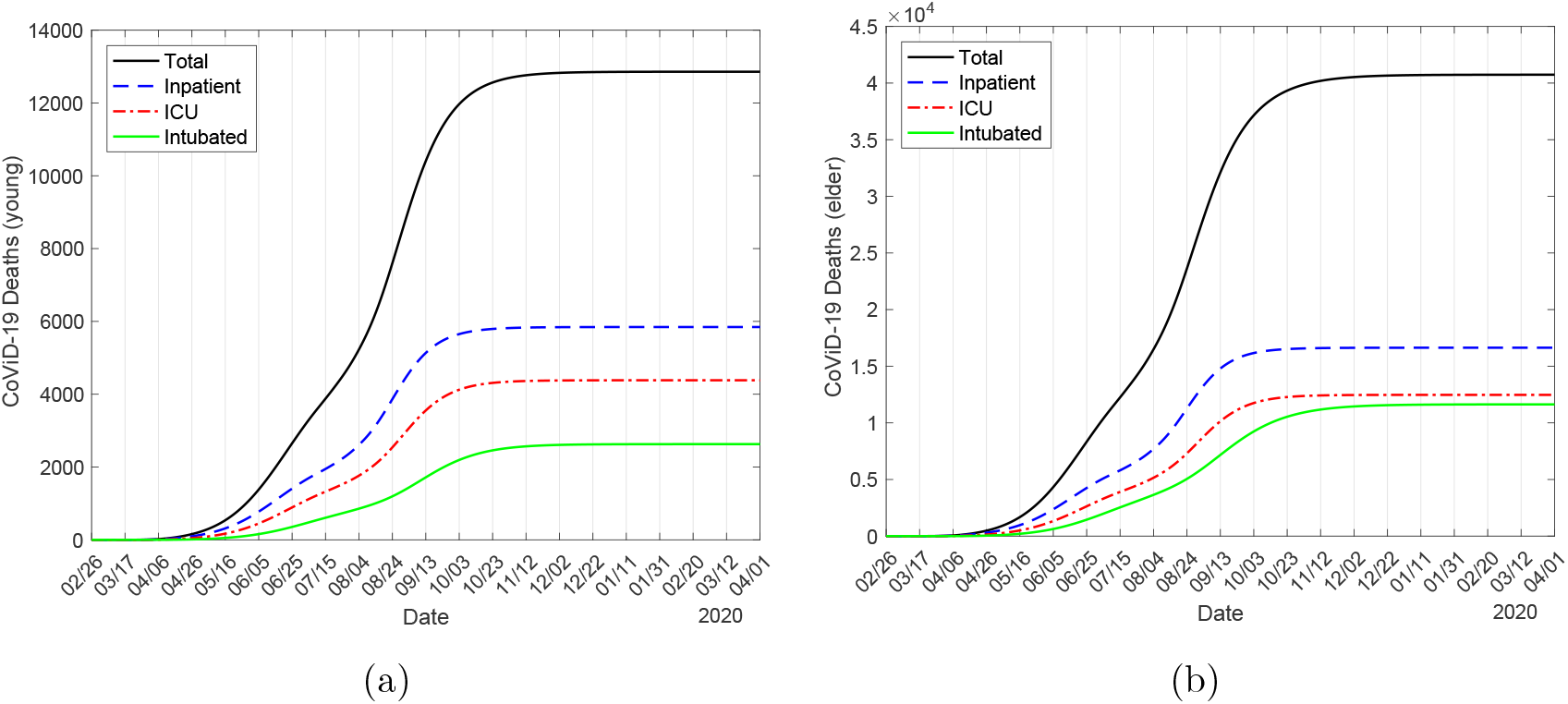
The number of deaths due to CoViD-19 by young (*j* = *y*) (a) and elder (*j* = *o*) (b) for inpatient (Π_1_*_j_*), ICU (Π_2_*_j_*) and ICU/intubated (Π_3_*_j_*) persons. Strategy C with *ε* = 0.5, release beginning on 6 July.

Now, we present strategy C with reduced protective factor *ε* = 0.2 adopted by all circulating persons after release (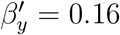 and 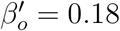 (both in *days*^−1^), giving *R*_0_ = 1.85). Figure C.7 shows the number of occupied beds due to CoViD-19 by young (*j* = *y*) (a) and elder (*j* = *o*) (b) for inpatients (*B*_1_*_j_*), ICU (*B*_2_*_j_*) and ICU/intubated (*B*_3_*_j_*) persons.

**Figure C.7:**
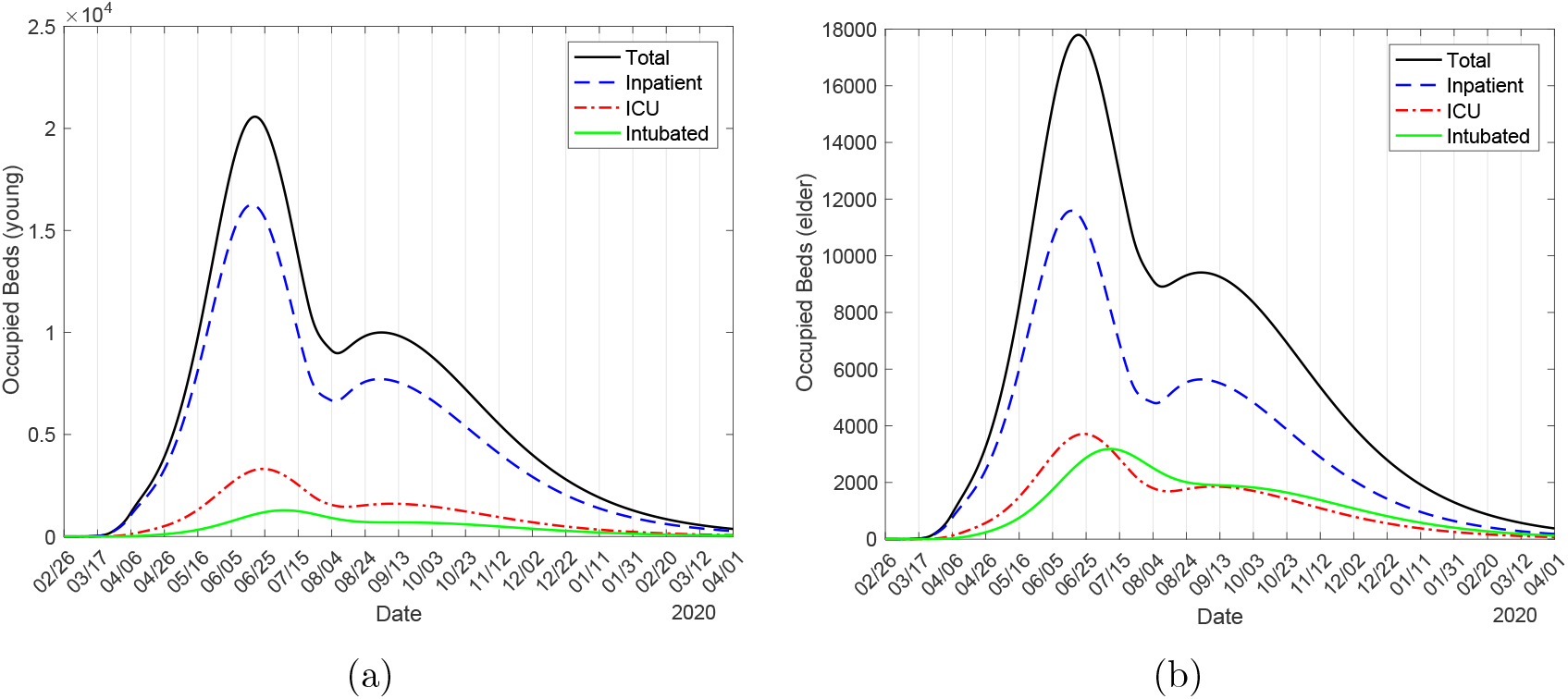
The number of occupied beds due to CoViD-19 by young (*j* = *y*) (a) and elder (*j* = *o*) (b) for inpatients (*B*_1_*_j_*), ICU (*B*_2_*_j_*) and ICU/intubated (*B*_3_*_j_*) persons. Strategy C with *ε* = 0.2, release beginning on 6 July.

The peak of numbers of occupied beds by young inpatient, ICU, ICU/intubated and total persons are, respectively, 16260, 3312, 1282 and 20570, which occur on 18, 23 June, 7 July and 19 June. For elder persons, we have, respectively, 11590, 3714, 3184 and 17790, which occur on 16, 24 June, 9 July 20 and June. The peak of all occupied beds (38360) is 56% of the peak of severe CoViD-19 cases (68450), which is 94% and 100% of the peak observed in strategies A and B, respectively. Maybe the health care system does not collapse.

Figure C.8 shows the number of deaths due to CoViD-19 for young (*j* = *y*) (a) and elder (*j* = *o*) (b) inpatient (Π_1_*_j_*), ICU (Π_2_*_j_*) and ICU/intubated (Π_3_*_j_*) persons. At the end of the first wave of epidemics, the numbers of deaths for young persons occurred in hospital, ICU, ICU/intubated and total are, respectively, 3957, 2964, 1771 and 8692. For elder persons, we have, respectively, 11830, 8854, 8216 and 28900. The total number of deaths (37592) is 70% of protective factor *ε* = 0.5, 90% and 95% of strategies A and B, respectively, 164% of that without release, and 68% of that without any interventions.

**Figure C.8:**
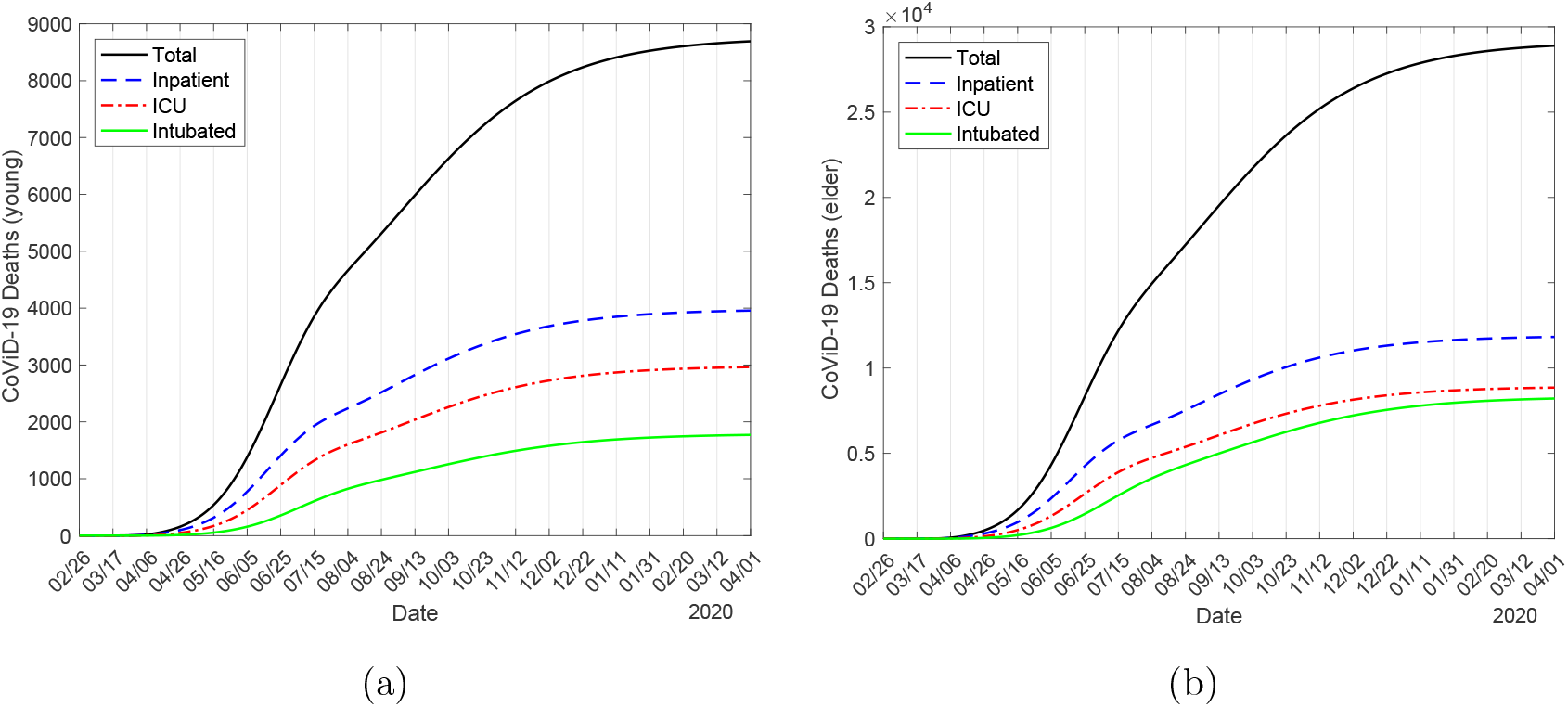
The number of deaths due to CoViD-19 by young (*j* = *y*) (a) and elder (*j* = *o*) (b) for inpatient (Π_1_*_j_*), ICU (Π_2_*_j_*) and ICU/intubated (Π_3_*_j_*) persons. Strategy C with *ε* = 0.2, release beginning on 6 July.

The little difference in the number of deaths in strategies A, B and C can be explained by *R_ef_* situating around one at the time of the first release.

## Footnotes

1 Simulations were done on 7-8 May.

2 Simulations were done on 9-11 May.

